# Serum Concentration of Continuously administered Vancomycin influences Efficacy and Safety in Critically Ill Adults: A Systematic Review

**DOI:** 10.1101/2022.10.20.22280821

**Authors:** Katrin Viertel, Elisabeth Feles, Melanie Schulte, Thorsten Annecke, Frauke Mattner

## Abstract

Vancomycin is an antibiotic to treat gram-positive infections in critically ill adults. For continuously administered vancomycin (CI), various target ranges have been used in the past, ranging from 15-20 mg/L to 30-40 mg/L. This systematic literature review was conducted to investigate the impact of steady-state serum concentration (Css) of CI on the safety and efficacy of therapy in critically ill adults. According to the PRISMA statement, relevant literature was identified by searching two electronic databases (PubMed, Cochrane Library) and Google Scholar from inception until July 2023, focussing on studies reporting measured Css and treatment outcomes (e.g. mortality, nephrotoxicity) with CI. Due to the heterogeneity of the studies, a narrative synthesis of the evidence was performed. Twenty-one publications were included with a total of 2,949 patients with CI (pts). Mortality was higher (two studies, n=388 pts) and clinical cure lower (one study, n=40 pts) with a Css <15 mg/L measured 24 hours after initiation of CI (C24). An adequate loading dose appeared most important for maintaining higher C24. Generally, higher Css were associated with higher rates of acute kidney injury (AKI) (fifteen studies, n=2,331 pts). We calculated that a Css <25 mg/L (vs. ≥25 mg/L) was preferable for reducing nephrotoxicity (three studies, n=515 pts). Despite sparse data availability, the target range of 15-25 mg/L in CI may increase clinical cure and reduce mortality and AKI. In future research, vancomycin Css cohorts should be formed to allow evaluation of the impact of Css of CI on treatment outcomes.

## 1 Introduction

Infections in intensive care units (ICU) are highly prevalent. More than half of ICU patients may become infected during their ICU stay [1], and between 8-22% of infections are acquired nosocomially [1, 2]. Infections with gram-positive pathogens regularly occur in critically ill patients [3–5] causing one third (21-67%) of all infections [6]. Available since the 1950s, the glycopeptide vancomycin remains an important antibiotic for the treatment of infections with gram-positive bacteria, particularly methicillin-resistant *Staphylococcus aureus* (MRSA), coagulase-negative staphylococci (CoNS) and *Enterococcus faecium* [7–11]. It is also frequently used in sepsis therapy as empirical treatment in combination with beta-lactam antibiotics in areas with high prevalence of MRSA [12]. Physiological changes caused by critical illness may often alter drug excretion and lead to inappropriate drug levels; renal insufficiency particularly affects drug elimination [13–15]. Additionally, vancomycin itself can impair renal function by inducing acute tubulointerstitial nephritis [16, 17] or acute tubular necrosis [18] leading to acute kidney injury (AKI). AKI worsens the outcome of the ICU stay [19, 20]. The appropriate dosage of vancomycin has therefore been under discussion for some time. Vancomycin is administered via intermittent infusion (II) or continuous infusion (CI). CI appears to hold several advantages over II. These include a lower potential risk of AKI [21–25], earlier or higher target attainment [24–35], less variability in serum concentrations [26, 32, 34, 36] and easier and less expensive monitoring of drug levels [26, 29, 31, 34]. Several studies have been published in the past with CI using multiple target ranges of vancomycin steady-state serum concentration (Css). These were (indication-independent) 15-20 mg/L [17, 31, 37–40], 15-25 mg/L [29, 30, 32, 41–47], 20-25 mg/L [33, 34, 48–50], 20-30 mg/L [26, 51–61], 25-30 mg/L [62], 20-40 mg/L [61] and 30-40 mg/L [63], respectively. As far as we are aware, there is a lack of a comprehensive comparative evaluation of the influence of vancomycin serum concentration during continuous infusion of vancomycin on therapeutic outcomes of efficacy (e.g. clinical and microbiological outcomes such as mortality or cure) and safety (e.g. nephrotoxicity such as AKI) in critically ill adult patients. Therefore, this review examines the current knowledge on the impact of Css on the efficacy and safety of CI in critically ill adults in the ICU.

## 2 Materials and Methods

A systematic review was carried out in accordance with the PRISMA guidelines (preferred reporting items for systematic review and meta-analysis) [64].

### 2.1 Selection Criteria

According to the PICOS questions (population, intervention, comparison, outcomes and study design), enrollment of papers was performed as follows:

- **Population**: Adults (age ≥18 years) having been treated with vancomycin in an ICU.
- **Intervention**: Continuous infusion of vancomycin.
- **Comparison**: Vancomycin with or without: intermittent infusion, chronic kidney injury (CKD), renal replacement therapy (RRT), obesity or different dosing regimens.
- **Outcomes**: Vancomycin steady-state serum level (about 24 hours after the start of therapy (C24), average serum level during the entire duration of therapy or at least three days of therapy (Cmean), area under the serum concentration-time curve for 24 hours (AUC24)) and efficacy or safety. Efficacy was defined as clinical or microbiological success or failure (e.g. survival or mortality, cure or relapse). Safety was defined as occurrence of nephrotoxicity (e.g. AKI).
- **Study design**: Clinical trials and observational studies.

Studies with the following criteria were excluded: Nonhuman data, paediatric patients (age <18 years), records without intravenous vancomycin application, records without CI, non-ICU setting, lacking efficacy or safety data, omitted vancomycin serum levels, case reports, comments, editorials, reviews and meta-analyses. In addition, investigations with less than 30 patients with CI, studies with a duration of less than one year and studies assessing fewer patient records than patients were included in the study were excluded from our analysis. Exclusion based on the language of publication was not performed.

### 2.2 Data Sources

A literature search from inception until 15 July 2023 in two electronic databases (MEDLINE through the PubMed interface, Cochrane Library) and Google Scholar was performed using the combined search terms *“vancomycin”* and “*continuous*” (PubMed: (vancomycin[Title]) AND (continuous), Cochrane Library: Title Abstract Keyword: vancomycin; AND All Text: continuous, Google Scholar: allintitle: vancomycin continuous).

### 2.3 Study Selection

Initially, the exclusion of doubles was carried out. Thereupon, articles were selected based on the information obtained from the title and abstract according to the inclusion criteria. Pertinent articles or those not providing sufficient information via title or abstract were evaluated in full text. Finally, the selected articles were critically read in total (Figure 1).

### 2.4 Quality Assessment

The quality of the studies selected for inclusion was rated using two tools: The Newcastle-Ottawa Scale (NOS) for non-randomized studies [65, 66] and the Cochrane risk of bias tool for randomized trials (RoB 2) [67]. Using the NOS a “star system” described the suitability of the selection of the study groups (one star maximum each), the comparability of the groups (two stars maximum) and the ascertainment of the exposure or outcome of interest (one star maximum each). The total number of stars of each study was interpreted as “good quality” at 8-9 stars, “fair quality” at 6-7 stars, “questionable quality” at 4-5 stars, “poor quality” at 2-3 stars and “serious risk of bias” at 0-1 stars. With the RoB 2 scale, risk of bias was categorized by “low risk”, “high risk” or “some concerns”, with “low risk” of overall bias interpreted as “good quality,” “high risk” as “serious risk,” and “some concern” as “questionable quality”. Due to the heterogeneity of the study designs (e.g. inclusion or exclusion of patients with chronic kidney disease) and outcome measures (e.g. use of different definitions for AKI or mortality), it was not possible to summarize the results in a meta-analysis. In accordance with the Cochrane Consumers and Communication Review Group guideline [68], we therefore performed a narrative synthesis of the evidence. To minimize the influence of seasonal variations in infection type and frequency on study results (selection bias) [69–71], investigations with a duration of less than one year or a population number of less than 30 patients with CI were excluded from our analysis. Attrition bias from publications that analyzed fewer patient records than were included in the study was prevented by excluding them from our evaluation. Because of the inhomogeneous coverage of potential factors affecting outcome, such as disease severity or concomitant nephrotoxin use, some degree of performance and reporting bias was to be expected. Language bias was avoided by including all languages of publication. The free internet translation programme www.Deepl.com/Translator was used for translation where necessary.

### 2.5 Data Extraction

The main characteristics of the included studies were outlined in five tables, which are accessible online via the supplementary material: design, type of study and main objective (Table S1), characteristics of the study population (Table S2), information regarding the treatment with vancomycin (Table S3), main findings in terms of efficacy (Table S4) and safety (Table S5). “*Efficacy*” connotes clinical or microbiological cure, improvement, persistence, progression, relapse or reinfection (further definitions displayed in Table S11), mortality/survival and target attainment, while “*safety*” stands for extrarenal but mainly nephrotoxic adverse events (Table 1). Some of the values given have been calculated by us in the presence of sufficient numerical data.

### 2.6 Analysis

The reported average Css were primarily divided into three categories: Vancomycin serum level on the second day, i.e. approximately 24 hours after initiation of CI (C24), mean vancomycin serum level during the entire duration of CI or at least three days of CI (Cmean) and area under the serum concentration-time curve for a period of 24 hours during CI (AUC24). The mortality rate was differentiated into “in-hospital”, “ICU”, “infection-related,” “x-days” (e.g. 28-days), “end of therapy” or “not reported”. Likewise, nephrotoxicity in the form of AKI was classified according to Table 1 in “AKIN”, “KDIGO”, “RIFLE”, “Rybak2009”, “Rybak2020”, “RRT” or “other”. To compare the reported Css, in cases where no mean but only the median was stated, the mean and standard deviation were calculated according to the method described by Wan et al.: *mean=(median+lower quartile+upper quartile)/3* [72]. In order to use only one value per study for the comparison, weighted means of serum concentration, mortality, nephrotoxicity or target attainment rates were calculated if values were given only for subgroups but not the entire CI population of a study. Because individual values of the study population were not available, no relationship between the variables could be tested by regression analysis. But to give an impression of interdependence, average Css, i.e. C24, Cmean, AUC24, were plotted against the corresponding percentage rates of clinical failure (mortality, persistence or progression of infection symptoms), clinical success (cure, improvement of infection symptoms), microbiological failure (persistence, escalation, relapse), microbiological success (cure), AKI or target attainment, respectively. Additionally, scatterplots were created comparing rates of target attainment to mortality or AKI, mortality to AKI and target attainment to target range. For comparison of the different cohorts and based on the parameters identified as significant predictors for outcomes in the included studies (Table 3), the dosing (planned loading dose (pLD) and planned maintenance dose (pMD), actually applied loading dose (aLD) and actually applied maintenance dose (aMD)), length of therapy with vancomycin (LOT), age, gender distribution, body weight (BW), kidney status at baseline (serum creatinine (SCr), creatinine clearance (CrCl)), amount of patients with sepsis, severity of illness at start of CI (APACHE II score (Acute Physiology and Chronic Health Evaluation score), SAPS II (Simplified Acute Physiology Score), SOFA score (Sepsis-related Organ Failure Assessment score)), amount of patients with mechanical ventilation and concomitant reported nephrotoxins were added to the scatterplots. The relative risk, its confidence interval, z-value and p-value for AKI at Css above or below 25 mg/L were calculated by the formulas described by Altman et al. [73] and displayed in a forest plot. The collected data were summarized and graphically plotted using Microsoft Excel (2016) and the *ggplot2* and *epitools* packages of the R Statistical Software (v4.2.0; R Core Team 2021).

## 3 Results

### 3.1 Bibliographic Search

Across the various databases, 1,770 articles were identified (915 from PubMed, 284 from Cochrane databases, 456 from Google Scholar and 115 from a manual search in the reference lists of related publications). 465 duplicate studies were excluded, leaving 1,305 records for further investigation. A total of 1,177 publications were classified as inappropriate according to the PICOS criteria after inspection of the title and abstract, mainly because of the absence of intravenous or continuous use of vancomycin. Of the remaining 128 full-text articles, twenty-one were evaluated for data extraction and inclusion in this systematic review. Eighteen reported efficacy data and sixteen presented safety data. Figure 1 shows the selection process.

### 3.2 Quality of Included Studies

The methodological quality of the studies included in this review varied, but did not influence inclusion in the analysis. The detailed results of the risk of bias assessment are shown in Supplementary Table S8 and Table S9.

### 3.3 Characteristics of Included Studies

Only two randomized controlled trials met the inclusion criteria [34, 47]. Two of the included studies were multicenter investigations [34, 46]. One-third had a prospective design [34, 41, 47, 52, 53, 55, 74, 75]. The studies were conducted between 2001 and 2020 and the majority (16/21) was performed in Europe [33, 34, 43, 44, 46–48, 52, 53, 55, 56, 58, 59, 61, 74, 76]. In two-thirds of the studies different patient groups were compared [33, 34, 42–44, 46, 47, 49, 53, 56, 61, 74, 76]. Equally, in thirteen studies only patients on CI were included [41–43, 46, 48, 52, 53, 55–59, 75]. In total 2,949 patients treated with CI were enrolled in the trials. In two-fifths of the studies only patients with sepsis were included [43, 52, 53, 55–59, 74], where different definitions were used [77–80]. Positive cultures were described in almost half of the studies [46, 48, 52, 55, 56, 58, 59, 61, 74, 75], which ranged from 6% to 100%. In two studies it was specialized in infections caused by beta-lactam-resistant pathogens (i.e. MRSA, methicillin-resistant CoNS) [34, 47]. Additional use of nephrotoxins was described in about two fifth of the publications [33, 34, 43, 46, 48, 56, 61, 76]. Aminoglycosides were listed most frequently (7/8) [33, 34, 43, 46, 48, 56, 61]. The heterogeneous characteristics of the included studies can be seen in Table 2 or in more detail in Supplementary Table S1 and Table S2.

### 3.4 Characteristics of Vancomycin Treatment

In general, the vancomycin therapy consisted of a loading dose (LD) and a maintenance dose (MD). The dosing regimen was differentiated according to a fixed or body weight-dependent protocol, if necessary with adaptation according to renal function. Mean LD ranged from 500 mg [74] to 2,894 mg [42] and from 8 mg/kg BW [74] to 35 mg/kg BW [52]. The average daily MD ranged from 396 mg/d or 5 mg/kg BW/d [43] to 3,039 mg/d or 42 mg/kg BW/d [42]. A total dose throughout the course of therapy was reported in five studies and ranged from 3.6 g to 14 g [43, 47, 56, 61, 76]. In seventeen studies the average duration of vancomycin therapy was reported [33, 34, 41–44, 46–49, 55, 56, 58, 59, 61, 75, 76]. It ranged from 3 days [55] to 15 days [46] (mean 6 days, IQR 5-9 days). Different Css ranges were aimed for, but no indication dependence was evident: 20-30 mg/L (n=7 [52, 53, 55–59]), 15-25 mg/L (n=6 [41–44, 46, 47]), 20-25 mg/L (n=4 [33, 34, 48, 49]), 20-40 mg/L (n=1 [61]), 20 mg/L (n=1 [74]). In half of the studies, average C24 were reported. They were distributed as follows: 15-<20 mg/L (n=2 [56, 74]), 20-<25 mg/L (n=9 [41, 42, 48, 52, 53, 55, 57–59]). No study had an average C24 <15 mg/L or ≥25 mg/L (Supplementary Figure S6a). In two thirds of the studies, Cmean were stated or could be calculated. They were distributed as follows: 15-<20 mg/L (n=3 [44, 47, 76]), 20-<25 mg/L (n=6 [34, 41, 49, 56, 59, 61]), 25-<30 mg/L (n=2 [33, 58]), ≥30 mg/L (n=1 [75]) (Supplementary Figure S6b). Different kinds of immunoassays were used for measurement, which measured the total vancomycin amount in the serum. In seven studies an average AUC24 (total) value was presented [33, 34, 42, 43, 52, 55, 75], which was most often calculated by a (log-) trapezoidal rule (n=4). Mean AUC24(/MIC) data ranged from 477 mg*h/L [43] to 788 mg*h/L [75] (Supplementary Figure S6c). The parameters for treatment with vancomycin are depicted in Table 2 and in Supplementary Table S3.

#### 3.4.1 Target Attainment

In half of the studies a target attainment (TA) rate was reported (n=1,391 pts; Table 2: 24-92%) [41–43, 48, 49, 52, 53, 55, 57–59]. Additionally, in one third each the rate of subtherapeutic and supratherapeutic levels was described [41, 44, 49, 55, 57–59]. In no study a dependence of TA on Css per se was detected, but a higher vancomycin dose increased TA (Table 3) [58, 59]. We noticed that a lower and wider target range was achieved more often (Figure 2). In no study the *time within* the target range was reported, but in three studies a *time required to reach* the target Css was stated, a link to efficacy would be questionable. It took 16 hours (target range 20-25 mg/L, ICU mortality 21%) [33], 36 hours (target range 20-25 mg/L, ICU mortality 37%) [34] and 48 hours (target range 20-30 mg/L, ICU mortality 30%) [59].

### 3.5 Characteristics of Outcome Parameters

Css-dependent efficacy and safety of CI were analyzed. However, there was not a single study whose primary objective was the target concentration range-dependent comparison of outcome parameters. Results are displayed in Table 2 and in Supplementary Table S4 and Table S5.

#### 3.5.1 Efficacy

Efficacy was mentioned in eighteen studies and could be divided into clinical or microbiological treatment failure or success (n=2,648 patients with CI (pts)) (Table 2 or Supplementary Table S4, Table S10 or Table S11) [33, 34, 41–43, 46–49, 52, 55–59, 61, 74, 76]. Only for C24 <15 mg/L could a significant association with in-hospital mortality or clinical cure be shown (n=388 pts) [43, 74]. No other statistically significant associations were found between Css and clinical or microbiological success or failure. Five studies compared cohorts with different Css and treatment effectiveness. Mohammedi et al. (n=40 pts) used a constant loading dose (500 mg = 7.5±1.5 mg/kg) and a weight-based approach (15 mg/kg = 1,147±317 mg) [74]. This resulted in different Css of 14.9±5 mg/L and 18.5±6 mg/L. Differences in ICU mortality (50% vs. 35%, p=not significant) and clinical cure (56% vs. 93%, p<0.02) were noticed, arguing for the higher dose and resulting higher C24 (Figure 3a). Spadaro et al. (n=348 pts) studied patients with CrCl above (A) and below (B) 50 mL/min [43]. They reported a significant correlation between subtherapeutic levels at first measurement (C24 target: 15-25 mg/L) and in-hospital mortality (OR 2.1, p=0.003) (Table 3). Additionally, they found lower AUC24/MIC with also numerically lower ICU mortality (M) in group A (A: 468±79, M: 21.4%; B: 490±84, M: 23.9%). Lin et al. (n=52 pts) distinguished between obese (o; BMI > 35 kg/m²) and non-obese (no; BMI < 30 kg/m²) patients [42]. No statistically significant differences were noticed between groups in respect to mean Css and mortality (C24: o/no: 20±4 mg/L; AUC24: o: 488±92 mg*h/L, no: 481±91 mg*h/L; M: o: 19.2%, no: 15.4%). In a subset analysis, Akers et al. (n=90 pts) distinguished between patients with gram-positive bacteremia (1), patients with sepsis without proven gram-positive bacteremia (2) and patients with pneumonia (3) (Cmean: 1: 19±3 mg/L, 2: 21±4 mg/L, 3: 22±4 mg/L) [49]. A slight numerical difference in serum levels was observed, which was not associated with in-hospital mortality (1: 16%, 2: 70%, 3: 35%) and was not studied in relation to the impact on microbiological failure (Cmean overall: 20±4 mg/L; failure overall: 18%). Wysocki et al. (n=61 pts) associated a Cmean of 23±4 mg/L or AUC24 of 596±159 mg*h/L with treatment success and a Cmean of 25±5 mg/L or AUC24 of 685±260 mg*h/L with treatment failure [34]. Higher SCr values were measured in therapy failure, so that an increase in SCr and subsequent increase in Css could be a marker for therapy failure.

### 3.5.2 Safety

Safety was discussed in sixteen studies (n=2,383 pts) (Table 2 or Supplementary Table S5), with “nephrotoxicity” in the form of AKI being dealt with most often (15/16, n=2,331 pts) [33, 34, 42–44, 46–49, 53, 56, 61, 74–76]. Different definitions were used to describe AKI (Table 1); the frequency varied from 0% [42, 47, 74] to 60% [48]. Higher Css (especially >30 mg/L) were identified as a significant predictor of AKI occurrence by multivariate regression analysis (Table 3) [46, 56, 76]. In contrast, Spadaro et al. found no relationship between AKI and Css [43]. In four studies (n=863 pts) vancomycin serum concentration-dependent nephrotoxicity was described, the incidences of which varied widely: Spadaro et al. (n=348 pts) saw no nephrotoxicity at Css of 25-30 mg/L and an incidence <8% (<28/348) when the Css exceeded 30 mg/L [43]. Cianferoni et al. (n=207 pts) described an increasing incidence the higher the Css, with a maximum of 38% (25/66) above a level of 25 mg/L [56]. They also established a link between Css, the onset of AKI and ICU mortality, with rising mortality and AKI rate at higher Css (no AKI: C24 18.7 mg/L, Cmean 21.2 mg/L, M 18%; early AKI: C24 24.5 mg/L, Cmean 27.2 mg/L, M 46%). However, it could not be ruled out that the escalated serum concentrations were only a marker for a declining glomerular filtration and did not cause AKI per se. Perin et al. (n=179 pts) noted an AKI rate of 55% (72/131) with Css <25 mg/L versus 77% (34/44) with Css >25 mg/L [48]. Spapen et al. (n=129 pts) demonstrated an increase in AKI the higher the Css: <25 mg/L: 4.5% (3/68), 25-30 mg/L: 31% (9/29), >30 mg/L: 81% (26/32) [46]. Additionally, mortality was higher in patients with AKI (no AKI: 20%, AKI: 53%, p=0.01). Comparing the information from the latter three studies (the only ones that provided patient numbers with concentration-dependent AKI rates), there appeared to be an advantage in terms of AKI occurrence when the target concentration was <25 mg/L (Figure 5, Supplementary Table S6 and Table S7).

## 4 Discussion

Although vancomycin has been used for decades to treat infections with gram-positive pathogens and in numerous studies data on vancomycin concentration and therapeutic outcome was published, this systematic review shows that there is few data on the target serum concentration range to achieve effective yet tolerable therapy during CI in critically ill adult patients.

### 4.1 Target attainment

The literature did not provide associations between TA and Css; achievement of target ranges was used to evaluate new dosing protocols [42, 53, 55] or different dosing modalities [24, 25]. Based on differences in tissue penetration of vancomycin [81] or pathogen susceptibility [82], an indication-dependent target range selection would be likely. For example, Tsutsuura et al. showed that in MRSA bacteraemia, but not MRSA infections per se, higher trough concentrations resulted in significantly less treatment failures [83]. However, no correlation between the selected target range and the investigated indication was apparent to us in the synopsis. When plotting the TA rate and Css of each trial, higher TA was observed with higher LD without influence of MD (Figure 2a/b). In studies with sepsis or burn patients, lower TA rates were found. Physiological changes during sepsis or burns may have played a role (e.g. capillary leakage or oedema) [13–15]. However, it should be noted that different definitions of sepsis were used in the studies, making it difficult to compare patients and sepsis rates. Temporal differences (16-48 h) to reach target Css could be due to another dosing regimen (higher vancomycin doses equal faster TA) [33, 34, 59].

### 4.2 Efficacy

An assessment of the reported mortality data was difficult. Mortality seemed to depend on numerous factors and was not only influenced by Css. Furthermore, there was very little comparative concentration and mortality data. Higher C24 resulted in lower mortality rates, with a concentration above 15 mg/L found to be favorable [43, 74]. A target concentration for Cmean or AUC24 leading to lower mortality was not evaluated. Looking at the available datasets of each study’s average Css and mortality rate, we also noticed an association of ICU mortality with C24 (R²=0.7435), but not with Cmean (R²=0.0057) or AUC24 (R²=0.0009) (Figure 3a-c). In a consensus review published in 2020 by several societies on the dosing and monitoring of vancomycin, for CI a lower limit of the target Css range of 20 mg/L (=AUC24/MIC ≥480 if MIC≤1 mg/L; PK/PD target not validated) was recommended [84]. This threshold was pharmacokinetically, microbiologically and clinically justified [34], was used in most studies considered [84], but was not derived from concentration comparative effectiveness studies. For II, the same practice guideline and meta-analyses recommended AUC_total_/MIC-guided monitoring with a value ≥400 (if MIC≤1 mg/L, determined by broth microdilution (BMD)) as the PK/PD target for efficacy [83–85]. For CI, this target would correspond to a concentration of 17 mg/L (if MIC=1 mg/L) and is thus close to the lower limit of the target concentration range found in our research. Cristallini et al. calculated AUC24/MIC ratios ≥400 (if MIC≤1 mg/L) for C24 ≥15 mg/L [52]. The PK/PD threshold resulted from studies with infections with MRSA whose ECOFF of ≤2 mg/L is half that of MRCoNS or E faecalis [82, 84]. Studies on the optimal vancomycin PK/PD target for infections with these germs are lacking. However, Ampe et al. calculated AUC_total_/MIC_BMD_=667, AUC_free_/MIC_BMD_=452, AUC_total_/MIC_Etest_=457 and AUC_free_/MIC_Etest_=301 as the thresholds between clinical success or failure in ward patients with mono-infections of various gram-positive pathogens and CI as the only effective agent [86]. A transfer of dose recommendations from II to CI is uncertain, though. When comparing mortality during CI and II, meta-analyses have found no difference [22–25]. Nevertheless, the average measured Css in the included studies were always higher for CI and additionally differed within the comparison groups. A meta-analysis on mortality of the same Css of CI vs. II is missing so far. Additionally, for II Dalton et al. demonstrated that the use of AUC/MIC to predict patient outcome was modest [87], and Tsutsuura et al. found no significant difference in mortality rates with trough-versus AUC-guided treatment monitoring [83]. For CI, the benefit of PK/PD-guided therapy remains completely unclear. Mohammedi et al. emphasized the need for sufficient LD to maintain high C24 and reduce mortality [74]. We observed the importance of sufficient LD as well (Supplementary Table S3). Additionally, higher MD seemed to result in lower mortality in the C24 cohort (Supplementary Table S3). Reaching high therapeutic levels as early as possible at the beginning of anti-infective therapy is in accordance with general recommendations [15, 88–93]. CI has advantages over II in this respect [26–29, 32–34]. However, sufficient dosing in critically ill patients is challenging [15], partly due to the increased volume of distribution with impaired capillary barrier function and the probable losses through renal function and replacement procedures [14]. The difficulty of appropriate dosing could account for the heterogeneity of doses applied in the studies included in this review. From our diagrams (Figure 3a-c), it could be deducted that dialysis was a predictor of mortality, supported by the known higher mortality of dialysis patients [94]. The initial severity of illness, as measured by APACHE II, SAPS II or SOFA score, may also have influenced mortality (Supplementary Table S2). Css and clinical cure in two CI cohorts were only compared by Mohammedi et al. (higher C24 equals higher cure) [74], whereas Css and microbiological efficacy were not analysed comparatively in any study. For Cmean and clinical success, the comparison of results by Stepan et al. and Wysocki et al. coincided with the analysis of Mohammedi et al. (higher Cmean equals higher cure), but not for microbiological success (higher Cmean equals lower cure) (Supplementary Table S10) [34, 47, 74]. This may be related to different observation periods (five days vs. end of treatment). With II, higher trough levels or AUC24/MIC values resulted in better clinical *and* microbiological cure [95–97].

### 4.3 Safety

The nephrotoxicity of vancomycin is known [98], with CI being significantly associated with a lower risk compared to II [22–25]. However, definitions of reported AKI varied. As Hutschala et al. and Koeze et al. showed, this had implications for the reported incidence, timing and outcome of AKI [33, 99]. Newer definitions such as AKIN (Acute Kidney Injury Network) [100] or KDIGO (Kidney Disease Improving Global Outcomes) [101] are more sensitive, resulting in higher reported rates of AKI. In our comparison, we related only similar AKI definitions. Due to poor tissue penetration, [81] the therapy with vancomycin is limited as the dose cannot be increased arbitrarily. The upper limit of Css that keeps the impairment of renal function within acceptable limits, weighing the benefits and harms, is much debated. In dataset plots of Css against AKI rate, we found that higher Css and longer duration of therapy increased the rate of AKI (Figure 4), as also calculated by Cianferoni et al. and Hanrahan et al. with multiple regression (Table 3) [56, 76]. For II, an upper trough level limit of 20 mg/L was established and AUC24/MIC ≤600 (if MIC≤1 mg/L, determined by BMD) was set as PK/PD target for safety [83, 84]. For CI, this target corresponds to a Css of 25 mg/L [84] and coincides with the preferred concentration we calculated. However, due to a lack of data, we could not compare 25 mg/L with other thresholds such as the previously described upper limits of 28 mg/L or 30 mg/L [46, 102]. Elevated AUC24 levels have also been reported to increase the risk of AKI in CI [37, 83]. AKI per se (vancomycin-independent) has been associated with worse treatment outcome (e.g. mortality, long-term impaired renal function) [20] and longer ICU and hospital stays, as well as higher costs for the healthcare system [103–105]. Cianferoni et al. and Spapen et al. also noted prolonged deterioration in renal function after AKI, and they and Omuro et al. described increased mortality in patients who developed AKI with CI [46, 56, 61]. AKI with CI has negative consequences and should consequently be avoided.

### 4.4 Limitations

Several limitation should be considered when interpreting the results. First, of the twenty-one included studies, only two were randomized controlled trials; most were retrospective or observational studies. Because of their observational design, allocation bias, selection bias and various types of other confounding factors may influence the results of our report. Publication bias is to be expected, since publications that demonstrate an effect are more likely to be published. Second, no raw data was available. Instead, our plotting of datasets was performed with means and medians. Thus, the compilation of a meta-analysis or the calculation of cut-off values for efficacy was not possible. Third, our forest plot included data only from the studies that provided the number of patients with concentration-dependent AKI. Fourth, to distinguish vancomycin-induced nephrotoxicity from the naturally high rate of AKI in ICU patients, a comparison group would have been necessary in all studies. Albeit, only Omuro et al. used a control group, but the included patients were randomly selected and no matching was done [61]. Fifth, in accordance with clinical TDM routine, only the total amount of vancomycin in serum (bound + free) was measured in the studies, although it is rather the drug not bound to plasma proteins that is active [106, 107]. Variations in protein binding of vancomycin (<10-82%) [107–110] and albumin concentration of critically ill patients [15] have been described. Thus, the active vancomycin concentration varies greatly. Berthoin et al. reported poor correlation between total and free vancomycin concentration (R²=0.55) [111] and concluded that to reduce the treatment failure rate in infection by less susceptible organisms, the free concentration should rather be determined. The procedure for sensitive germs remains unclear. Since the free concentration cannot be determined in most laboratories, it should be investigated whether the individual protein concentration is a useful surrogate parameter for free vancomycin. Finally, only few AUC24 and only one AUC24/MIC value were available. Rybak et al. argue for sufficiency of AUC24 determination because the MIC distribution is narrow at≤1 mg/L, measurement is not very accurate or values are not readily available, and test methods vary widely [84]. Nevertheless, knowledge of MIC values and inclusion of these in therapy assessment is important, since higher therapy failure has been described for MIC >1 mg/L and higher necessary dosage would increase nephrotoxicity [86, 112].

## 5 Conclusions

Despite currently sparse data availability, it appeared that for continuous infusion of vancomycin (CI) mortality was reduced and clinical cure increased with C24 above 15 mg/L and AKI may be reduced with Css below 25 mg/L. The range of 15-25 mg/L to aim for in CI needs to be validated by direct comparison with other concentration ranges, just as the definition of specific AUC(/MIC) or indication-dependent target Css need to be further investigated to achieve a safe (i.e. least damaging to the kidneys) and simultaneously most effective (i.e. therapeutically successful) therapy. To this end, future research should always sort patients by vancomycin serum concentration groups. Large prospective controlled studies are needed for this purpose.

## Data Availability

Data is contained within the article.

## 6 Declarations

## Acknowledgments

We acknowledge Viola Fuchs for her support in supervising the research project and we thank Daniel Mai for providing the R Statistical Software, Alexander and Renate Viertel for assistance with data analysis and Andreas Ismair for basic help in the writing process of the manuscript.

## Ethical Approval

Not required.

## Author contributions

K.V. and F.M. conceived of the presented idea. K.V. conducted the study selection and data extraction. M.S. extracted data from a sample of eligible studies, good agreement was achieved with the data extracted by K.V.. K.V. and F.M. analysed the data. K.V. wrote the manuscript with support and critical revision from F.M., T.A., E.F. and M.S.. F.M. supervised the project. All authors reviewed and contributed to the final manuscript. All authors have read and agreed to the published version of the manuscript.

## Declaration of Generative AI and AI-assisted technologies in the writing process

During the preparation of this work the author(s) used the free internet translational programme Deepl (www.Deepl.com/Translator) in order to improve the linguistic quality of the manuscript. After using this tool/service, the author(s) reviewed and edited the content as needed and take(s) full responsibility for the content of the publication.

## 8 Tables and Figures

### 8.1 Definition of nephrotoxicity

**Table 1:**
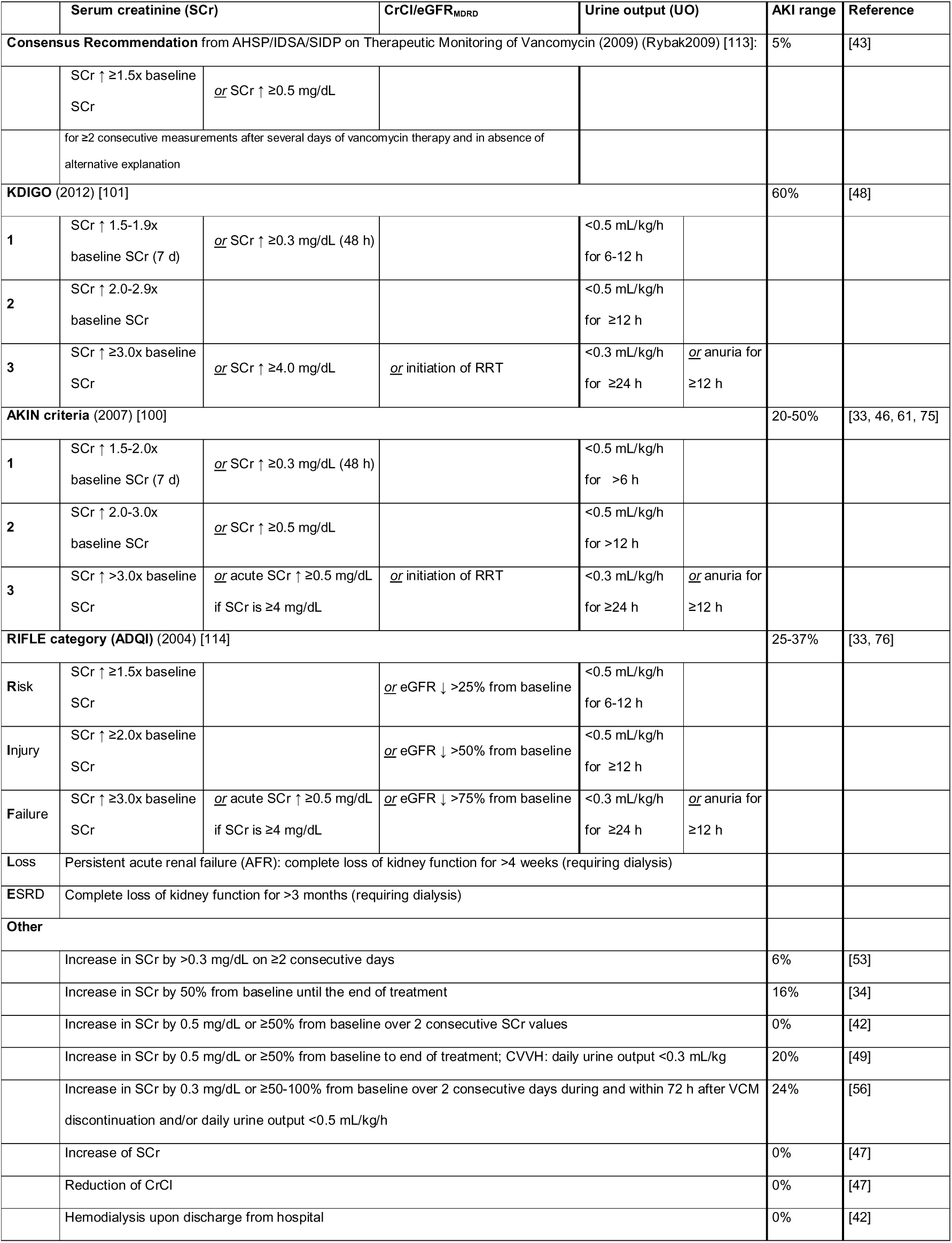
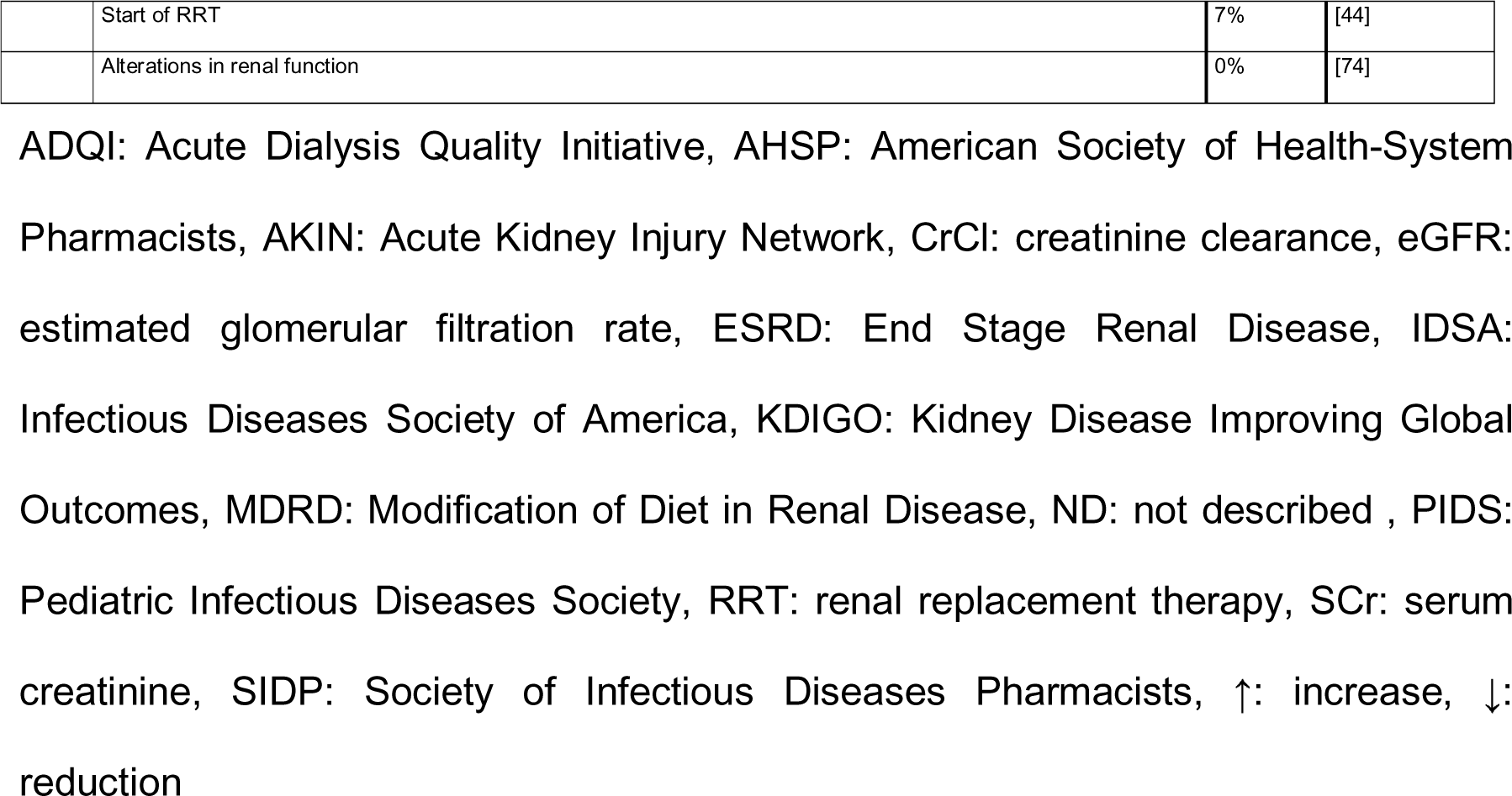
Compilation of all definitions of acute renal insufficiency (AKI) used in the included publications.

### 8.2 Flowchart

**Figure 1:**
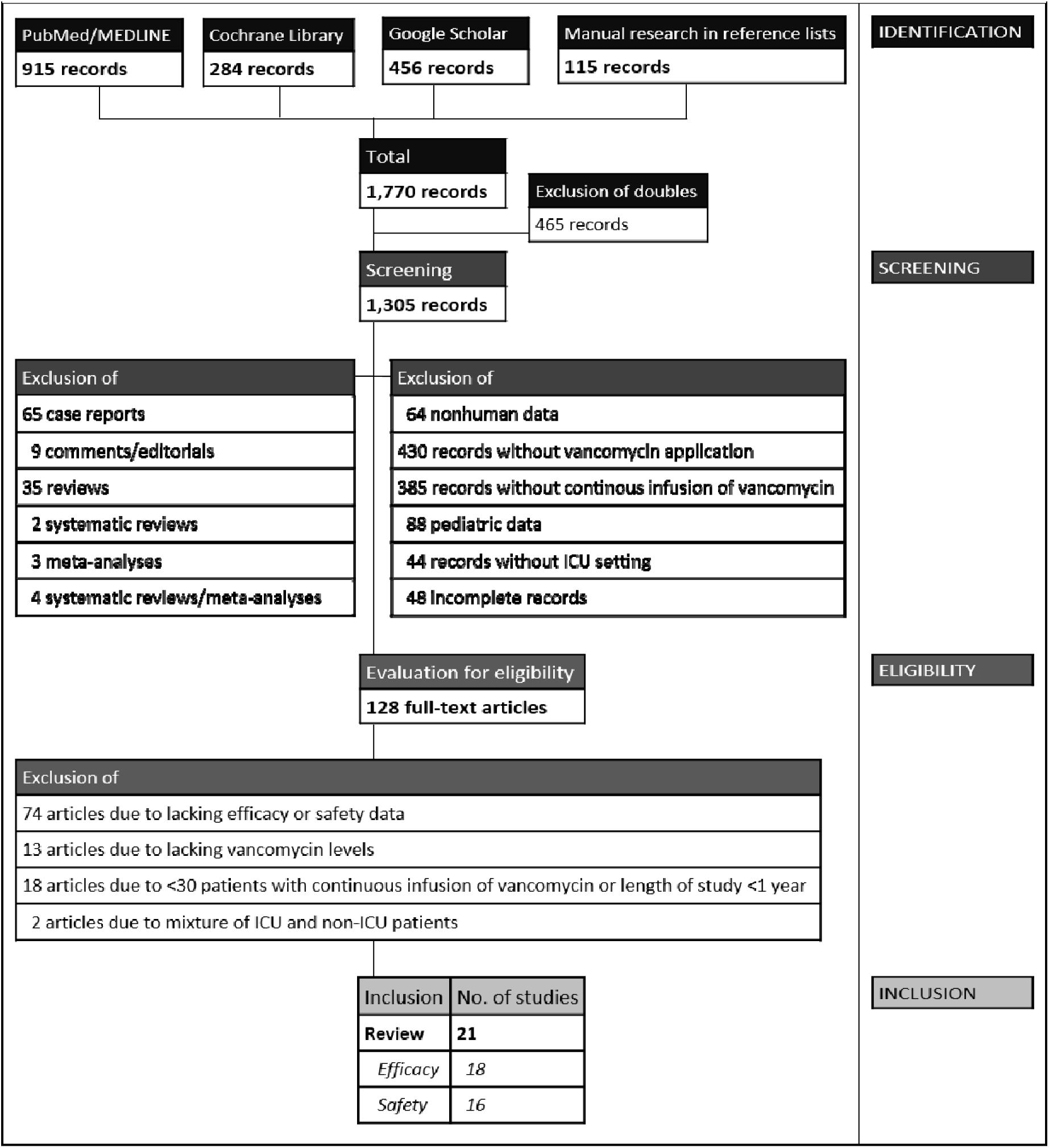
Flowchart of the study selection.

### 8.3 Overview

**Table 2:**
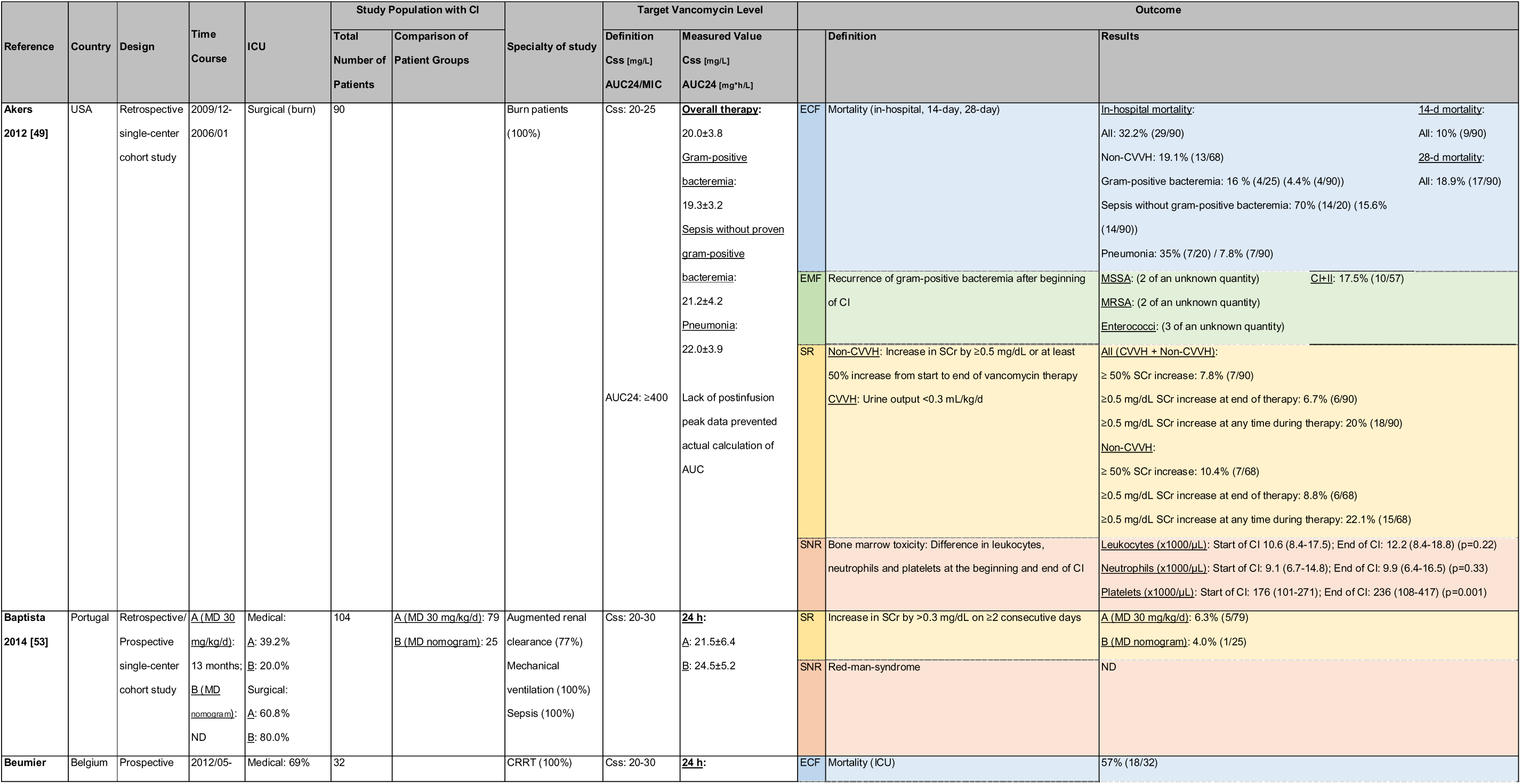
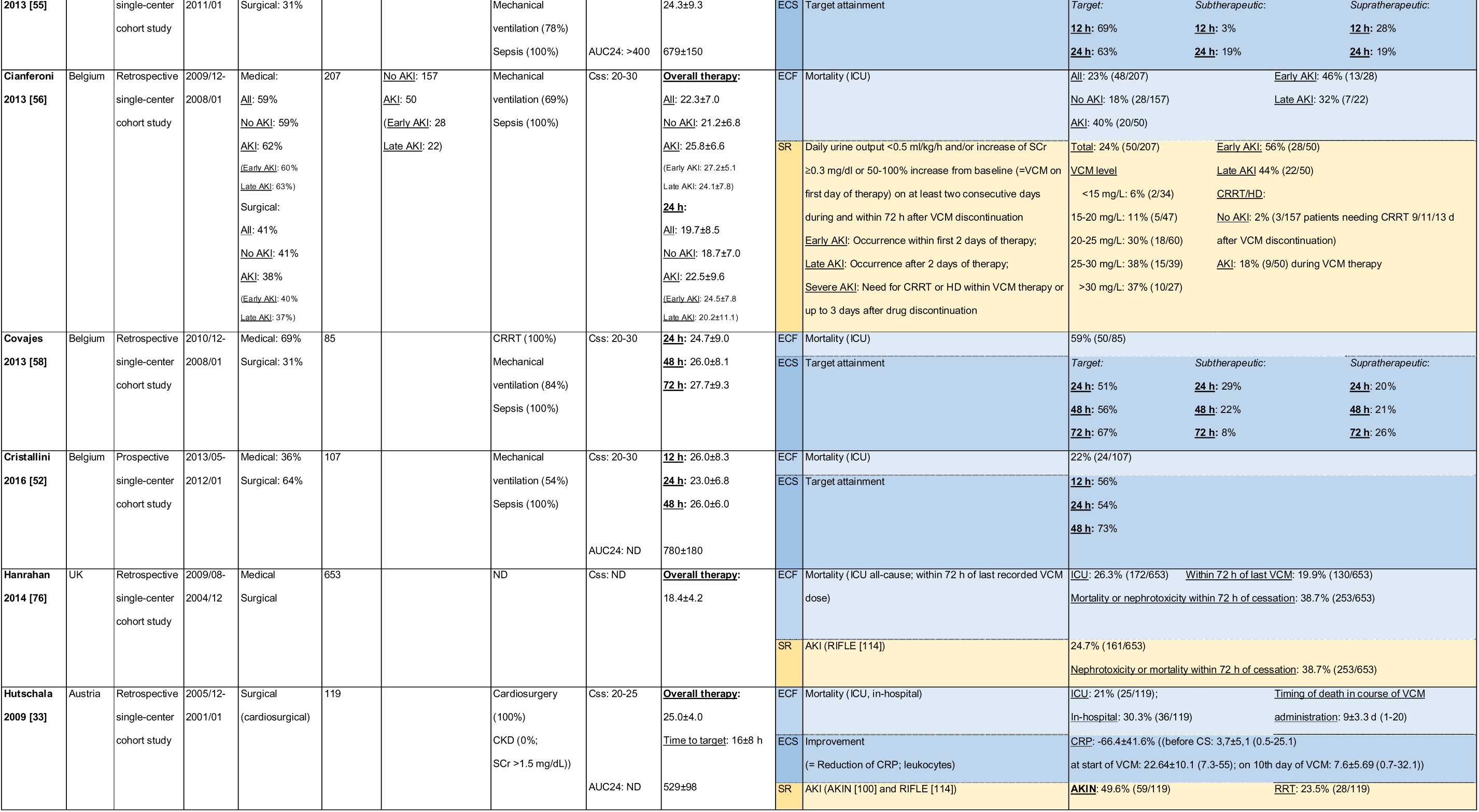
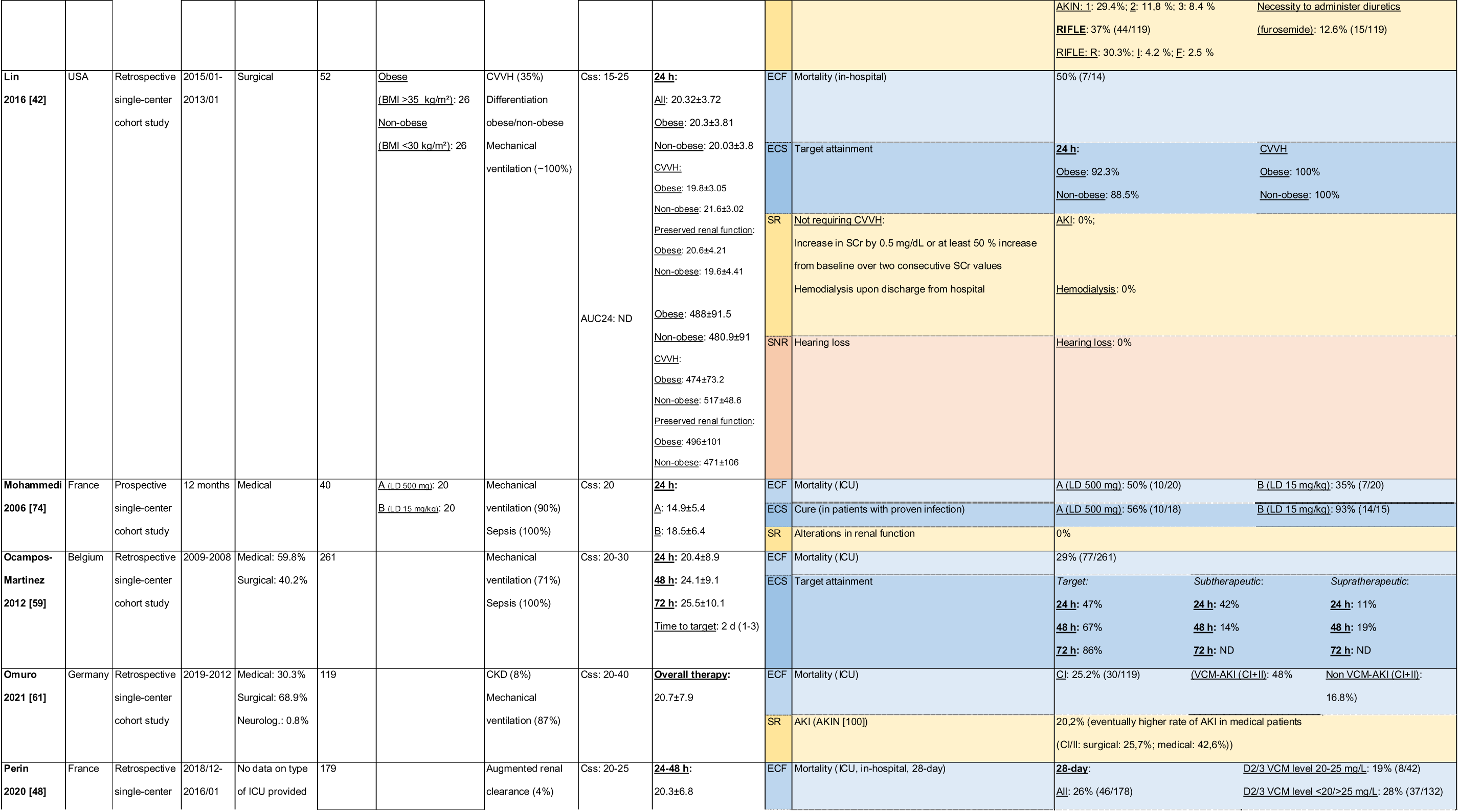
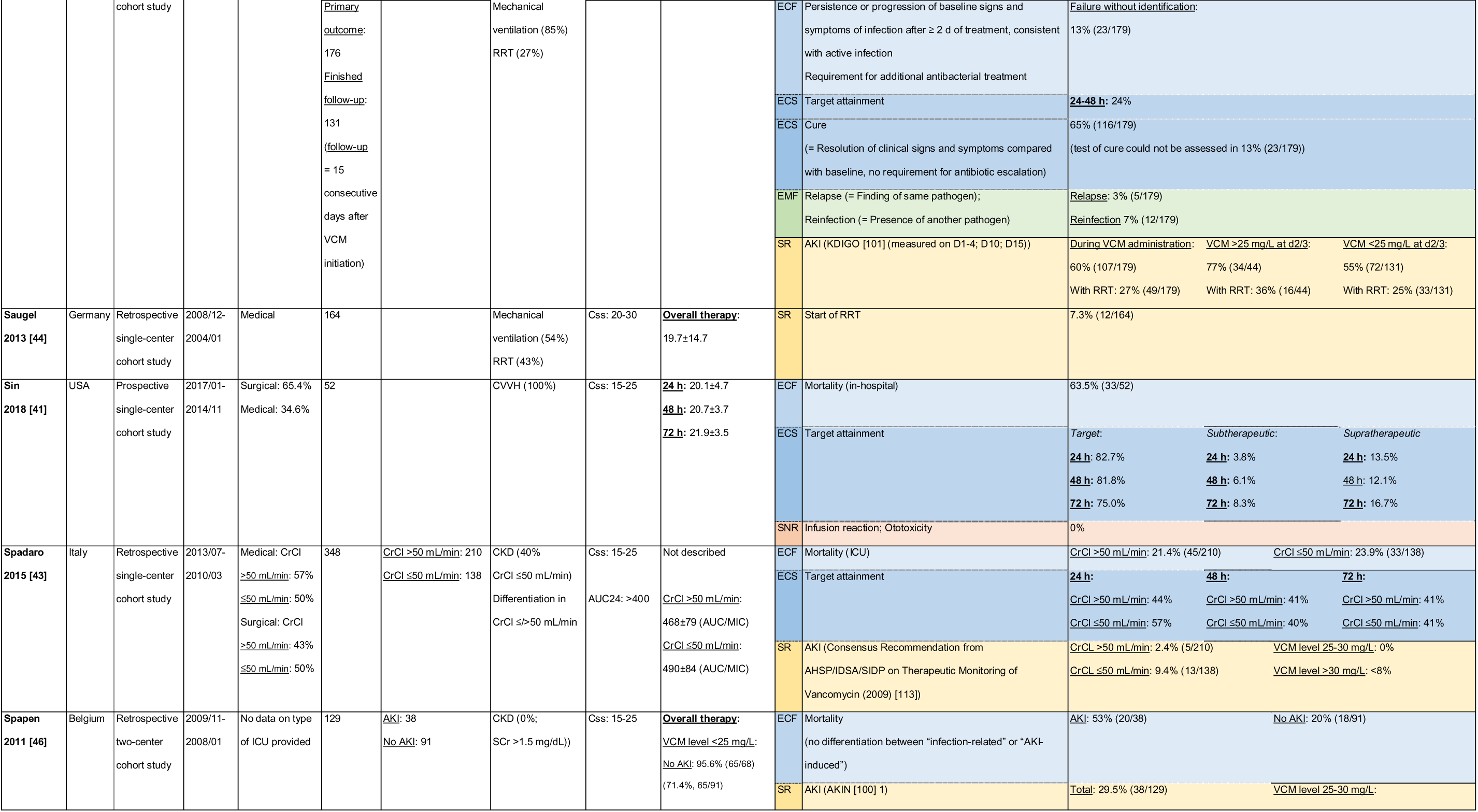
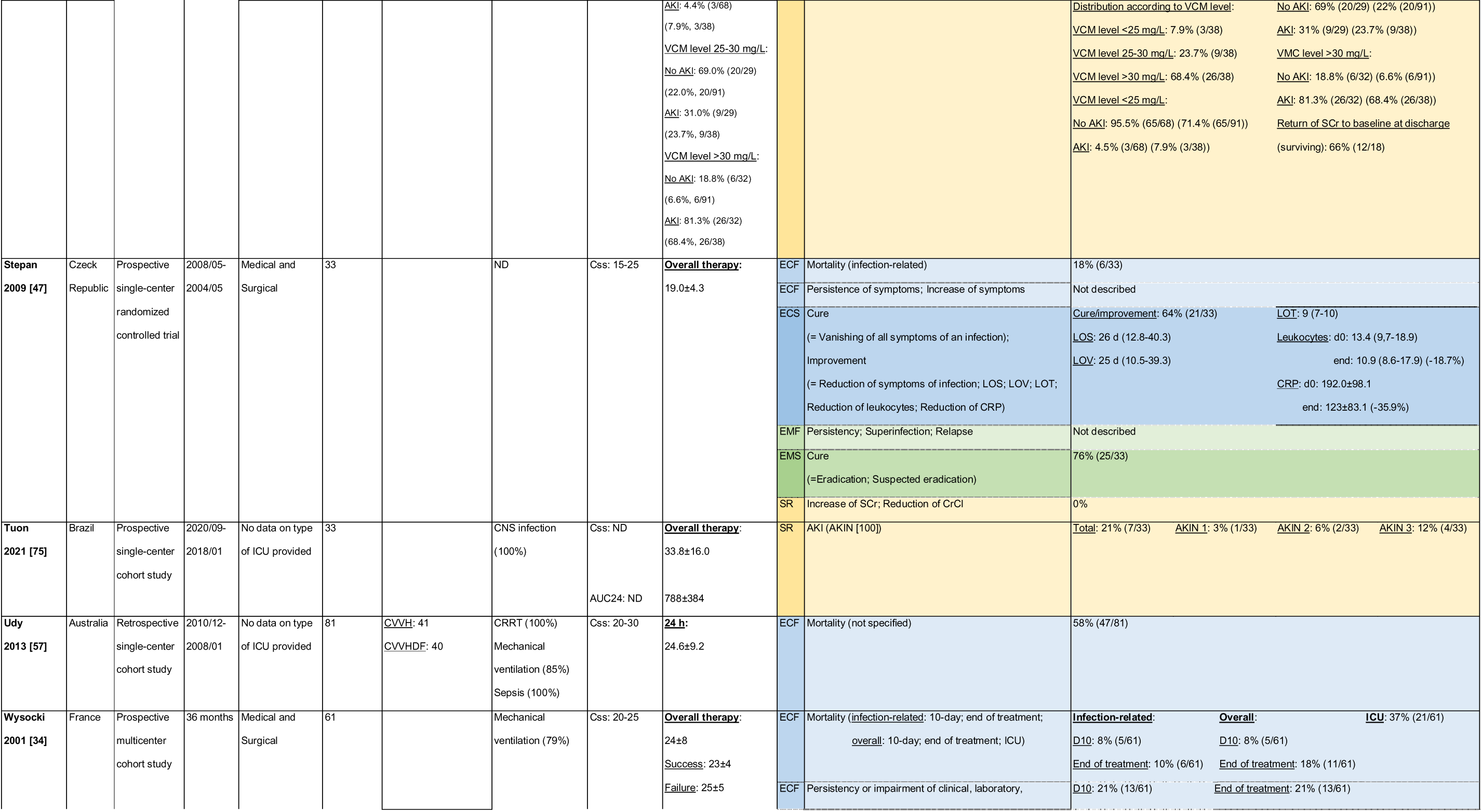
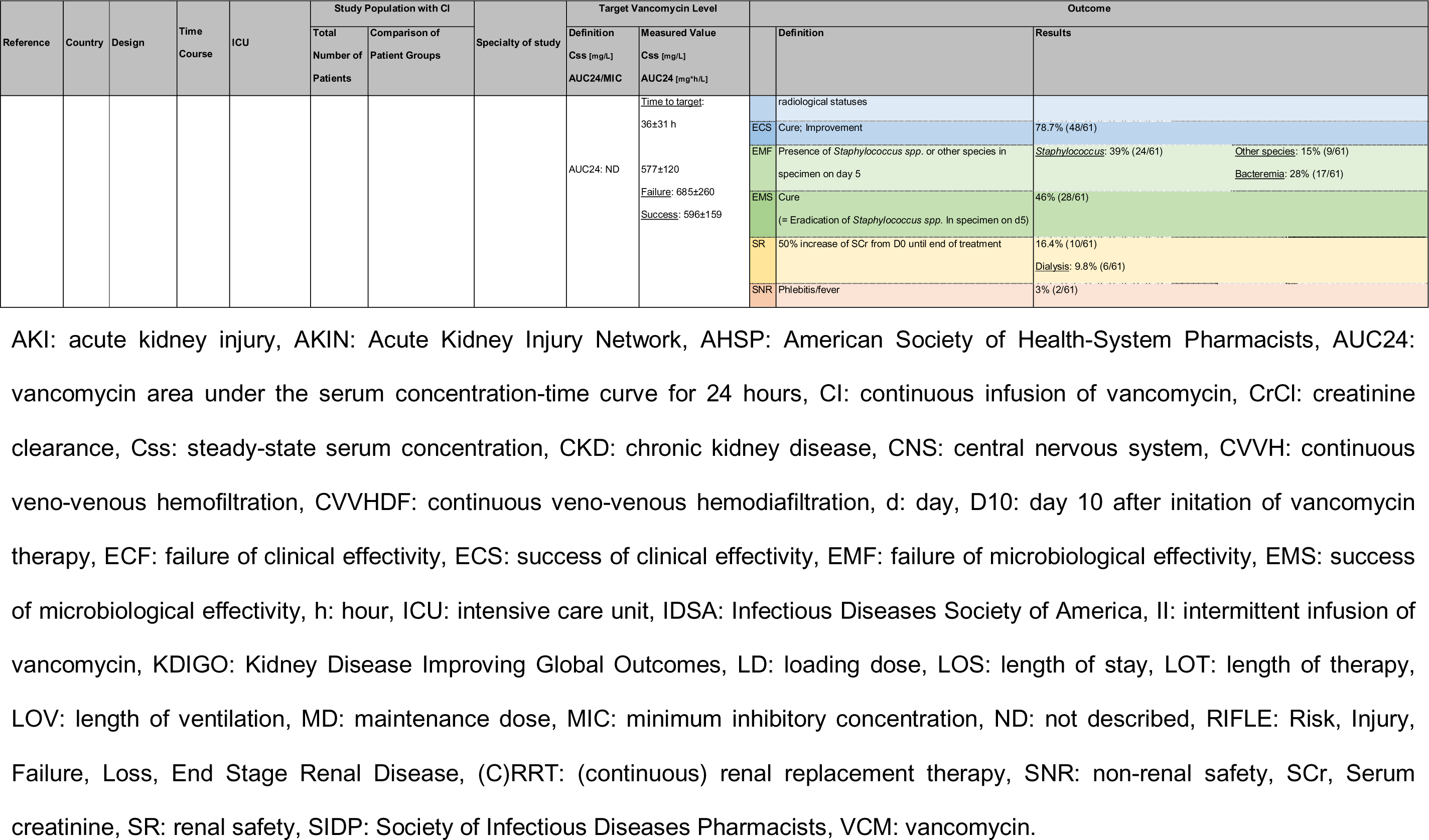
Overview of the main characteristics of the twenty-one included studies with continuous infusion of vancomycin.

### 8.4 Significant predictors of outcomes

**Table 3:** Significant predictors for the outcomes “acute kidney injury”, “mortality” and “target attainment” identified by univariate and multivariate regression analysis in the studies.

| Univariate | Multivariate |
| --- | --- |
| *Target attainment (TA)*<br>• Loading dose (↑ dose → ↑ TA) [59]<br>• Daily vancomycin dosage (↑ dosage → ↑ TA) [58]<br>• Serum creatinine (↑ SCr → ↑ TA) [59]<br>• Creatinine clearance (↓ CrCl → ↑ TA) [59]<br>• CRRT intensity (↓ intensity → ↑ TA) [55, 58]<br>• Age (↓ age → ↑ TA) [58]<br>• Body weight (↑ body weight → ↑ TA) [52, 55]<br>• Gender (male → ↓ TA) [59]<br>• Prolonged ICU stay before initiation of vancomycin (↑ stay → ↓ TA) [48]<br>• SOFA score (↑ score → ↑ TA) [59]<br>*ICU mortality*<br>• AKI (AKI → ↑ mortality) [61]<br>*Acute kidney injury (AKI)*<br>• Diabetes mellitus (diabetes → ↑ AKI) [46]<br>• Shock (shock → ↑ AKI) [46] | *Target attainment (TA)*<br>• Daily vancomycin dosage (↑ dosage → ↑ TA) [58]<br>• Creatinine clearance (↓ CrCl → ↑ TA) [52, 59]<br>• CRRT intensity (↓ intensity → ↑ TA) [58]<br>• Age (↓ age → ↑ TA) [58]<br>• Body weight (↑ body weight → ↑ TA) [52]<br>• Body mass index (↑ BMI → ↑ TA) [52]<br>• Gender (male → ↓ TA) [59]<br>• Prolonged ICU stay before initiation of vancomycin (↑ stay → ↓ TA) [48]<br>*In-hospital mortality*<br>• C24 <15 mg/L (↓ C24 → ↑ mortality) [43]<br>• SAPS II score (↑ score → ↑ mortality) [43]<br>*Acute kidney injury (AKI)*<br>• Css of CI (esp. >30 mg/L) (↑ Css → ↑ AKI) [46, 56, 76]<br>• Duration of therapy (at time of highest SCr (risk/injury)) (↑ duration → ↑ AKI) [56, 76] |
|  | CrCl at ICU admission [43]<br>Body weight (↑ lean body weight → ↑ AKI) [46]<br>SAPS 3 score (↑ score → ↑ AKI) [46]<br>*Early AKI*<br>Bacteremia (bacteremia → ↑ AKI) [56]<br>Cmean day 1-3 (↑ Cmean → ↑ AKI) [56]<br>Daily vancomycin dosage (day 1-3) [56]<br>(↓ dosage → ↑ AKI)<br>*Late AKI* [56]<br>Diabetes (diabetes → ↑ AKI) [56]<br>Duration of therapy (↑ duration → ↑ AKI) [56] |

AKI: acute kidney injury, BMI: body mass index, C24: mean steady-state vancomycin serum concentration about 24 hours after initiation of therapy, Cmean: mean steady- state vancomycin serum concentration during the entire duration of therapy or at least three days of therapy, CI: continuous infusion of vancomycin, CrCl: creatinine clearance, CRRT: continuous renal replacement therapy, Css: steady-state serum concentration, ICU: intensive care unit, SAPS: Simplified Acute Physiology Score, SCr: serum creatinine, SOFA score: Sepsis-related Organ Failure Assessment score, TA: target attainment, ↑: increase, ↓: reduction

### 8.5 Target attainment

**Figure 2:**
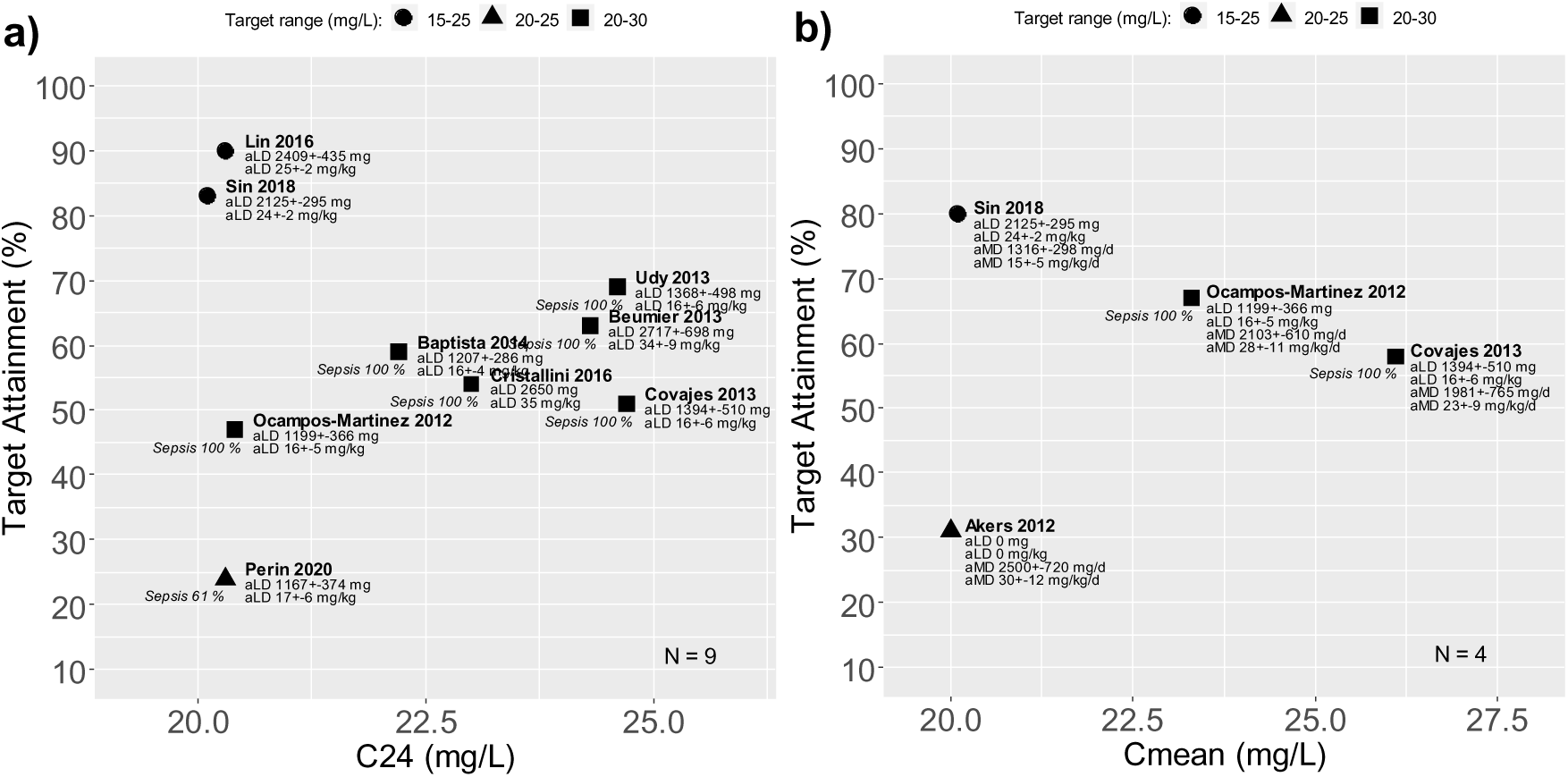
Scatterplots of mean vancomycin steady-state serum concentrations (Css) plotted against each cohort’s rate of target attainment. **a)** Studies in which Css approximately 24 h after initiation of therapy with continuous infusion of vancomycin (C24) were reported (n=9 studies representing 953 patients). **b)** Studies in which Css during the entire duration of therapy with continuous infusion of vancomycin or at least three days of therapy (Cmean) were reported (n=4 studies representing 488 Filled square: Css target range of 20-30 mg/L, filled circle: Css target range of 15-25 mg/L, filled triangle: Css target range of 20-25 mg/L, aLD: applied average loading dose, aMD: applied average maintenance dose, Sepsis: proportion of patients with sepsis in the study population

### 8.6 Mortality

**Figure 3:**
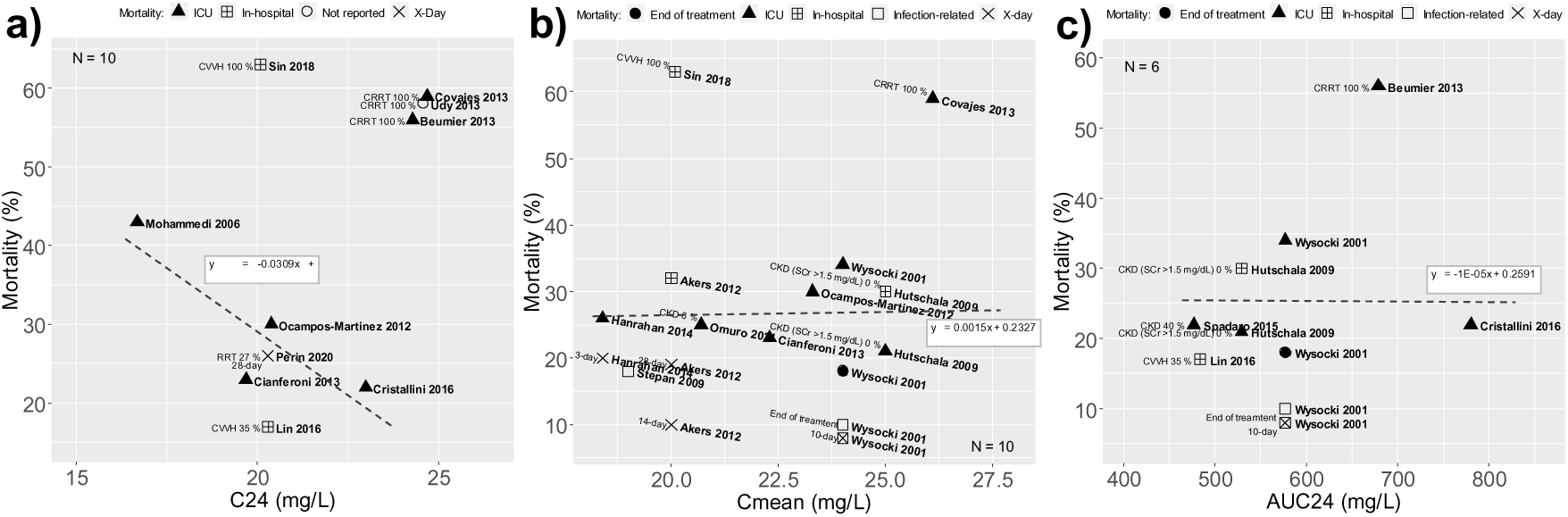
Scatterplots of mean vancomycin steady-state serum concentrations (Css) plotted against each cohort’s mortality rate. The dashed lines represent the regression lines (RL) of ICU mortality (▴ filled black triangle) in studies without 100% dialysis patients. **a)** Studies in which Css approximately 24 h after initiation of therapy with continuous infusion of vancomycin (C24) were reported (n=10). RL: n=4 studies representing 615 patients; R²=0.7435. **b)** Studies in which Css during the entire duration of therapy with continuous infusion of vancomycin or at least three days of therapy (Cmean) were reported (n=10). RL: n=6 studies representing 1,420 patients; R²=0.0057. **c)** Studies in which mean vancomycin areas under the serum concentration-time curve for 24 h for continuously administered vancomycin (AUC24) were reported (n=6). RL: n=4 studies representing 635 patients; Filled black circle: mortality at the end of treatment with vancomycin, filled black triangle: ICU mortality, bordered cross: in-hospital mortality, X: x-day mortality (e.g. 10-day, 28-day, 30-day), non-filled circle: mode of mortality not reported, non-filled square: infection-related mortality. CKD: chronic kidney disease, (C)RRT: (continuous) renal replacement therapy, CVVH: continuous veno-venous hemofiltration, ICU: intensive care unit, SCr: serum creatinine.

### 8.7 Nephrotoxicity

**Figure 4:**
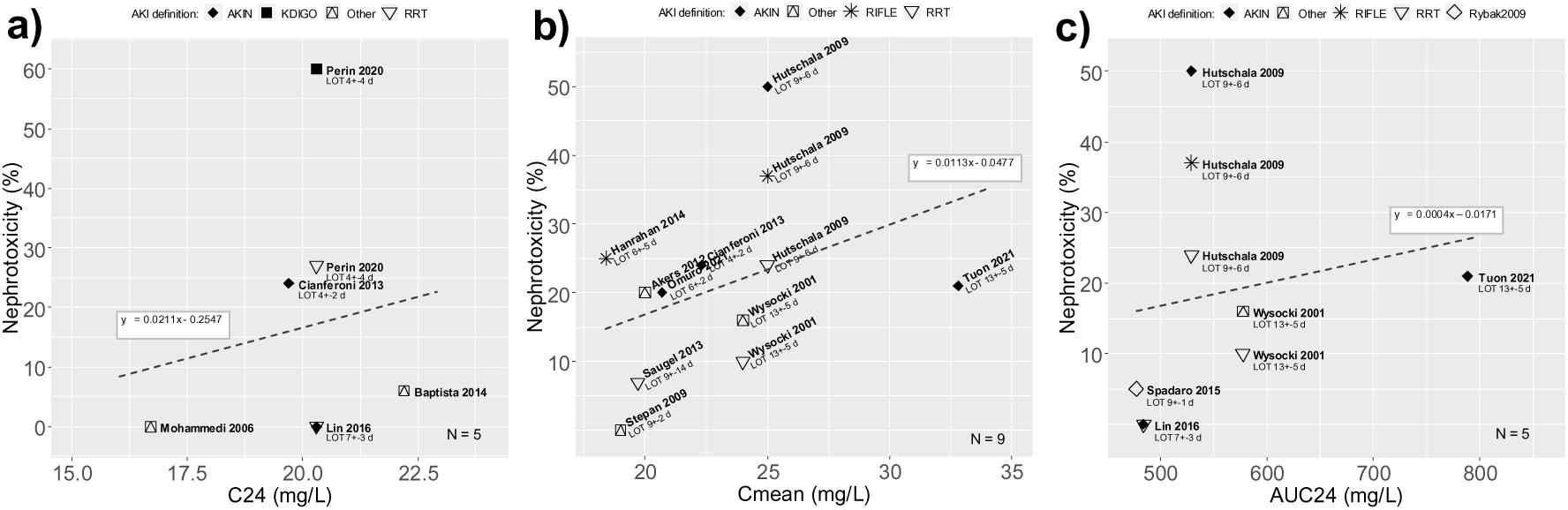
Scatterplots of mean vancomycin steady-state serum concentrations (Css) plotted against each cohort’s rate of acute kidney injury (AKI) (= Nephrotoxicity). The dashed lines represent the regression lines calculated from the data of all cohorts of a respective Css class. **a)** Studies in which Css approximately 24 h after initiation of therapy with continuous infusion of vancomycin (C24) were reported (n=5 studies representing 582 patients; R²=0.0242). **b)** Studies in which Css during the entire duration of therapy with continuous infusion of vancomycin or at least three days of therapy (Cmean) were reported (n=9 studies representing 1,479 patients; R²=0.1174). **c)** Studies in which mean vancomycin areas under the serum concentration-time curve for 24 h for continuously administered vancomycin (AUC24) were reported (n=5 studies representing 613 patients; R²=0.0411). Filled black diamond: AKIN, filled black square: KDIGO, squared triangle: other definition of AKI, star: RIFLE, non-filled triangle: RRT, non-filled diamond: Rybak2009 AKIN: Acute Kidney Injury Network, KDIGO: Kidney Disease Improving Global Outcomes, LOT: length of therapy with continuously administered vancomycin, RIFLE: Risk, Injury, Failure, Loss, End Stage Renal Disease, RRT: renal replacement therapy, Rybak2009: Consensus Recommendation from AHSP/IDSA/SIDP on Therapeutic Monitoring of Vancomycin (2009) (AHSP: American Society of Health-System Pharmacists, IDSA: Infectious Diseases Society of America, SIDP: Society of Infectious Diseases Pharmacists).

**Figure 5:**
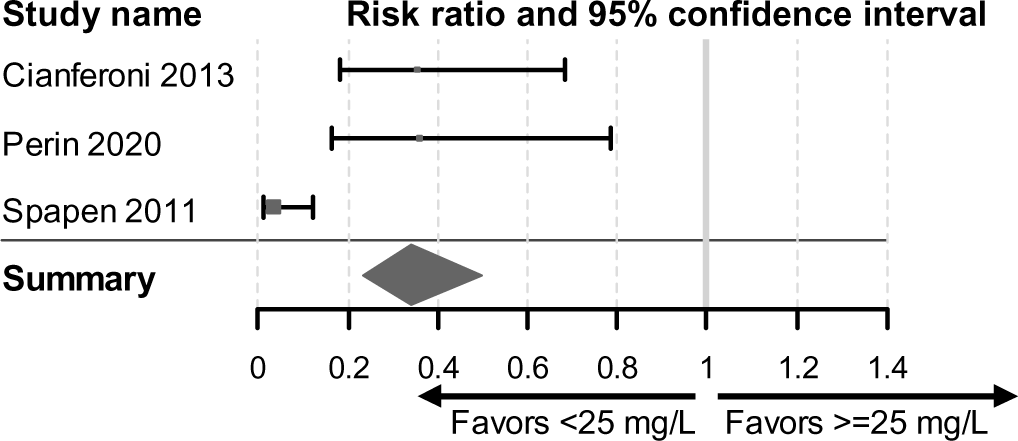
Forest plot comparing the influence of vancomycin steady-state serum concentration below or above 25 mg/L on acute kidney injury (AKI) (n=3 studies representing 515 patients). Overall risk ratio = 0.535 (95% confidence interval 0.432-0.662; z = −5.745; p = <0.0001). Risk ratio <1 indicates a lower risk of acute kidney injury at a vancomycin serum concentration of <25 mg/L as opposed to ≥25 mg/L. The dark grey diamond represents the overall risk ratio.

## 9 Supplementary data

### 9.1 Study Design

**Table S1:**
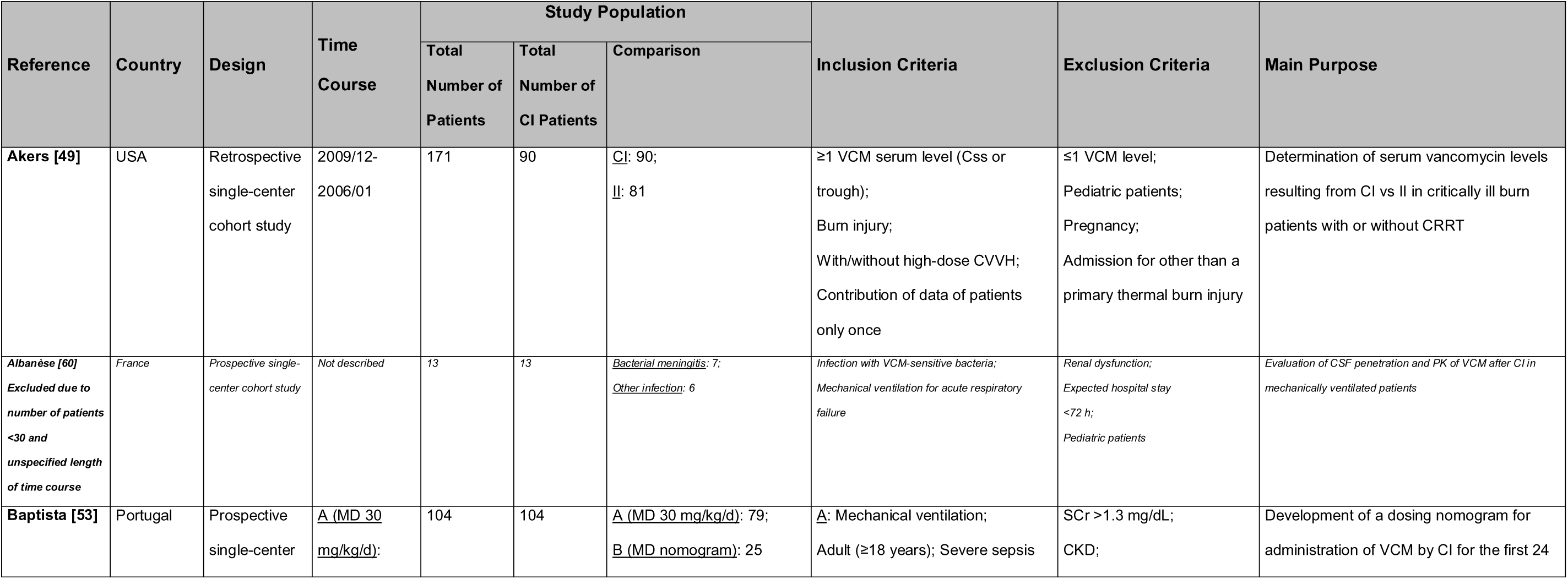
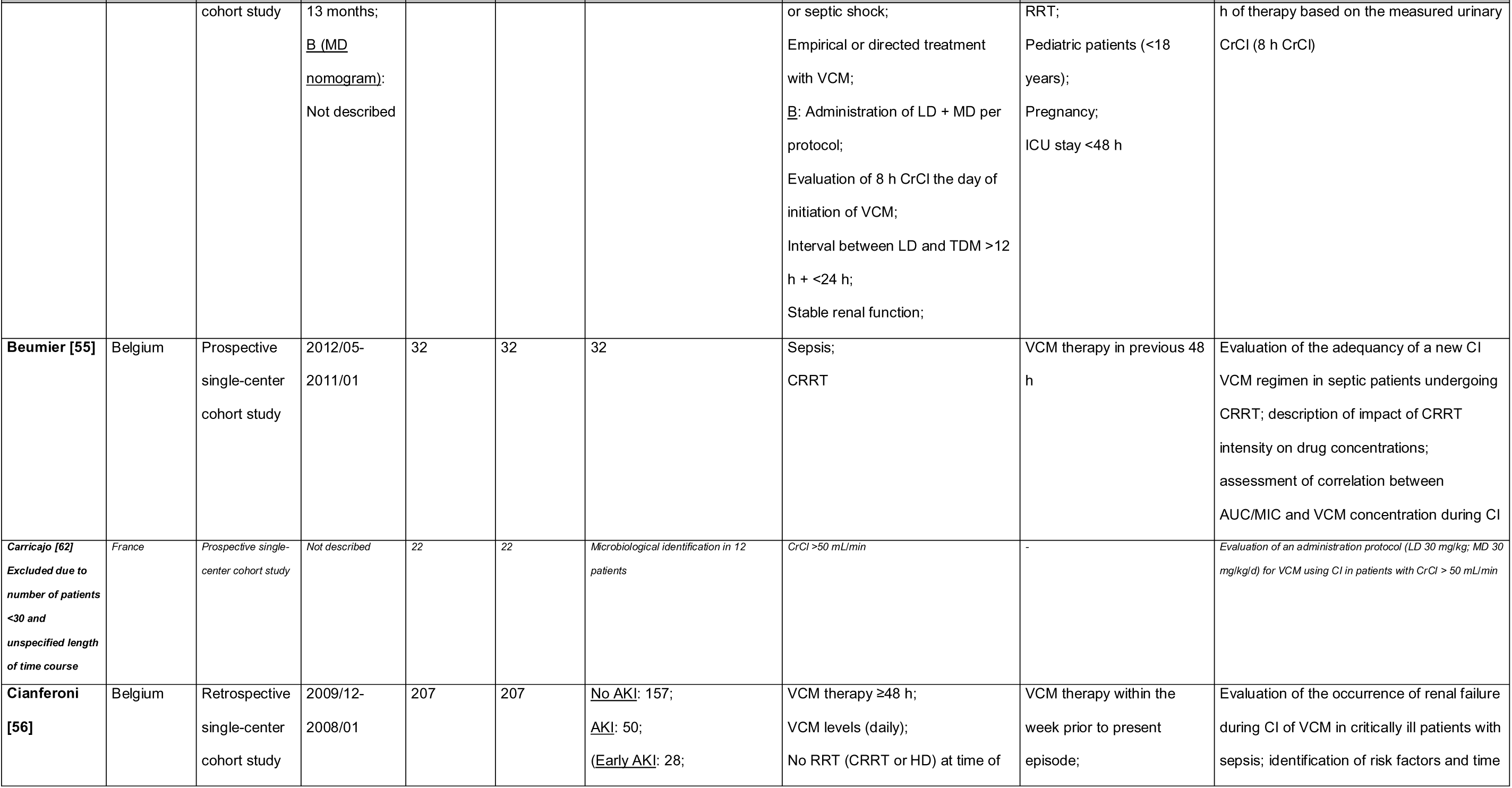
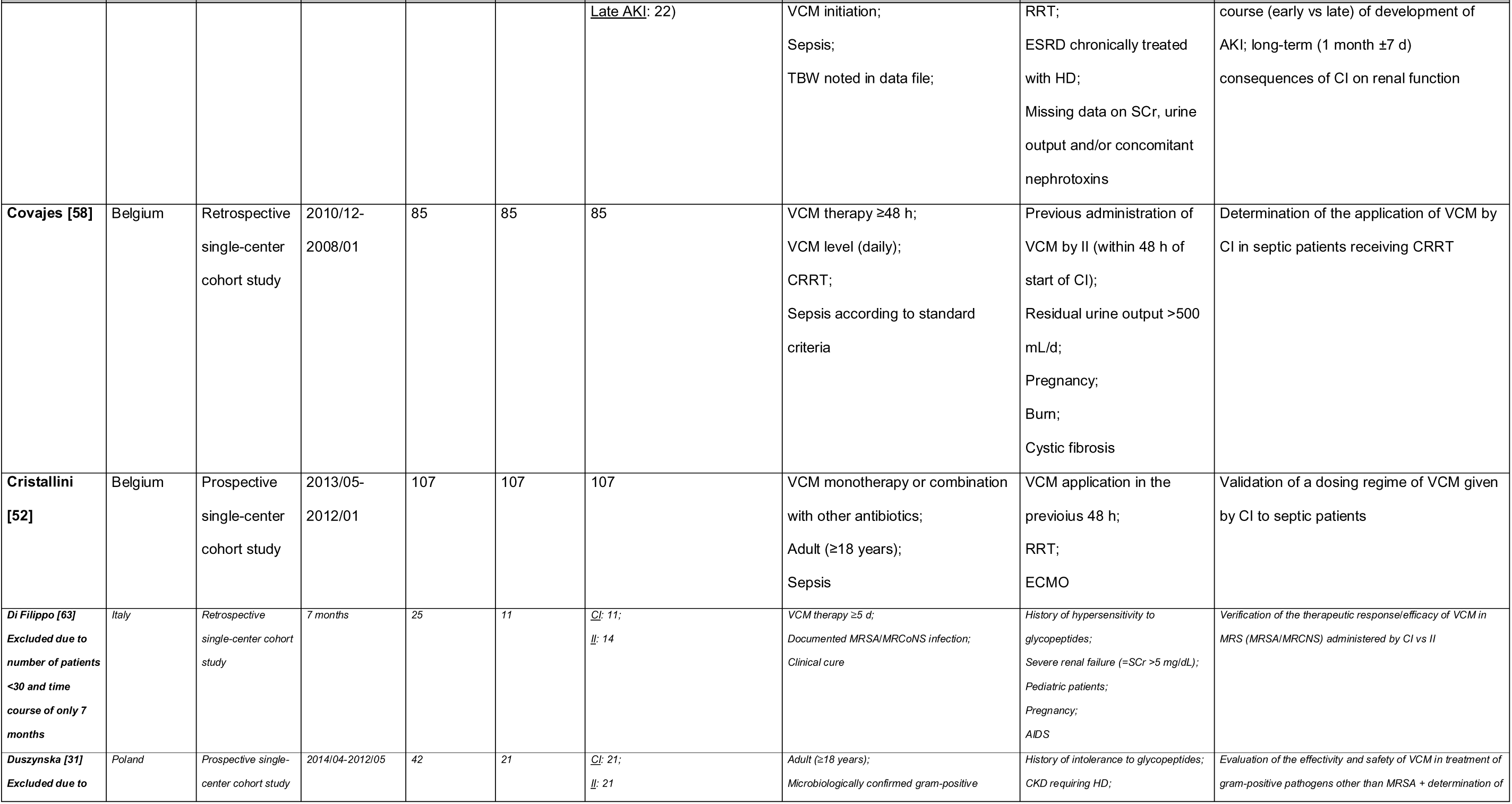
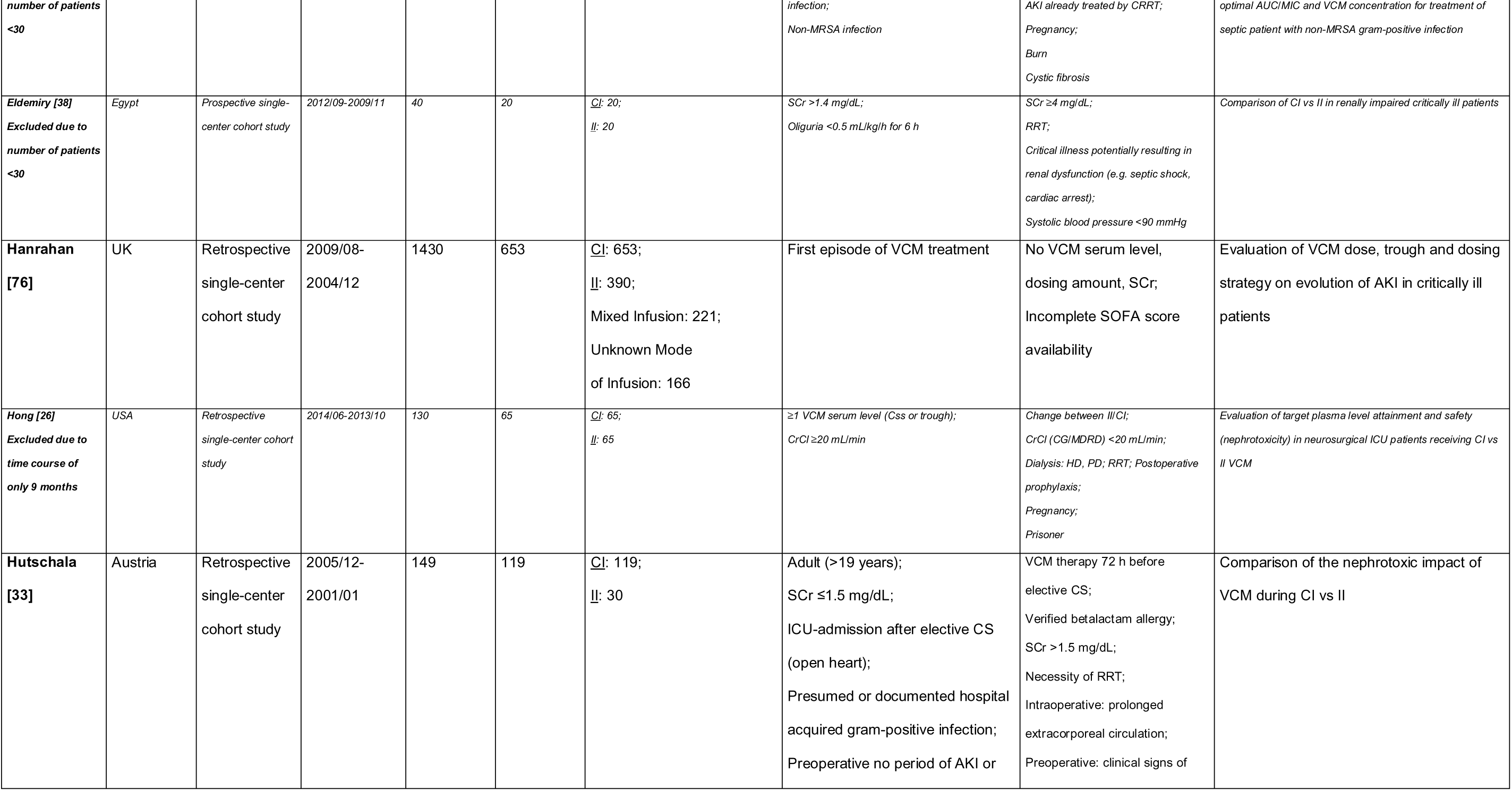
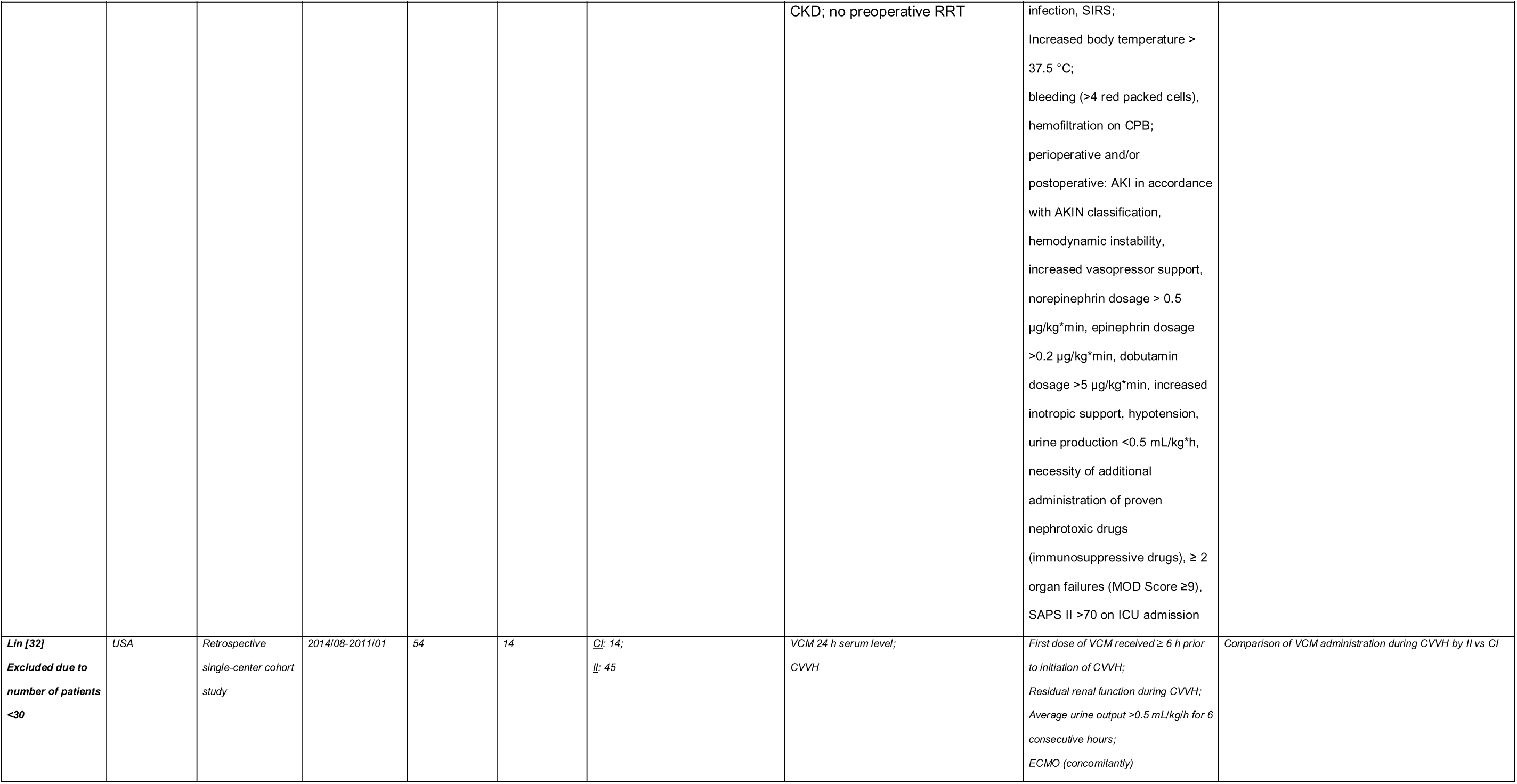
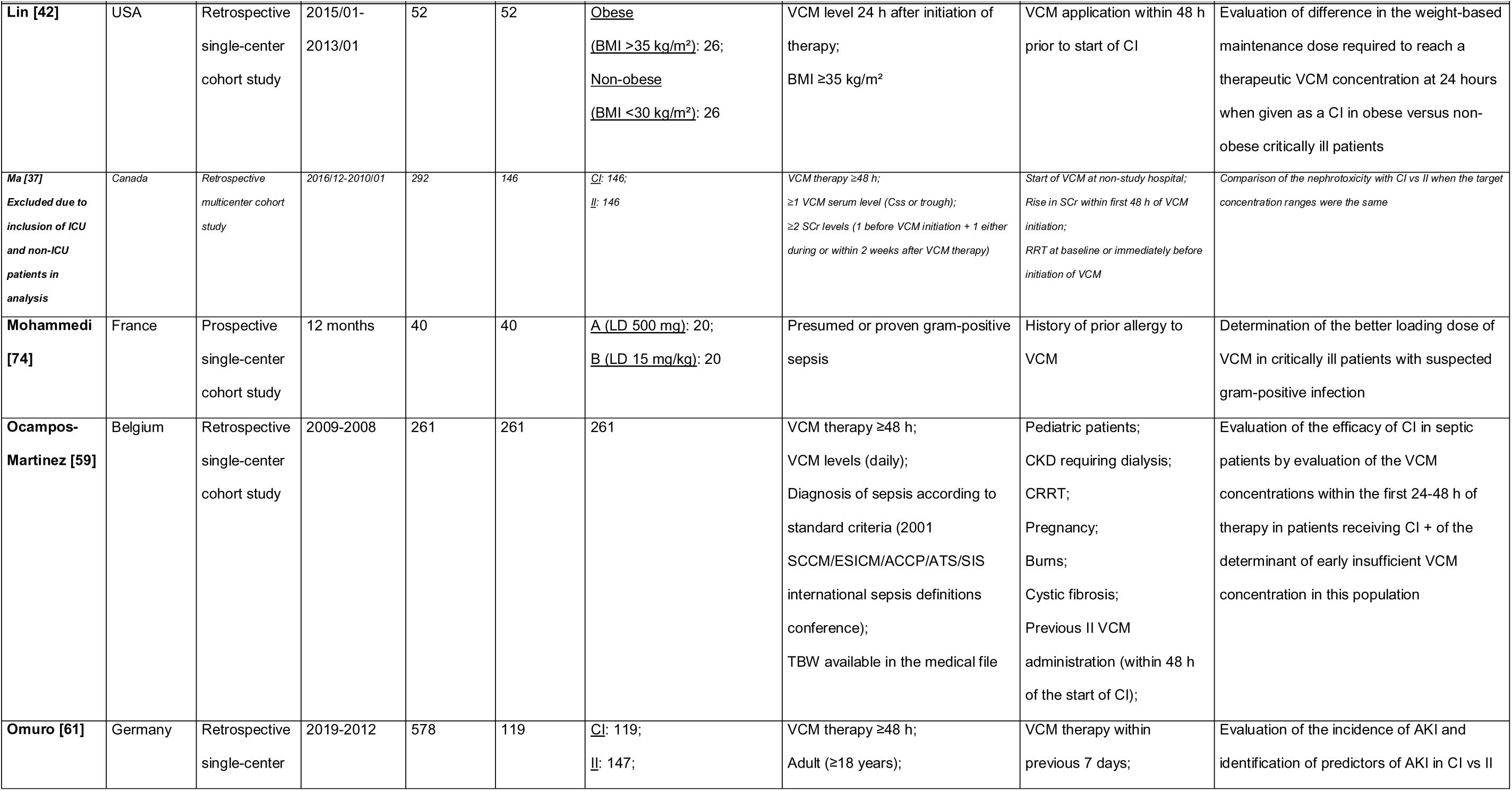
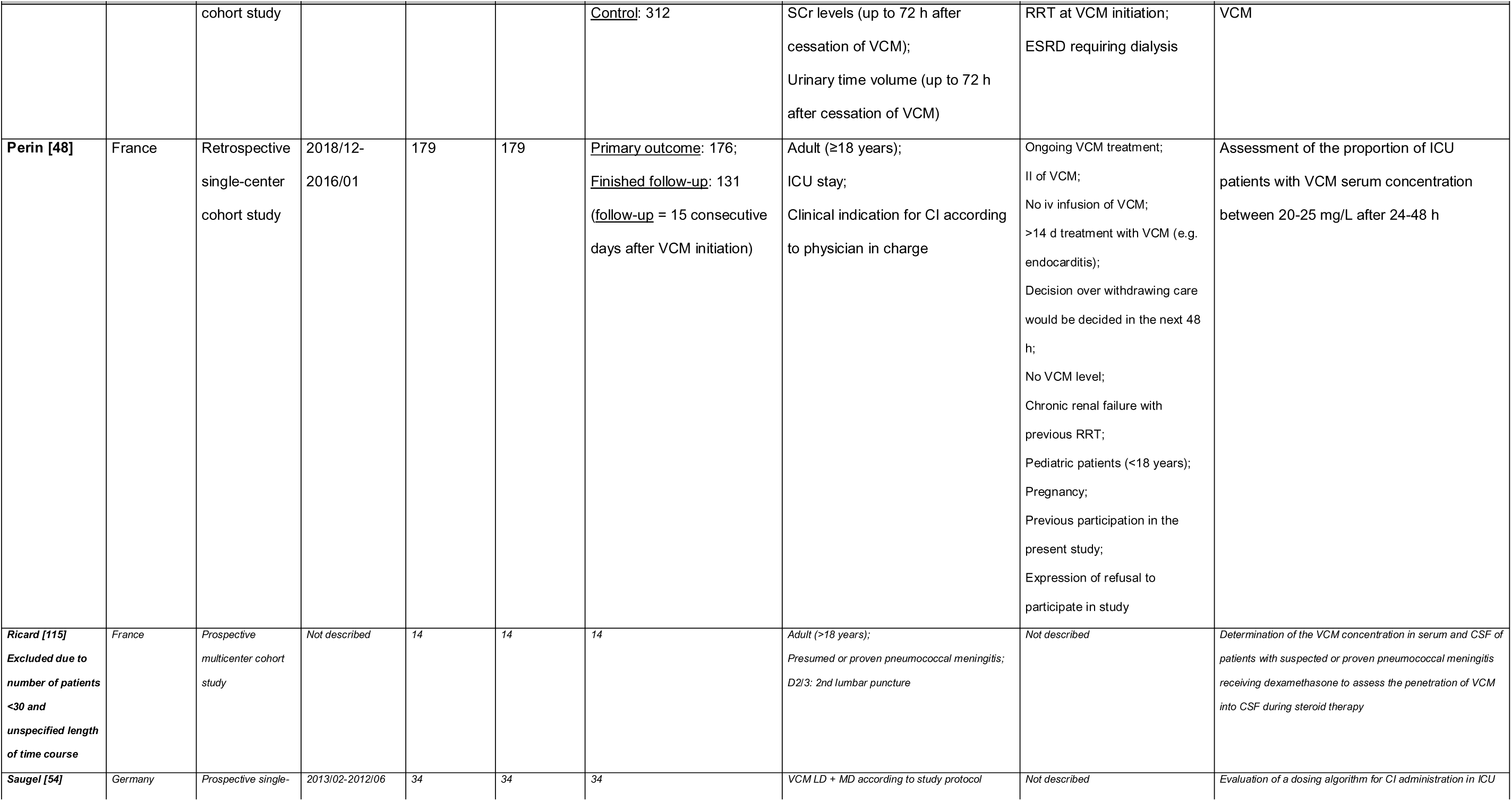
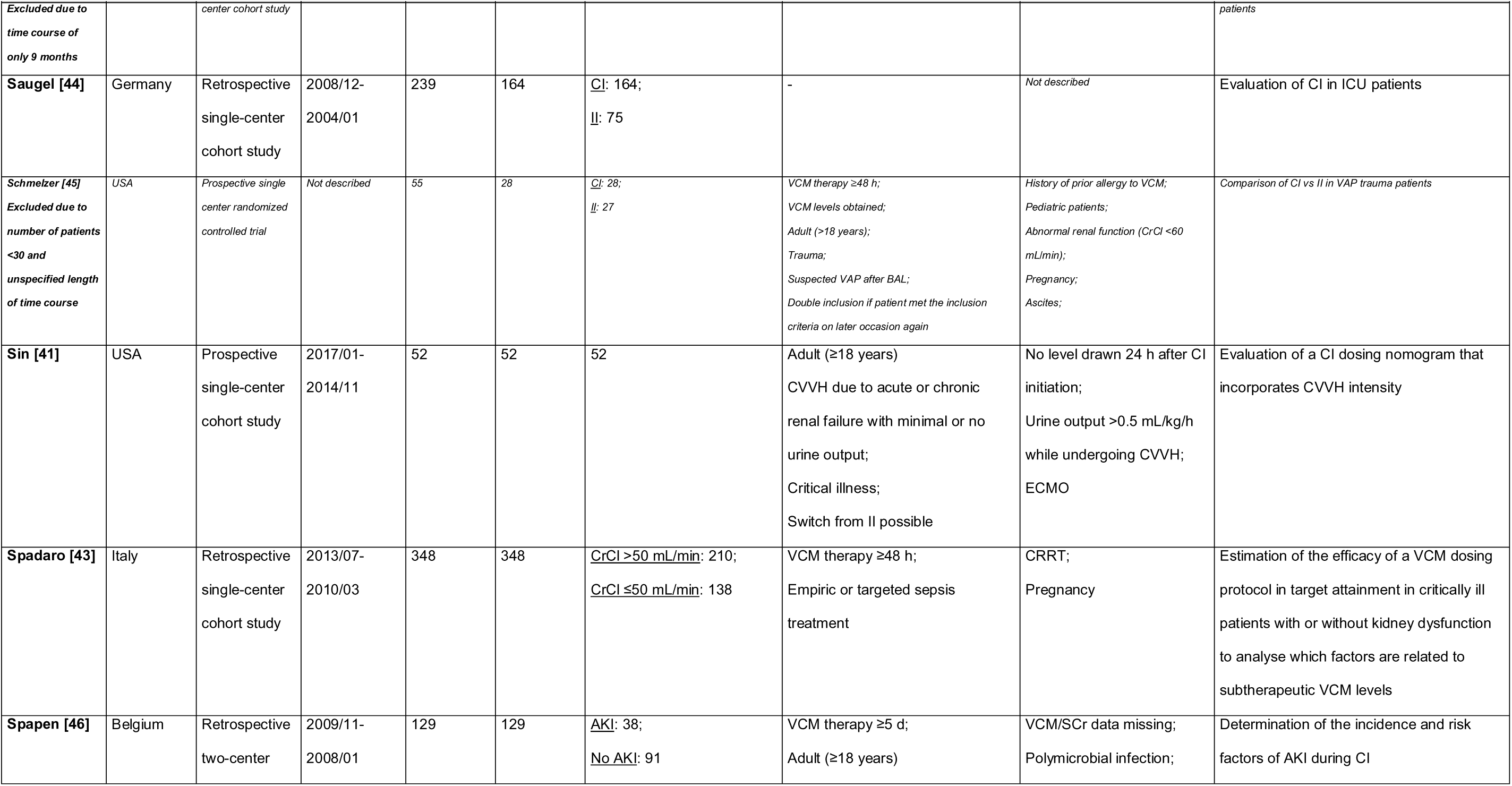
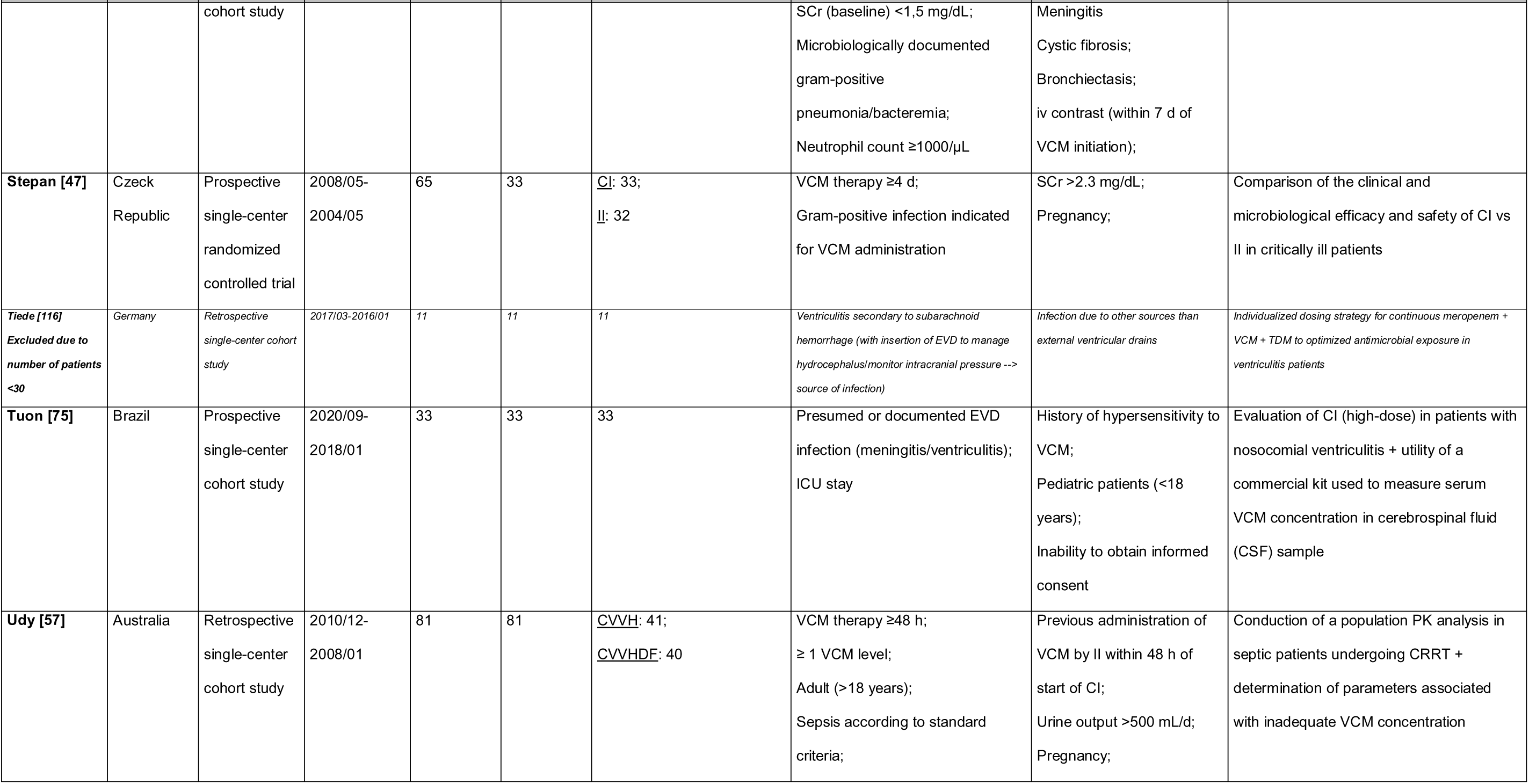
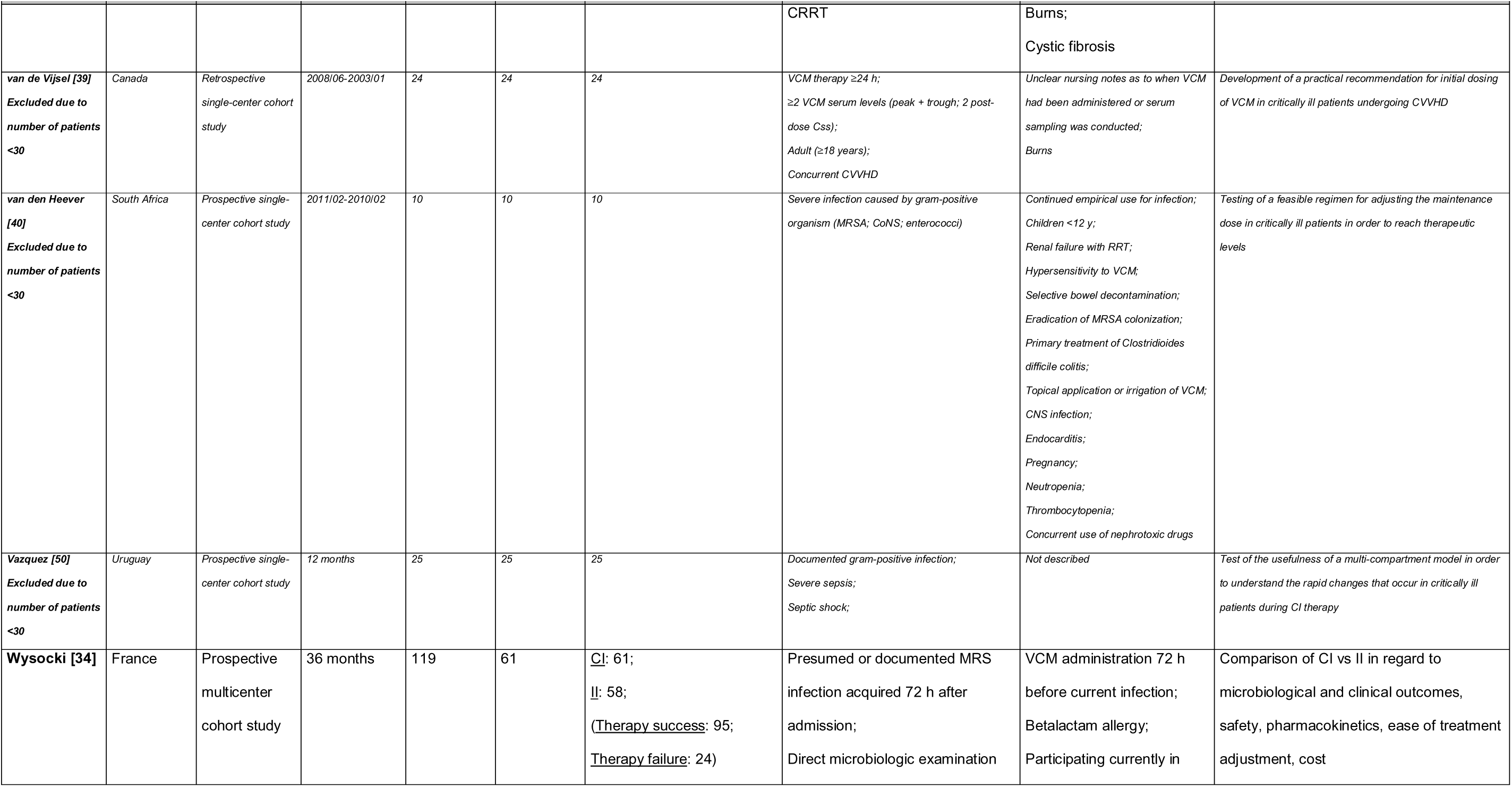
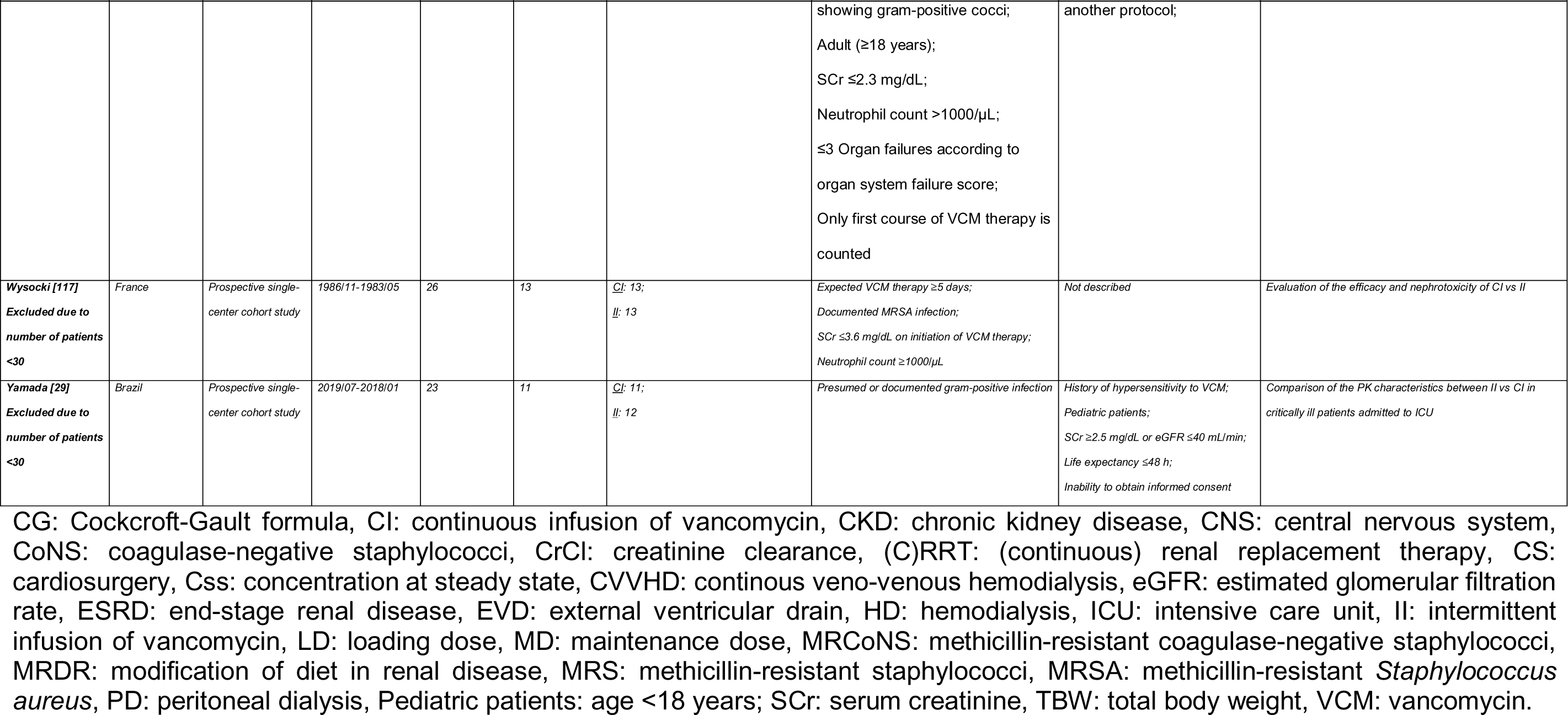
Overview of study designs, number of patients, inclusion and exclusion criteria and primary purpose of studies. Thirty-eight studies were identified in the initial full-text selection process that presented data of steady-state serum concentrations of continuously administered vancomycin (CI) and efficacy (clinical or microbiological success (e.g. cure) or failure (e.g. mortality)) or safety (e.g. acute renal failure). Only twenty-one studies made it into the final analysis. Studies excluded from the final selection because of an increased risk of bias (e.g. unspecified time course or time course of less than 12 months or a CI study population of less than 30 patients) are written in italics.

### 9.2 Study population

**Table S2:**
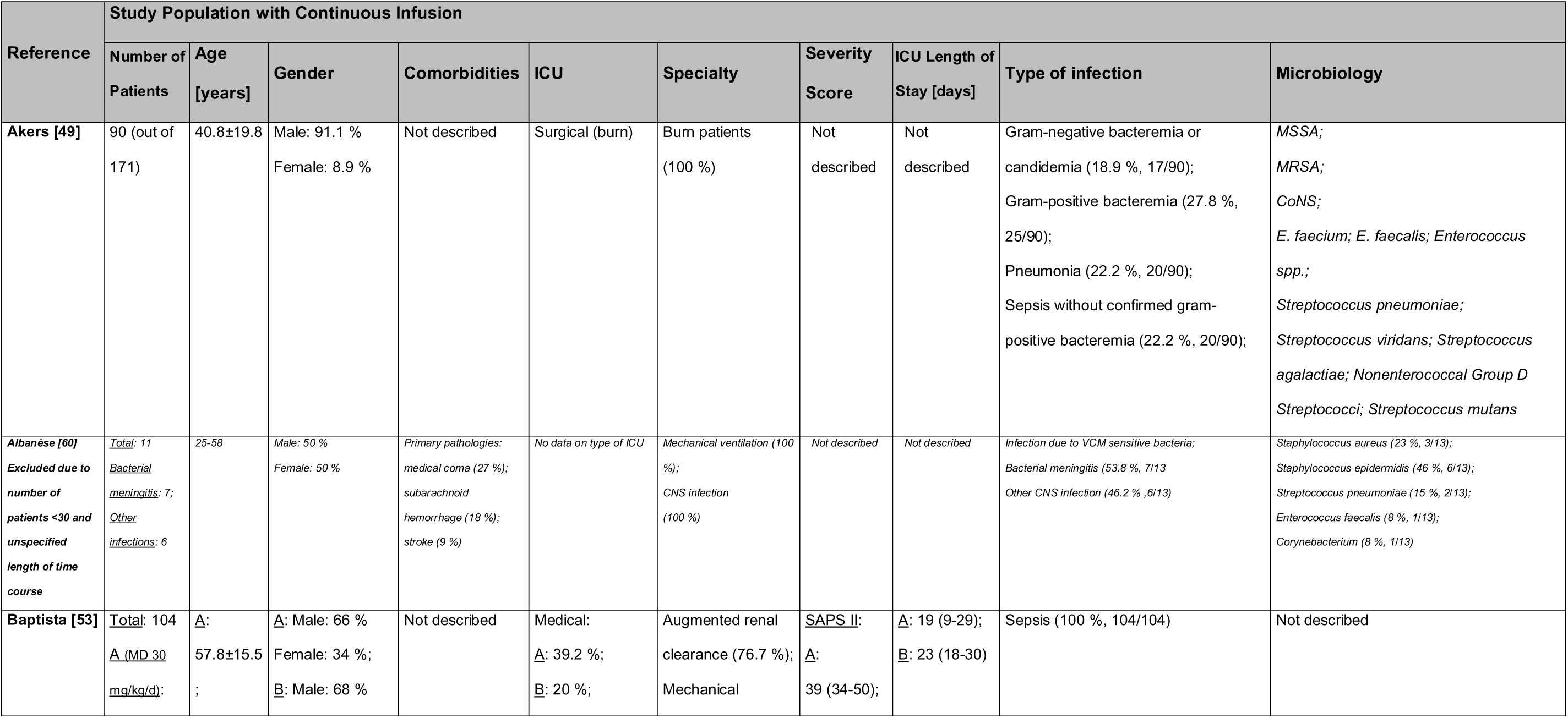
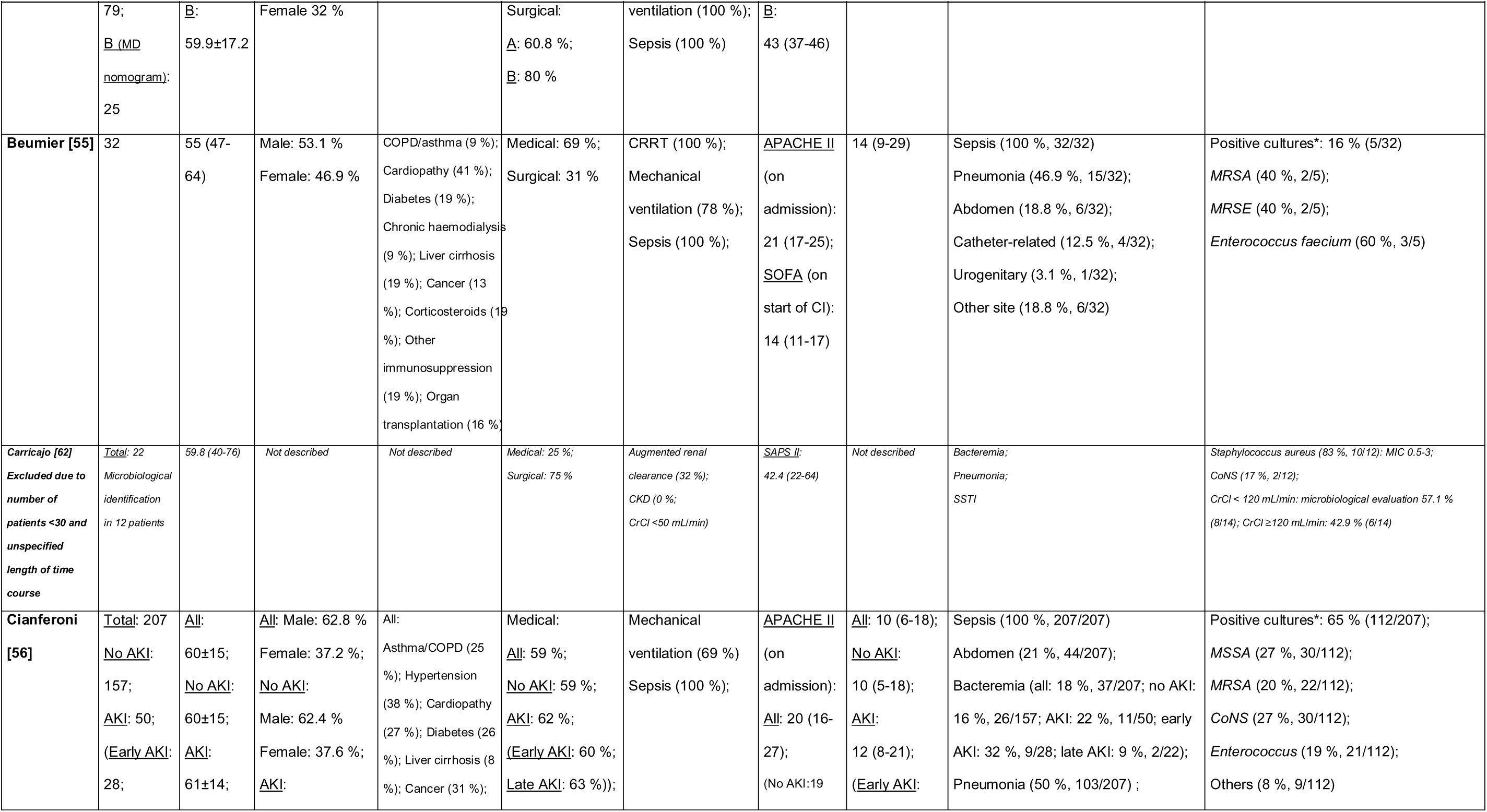
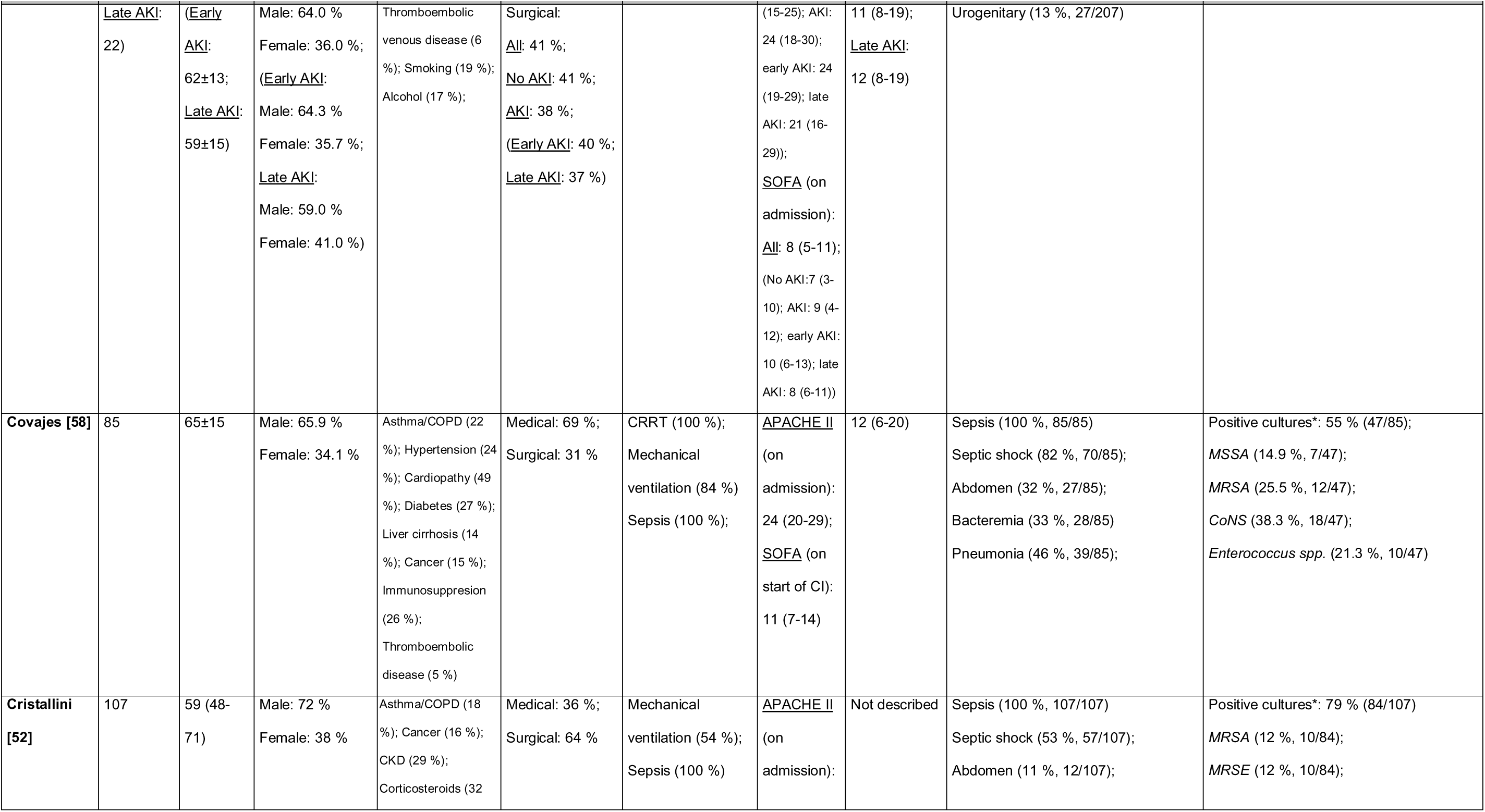
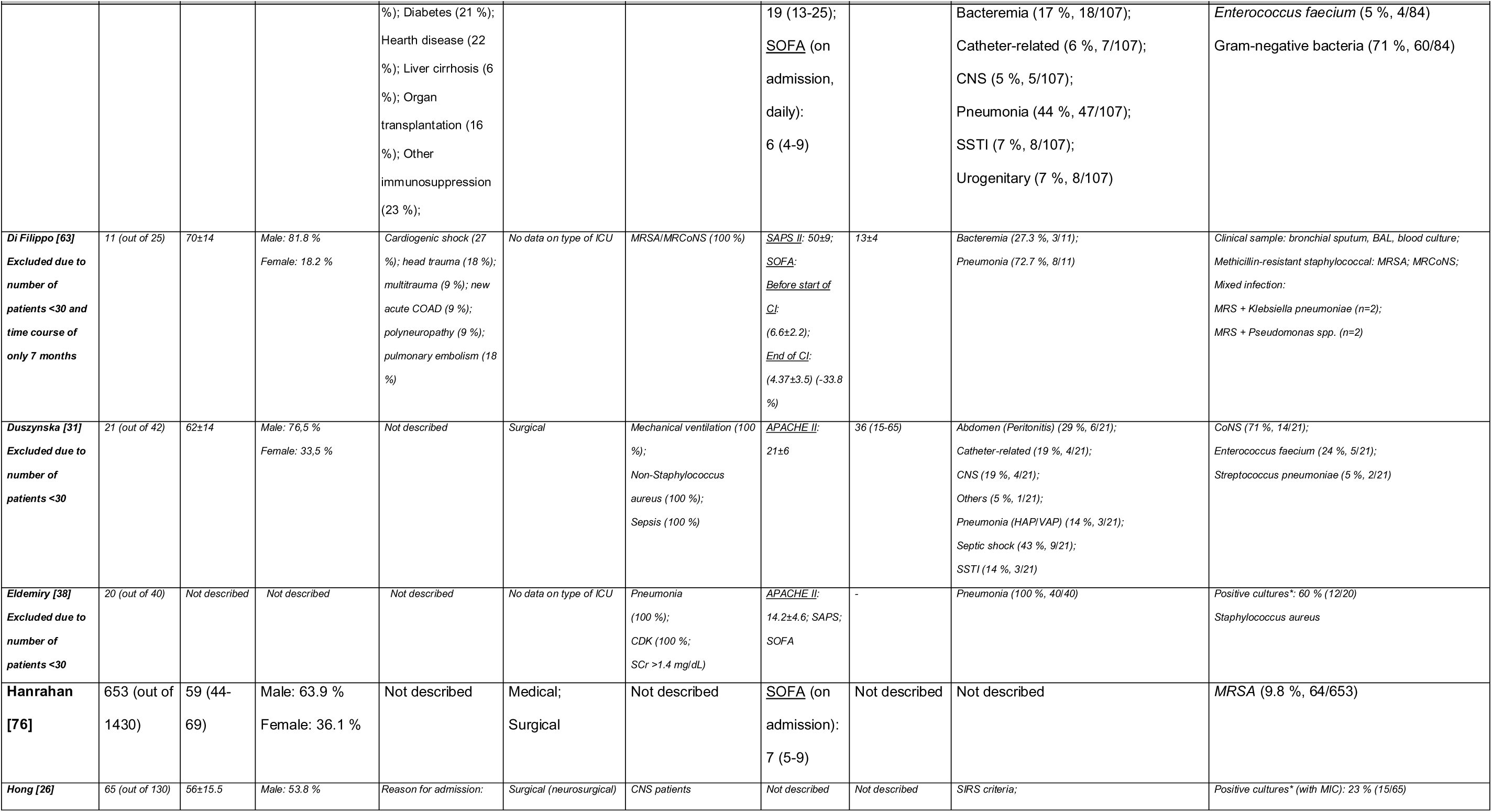
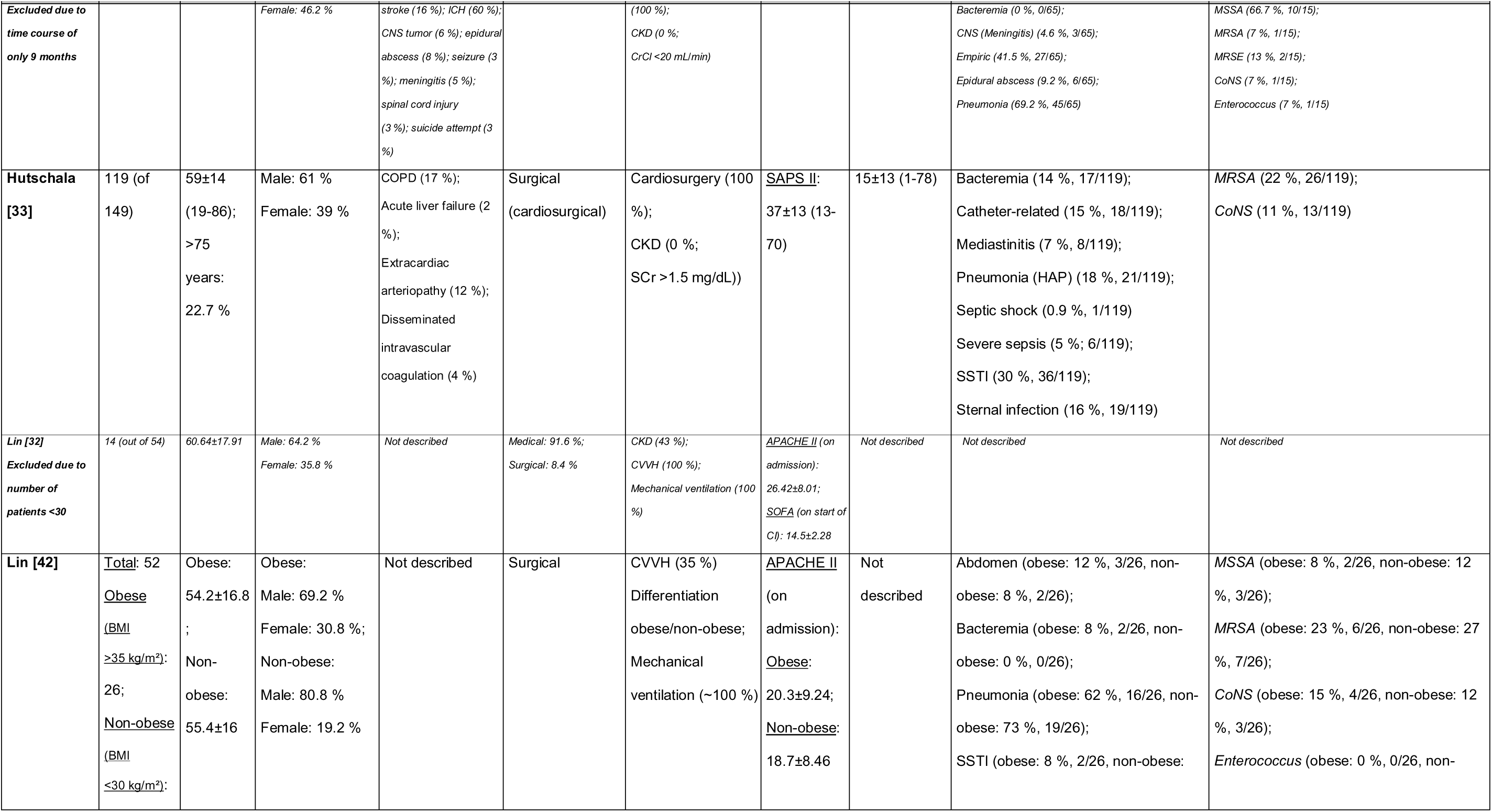
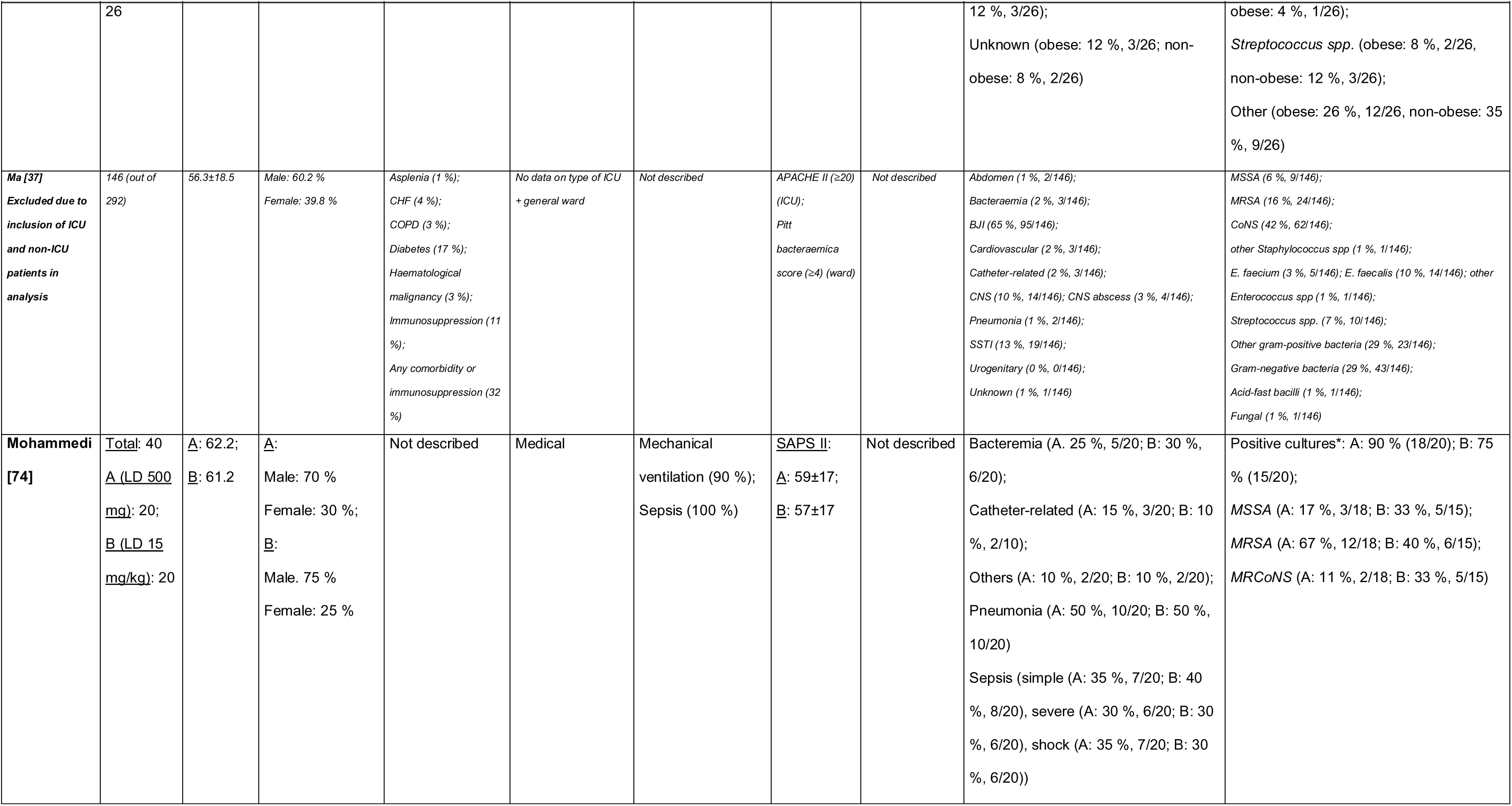
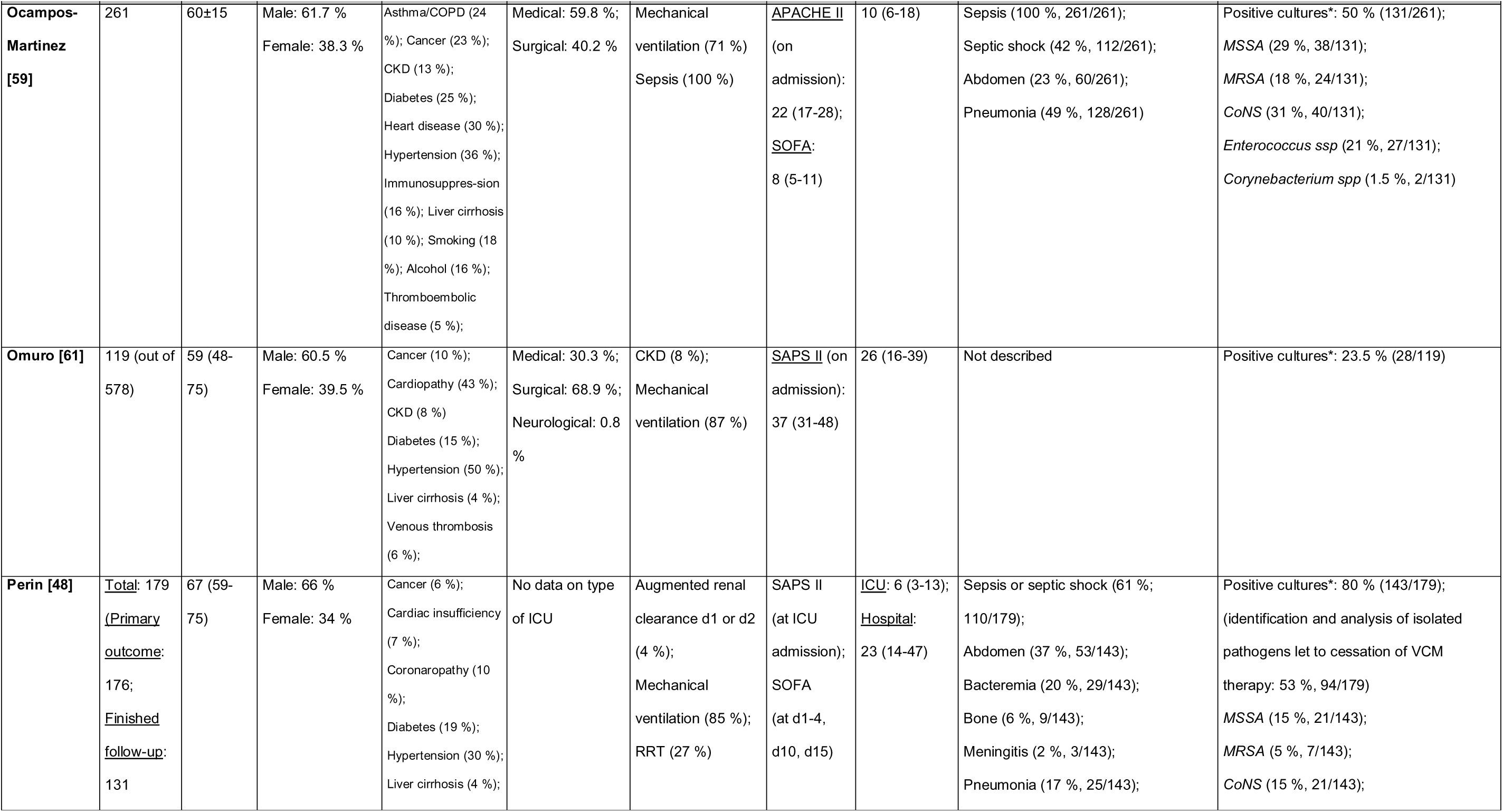
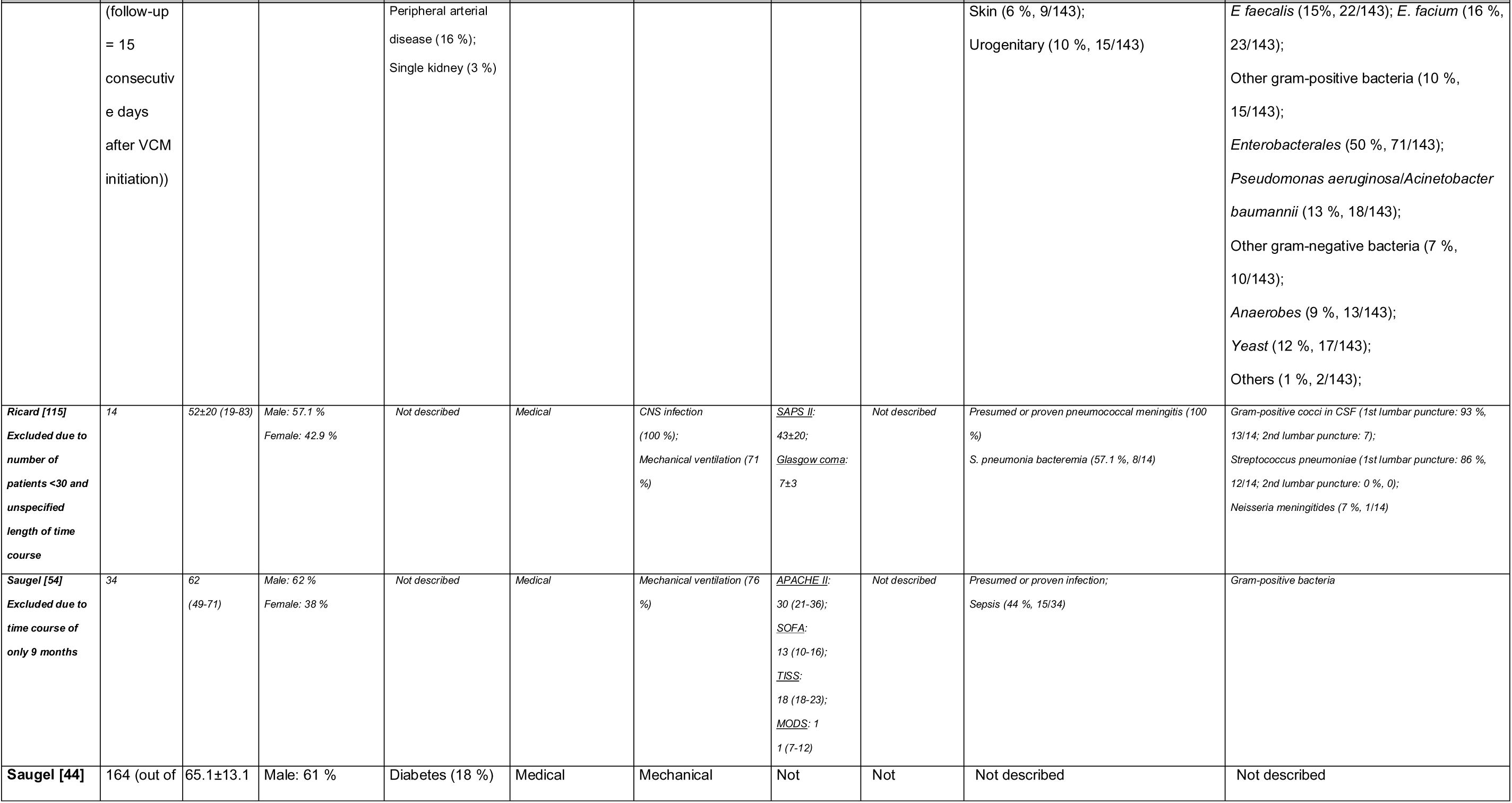
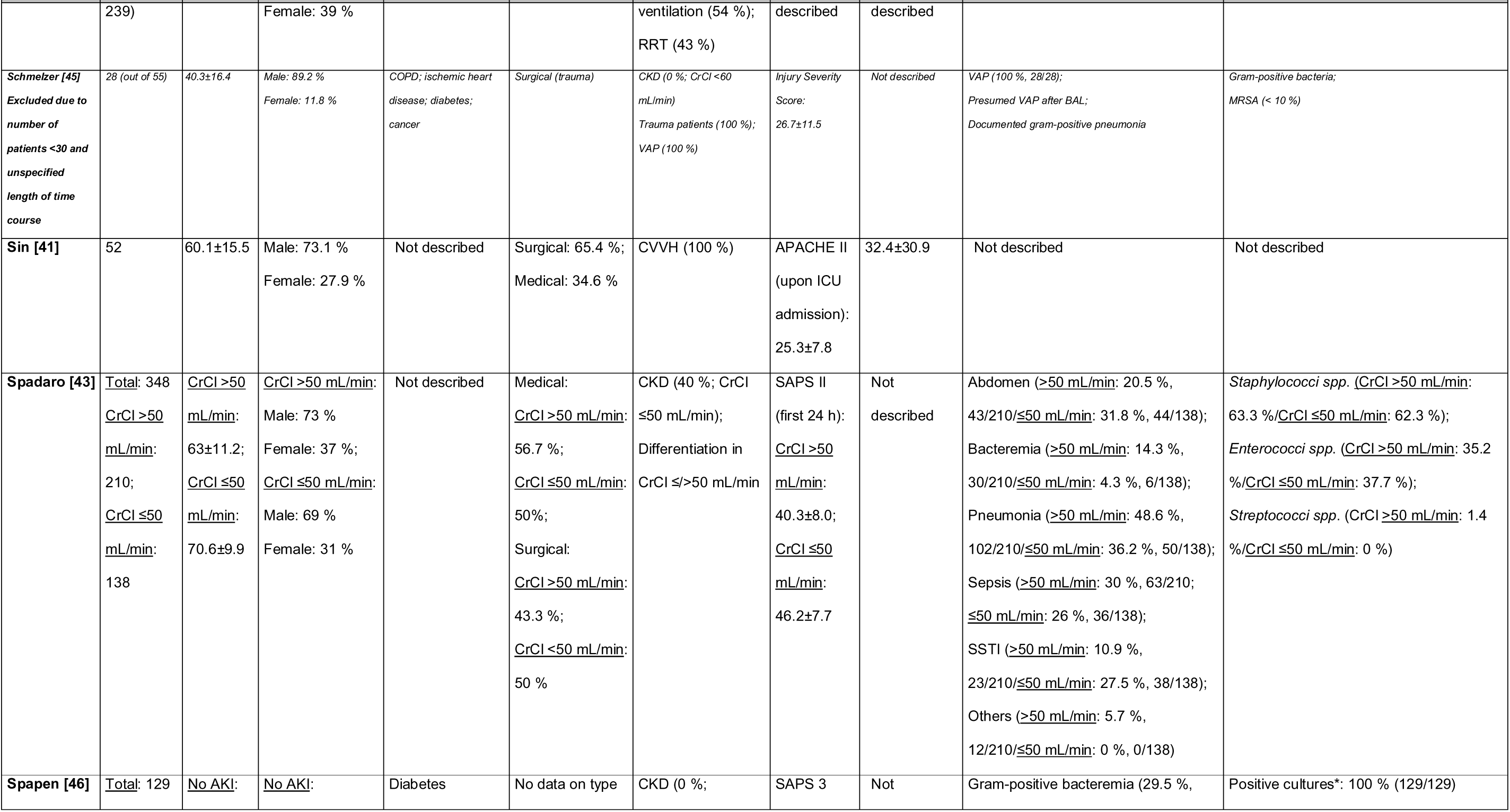
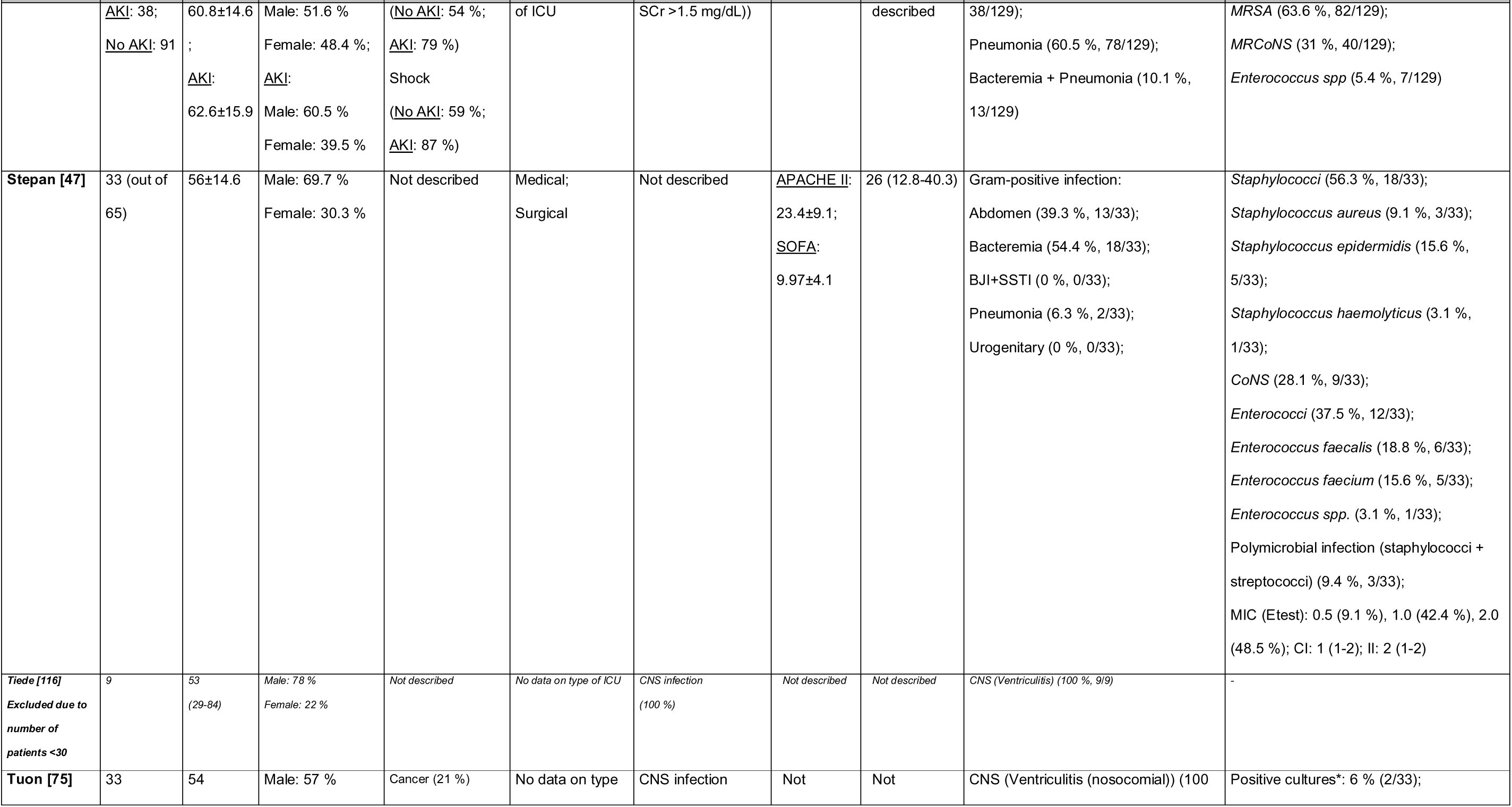
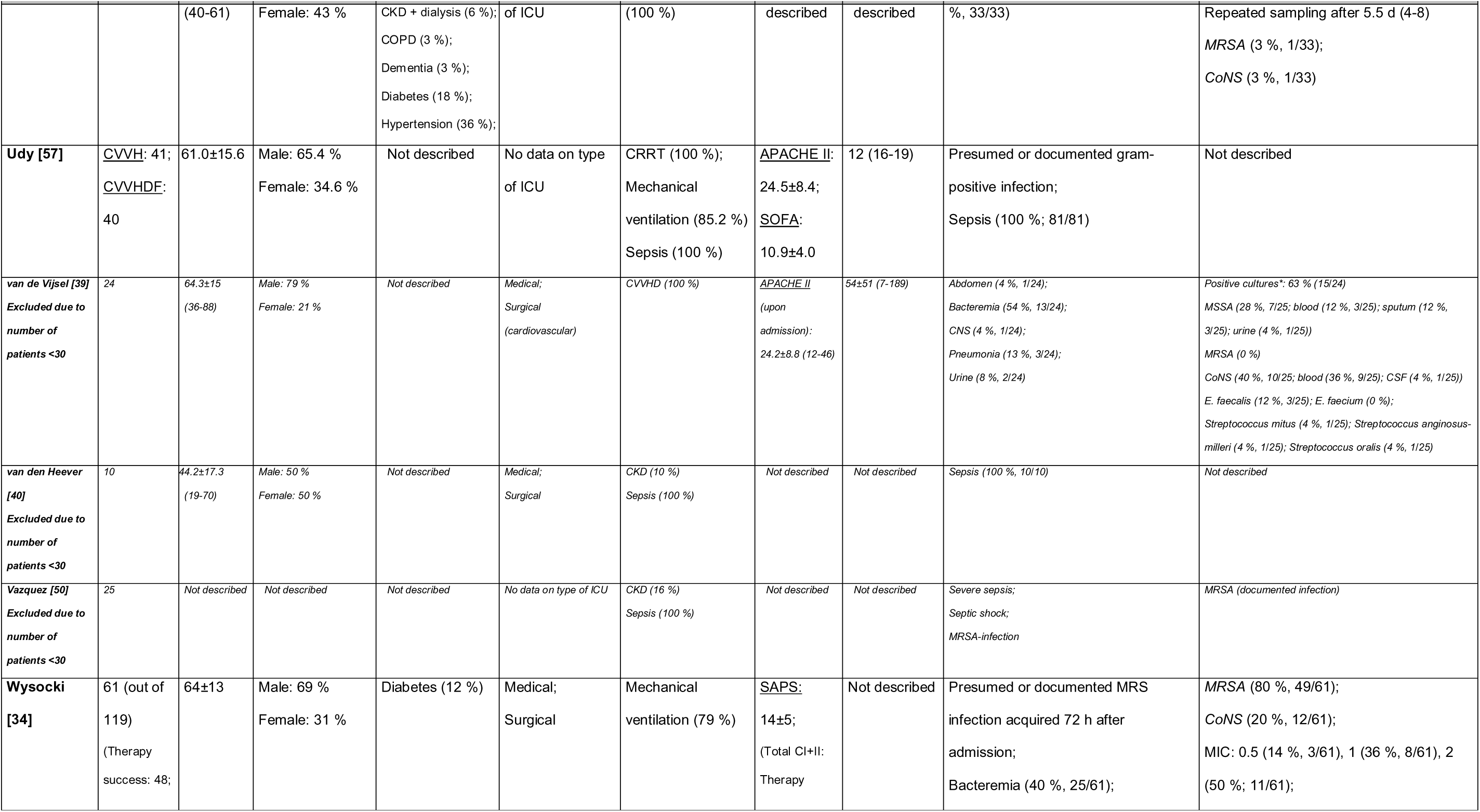
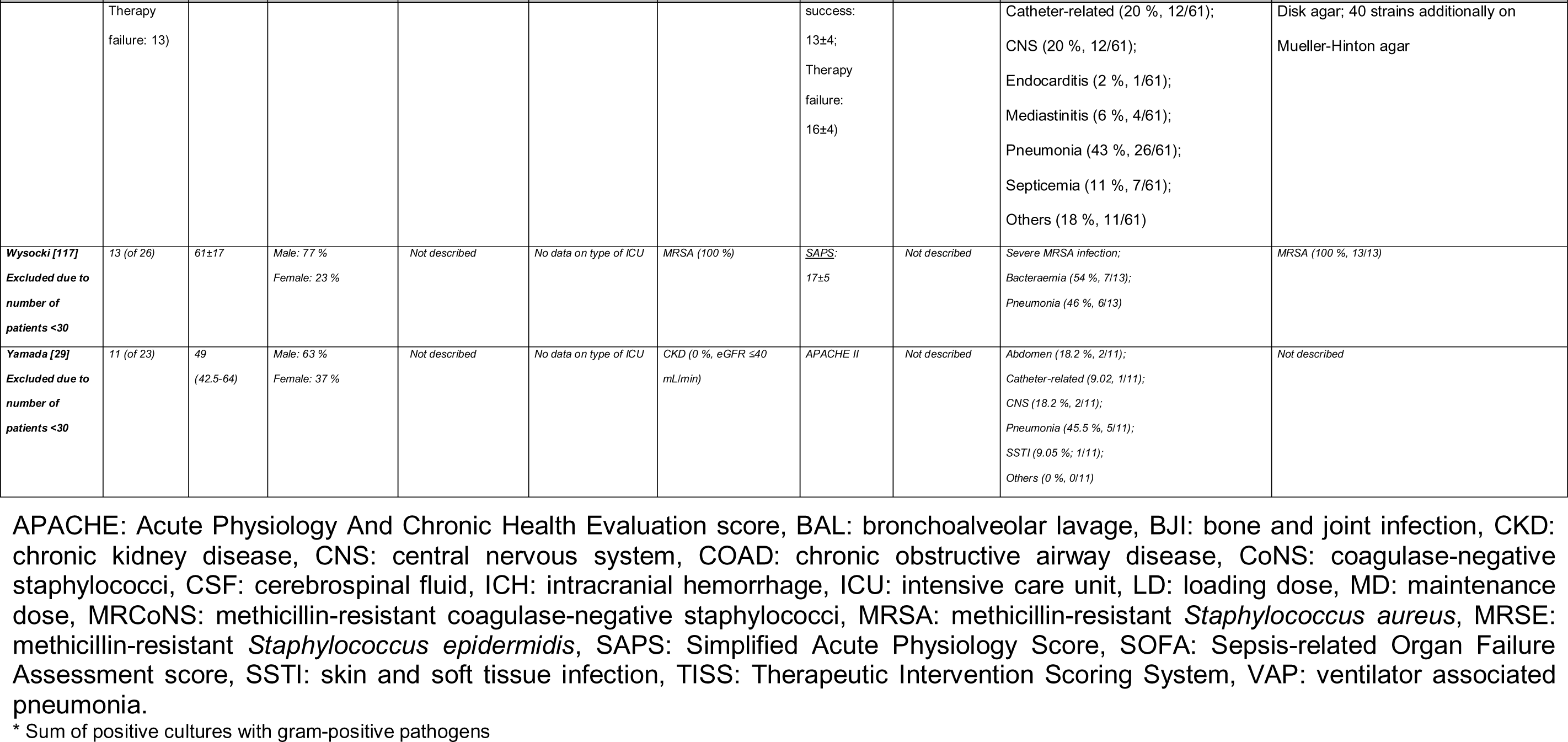
Overview of the study population. Thirty-eight studies were identified in the initial full-text selection process that presented data of steady-state serum concentrations of continuously administered vancomycin (CI) and efficacy (clinical or microbiological success (e.g. cure) or failure (e.g. mortality)) or safety (e.g. acute renal failure). Only twenty-one studies made it into the final analysis. Studies excluded from the final selection because of an increased risk of bias (e.g. unspecified or time course of less than 12 months or a CI study population of less than 30 patients) are written in italics.

### 9.3 Characteristics of Continuously Administered Vancomycin Therapy

**Table S3:**
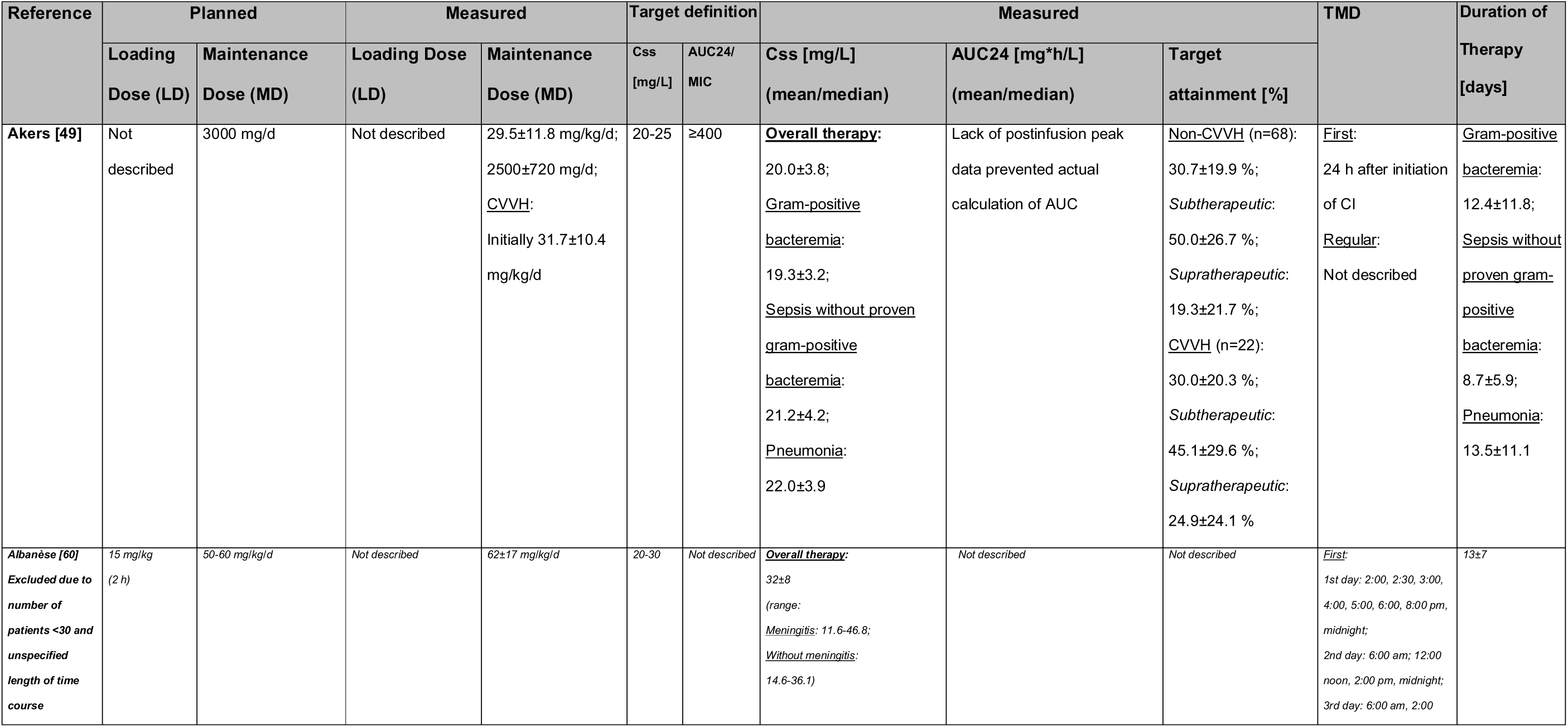
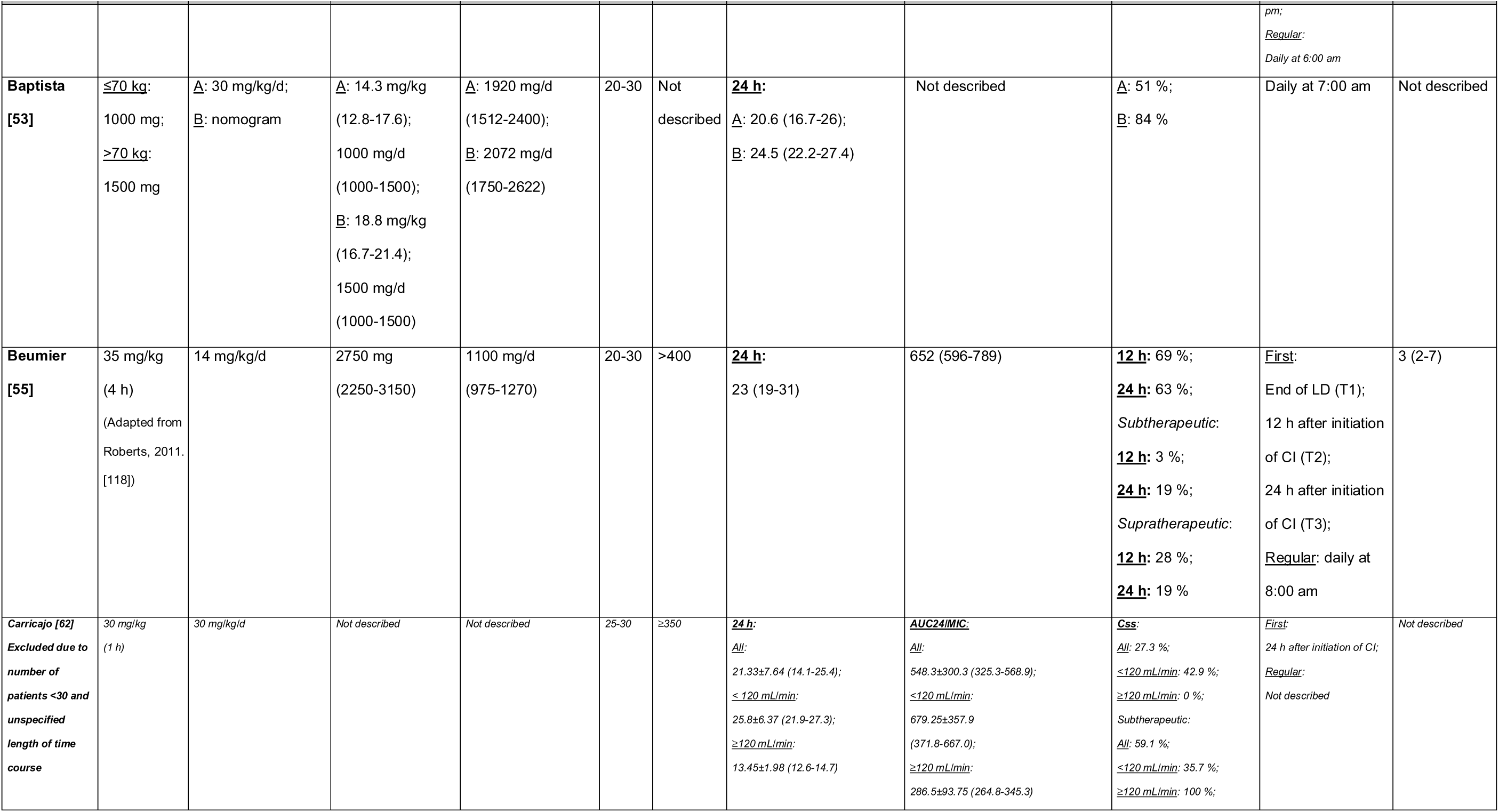
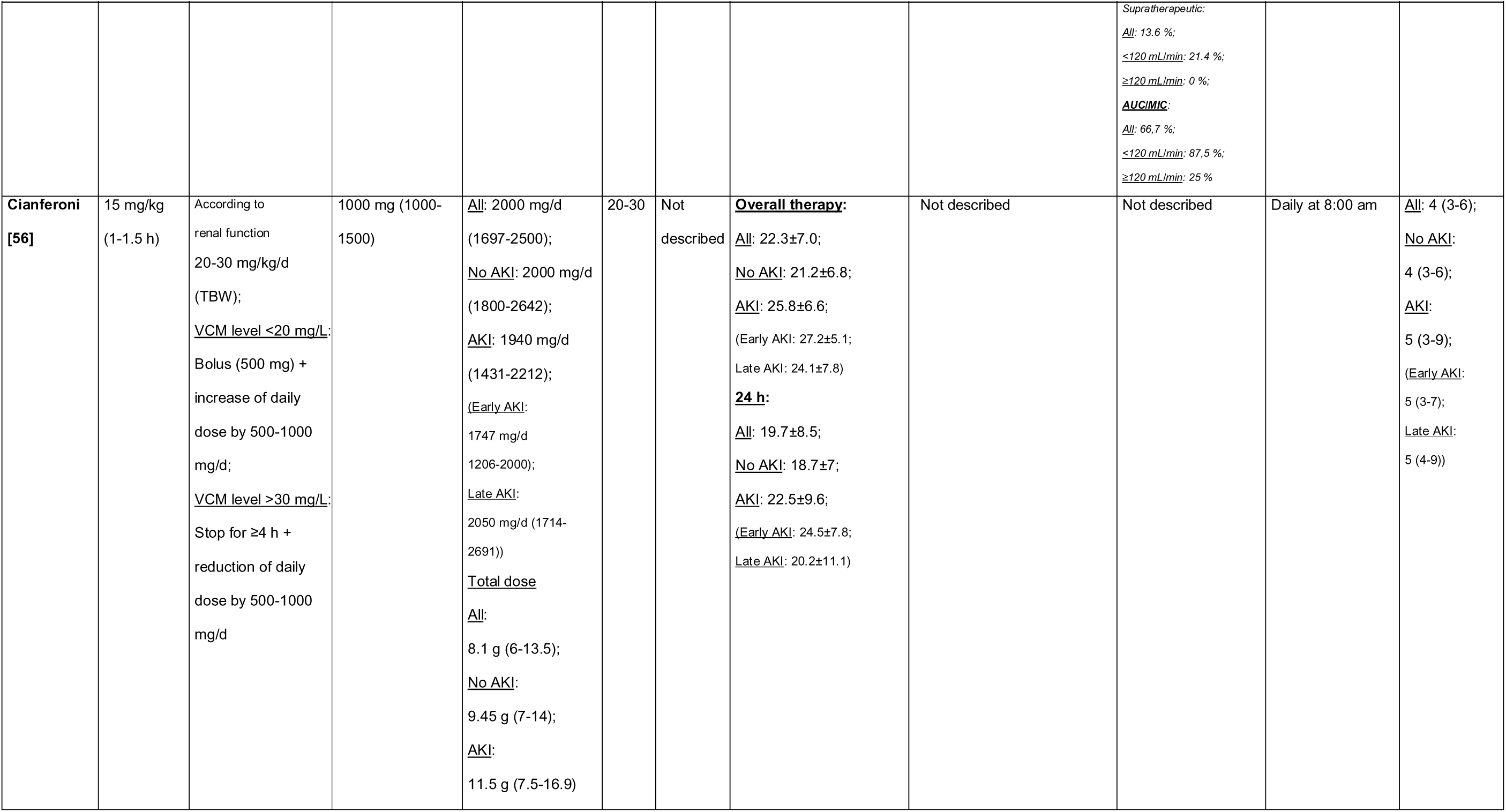
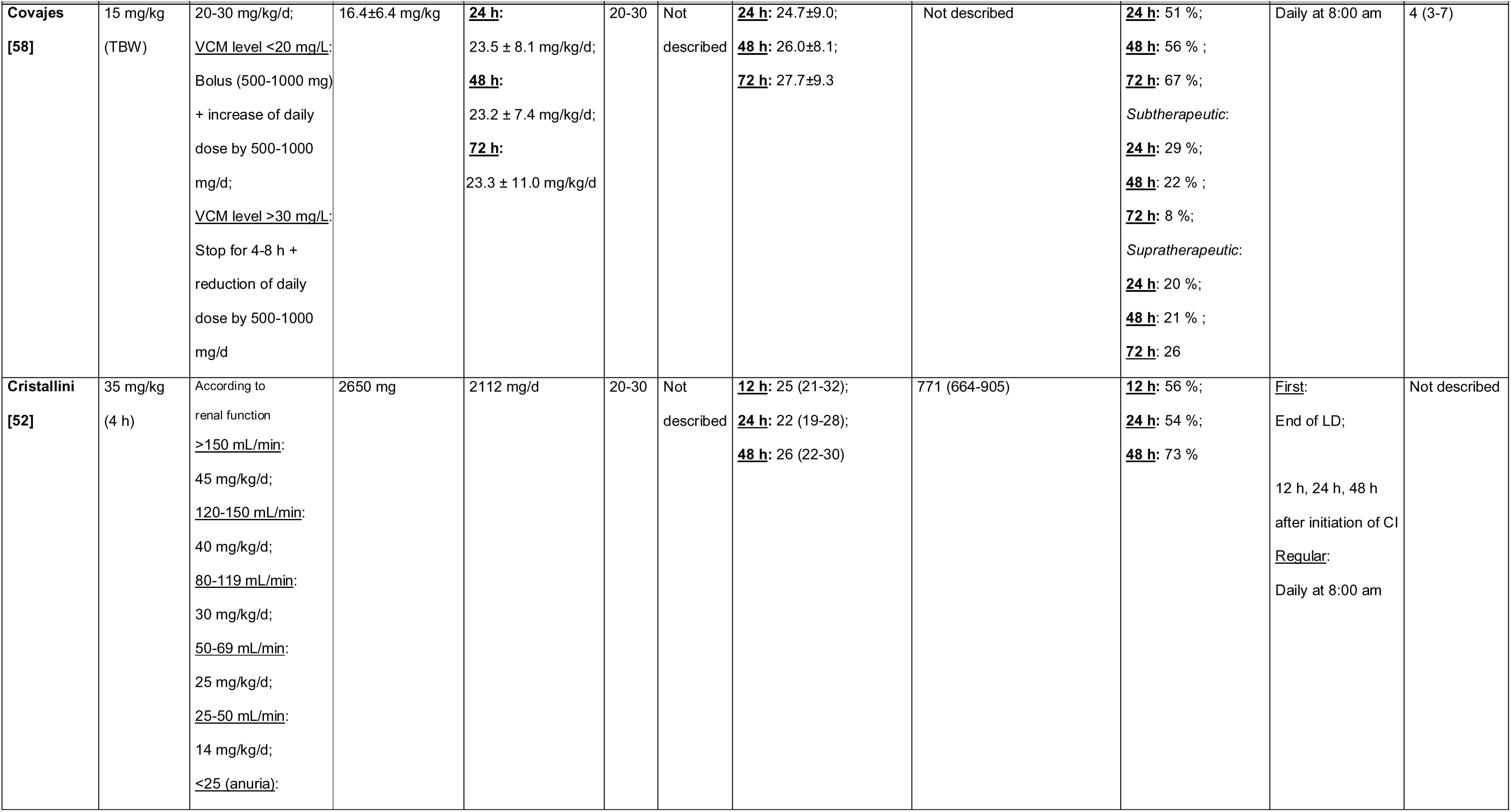
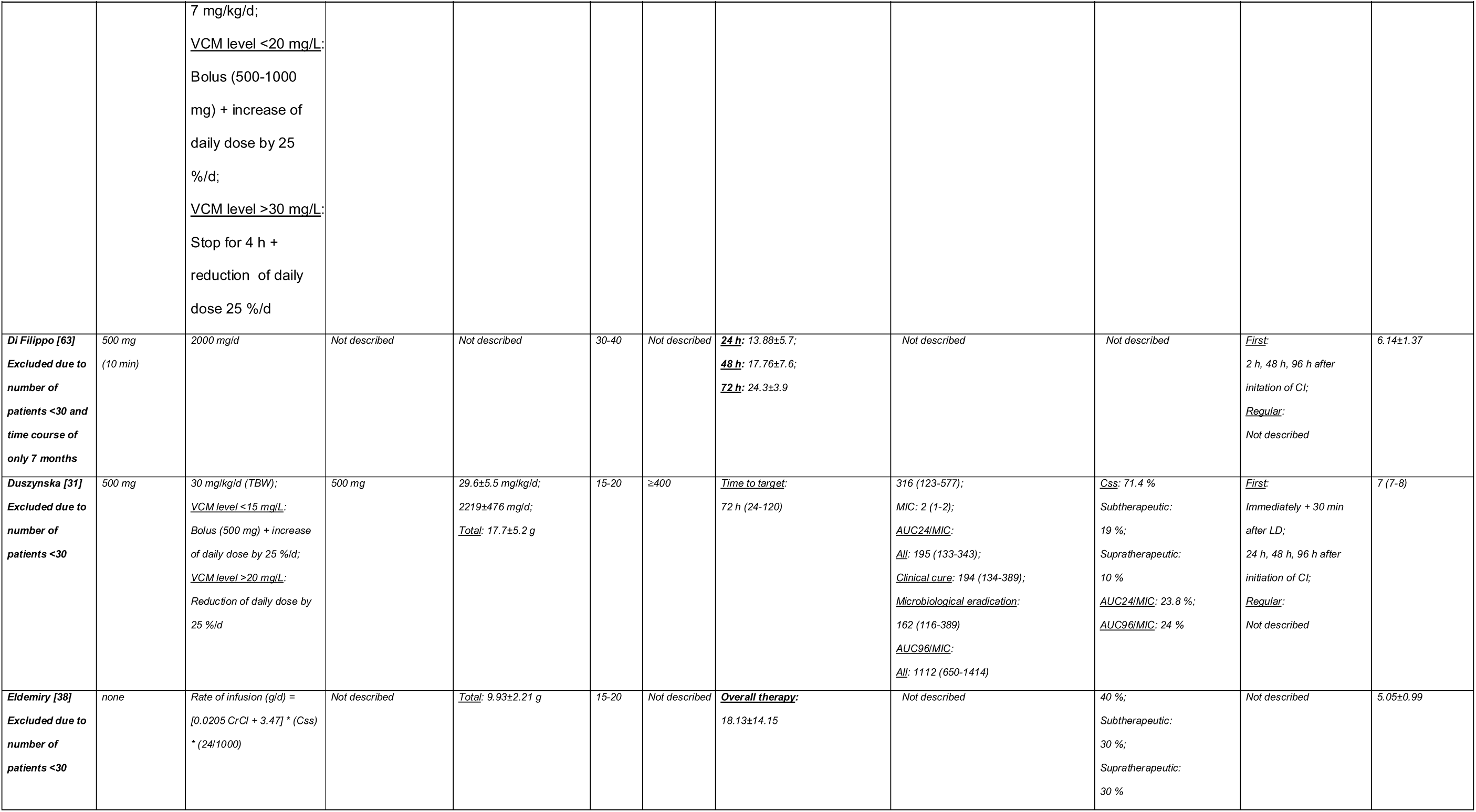
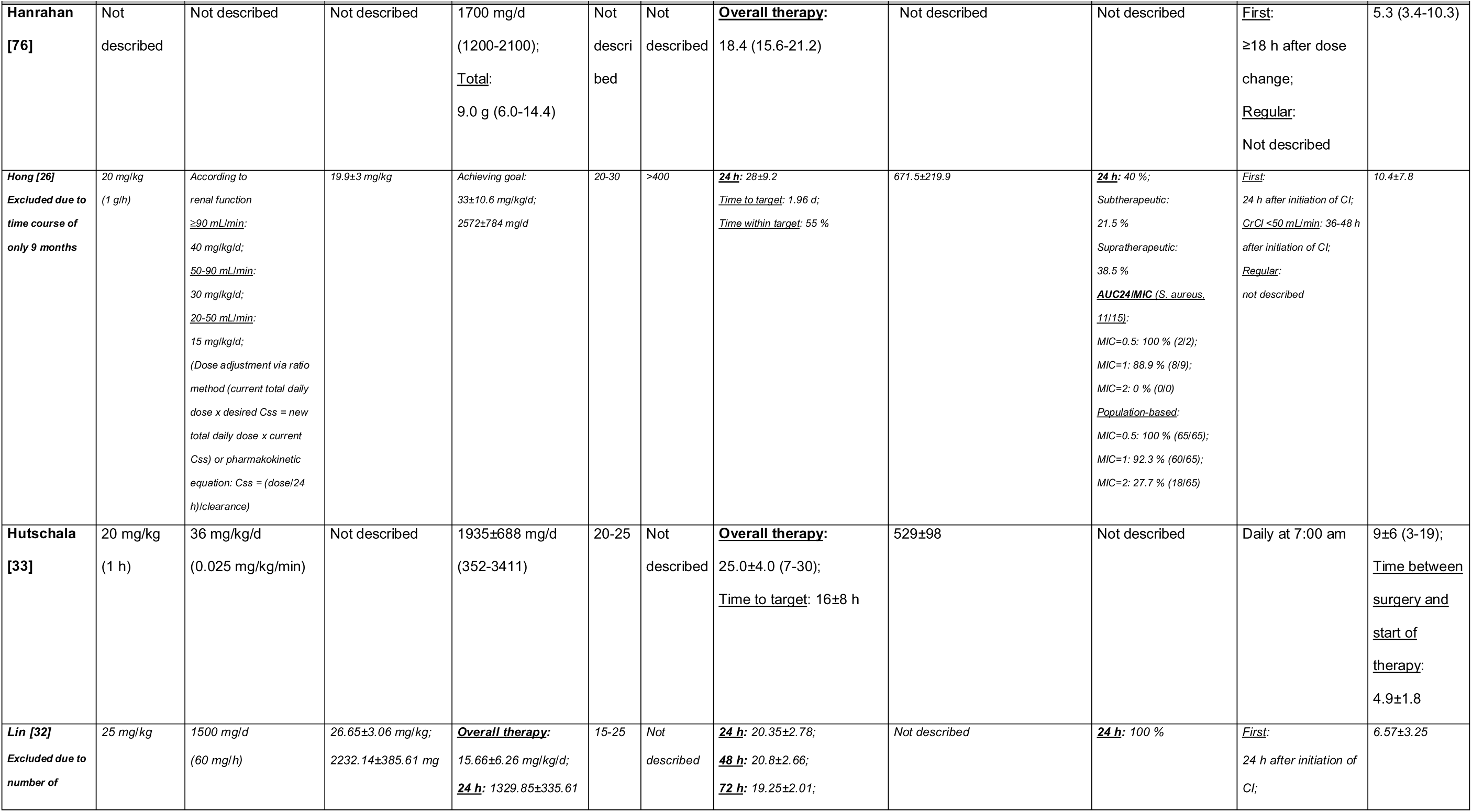
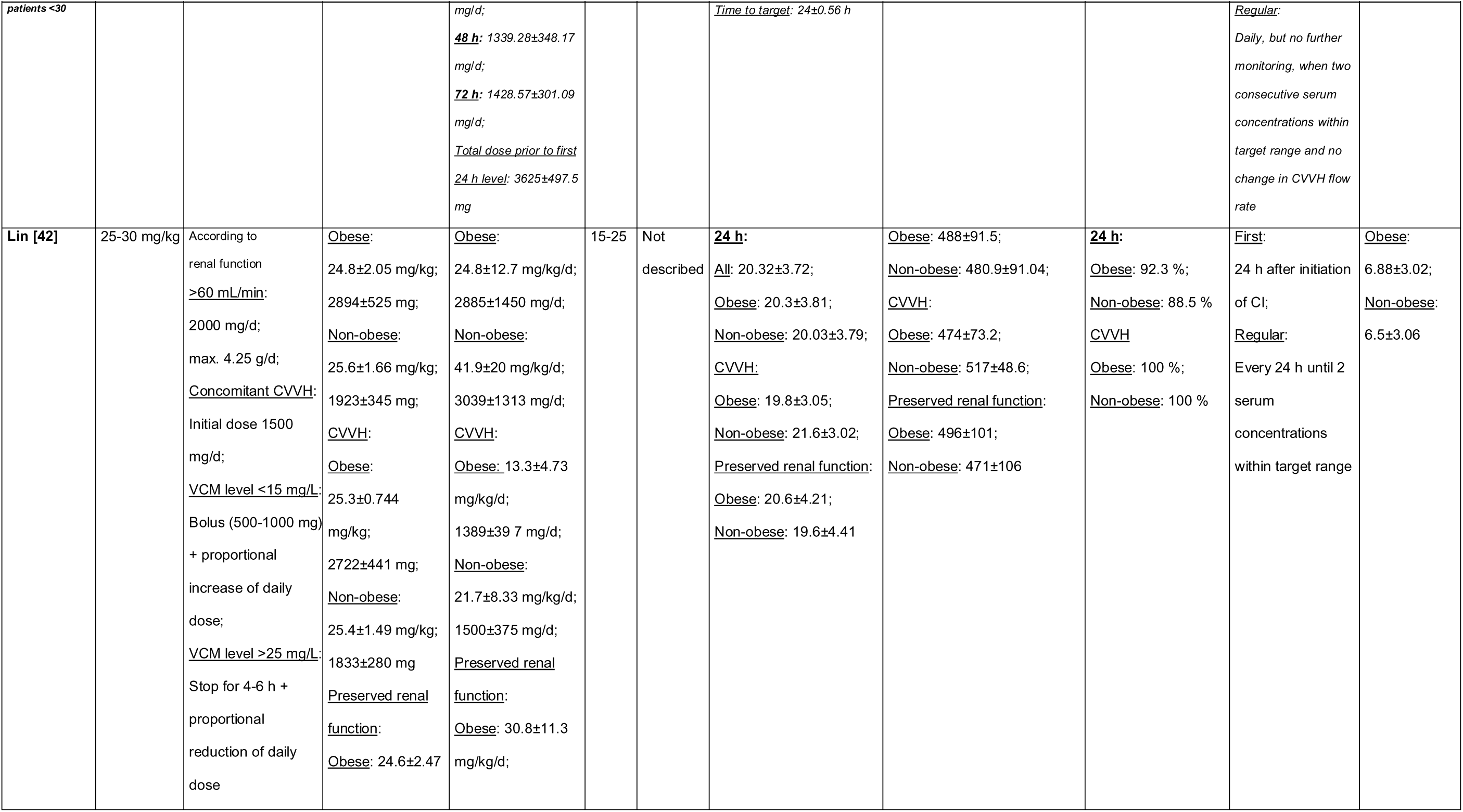
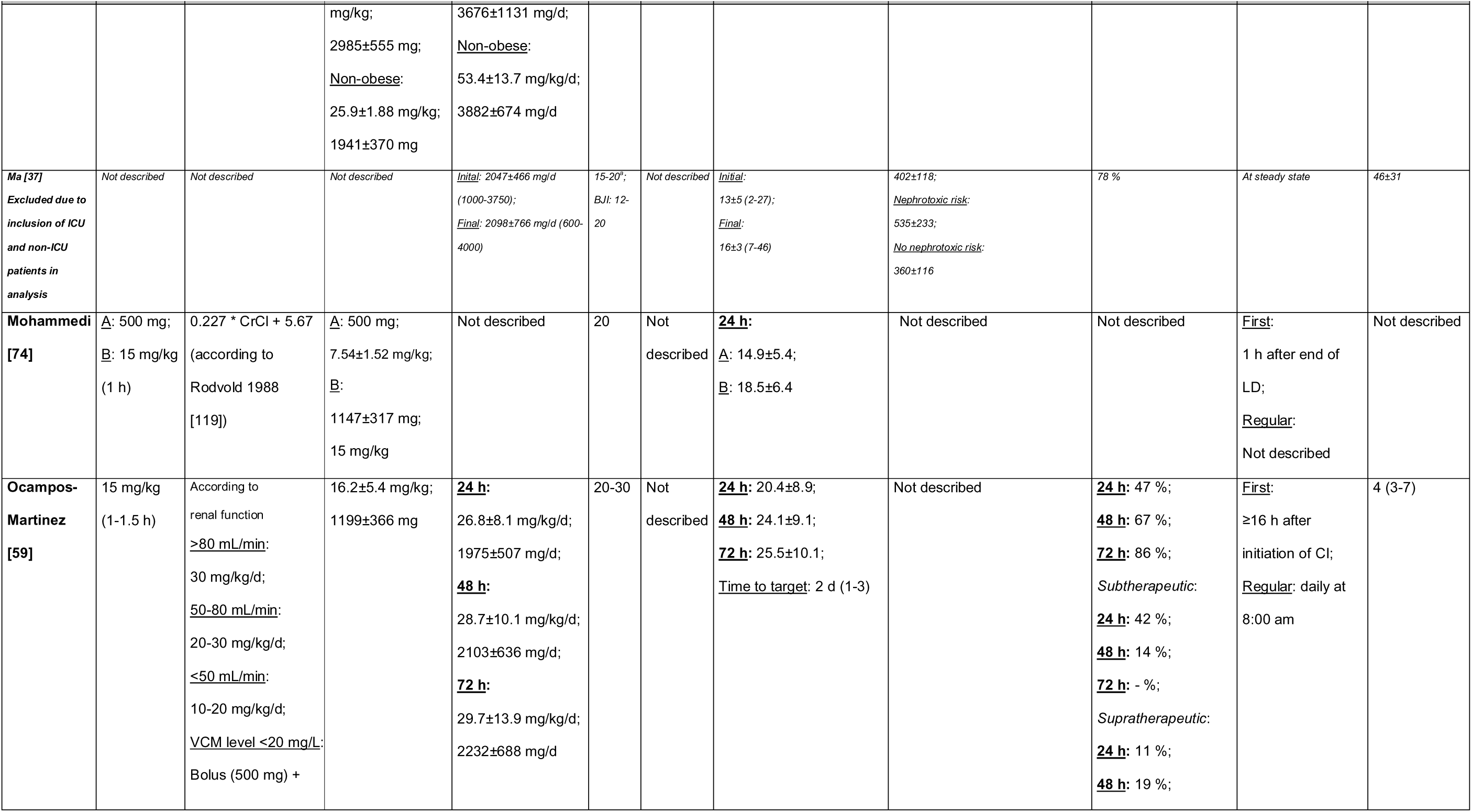
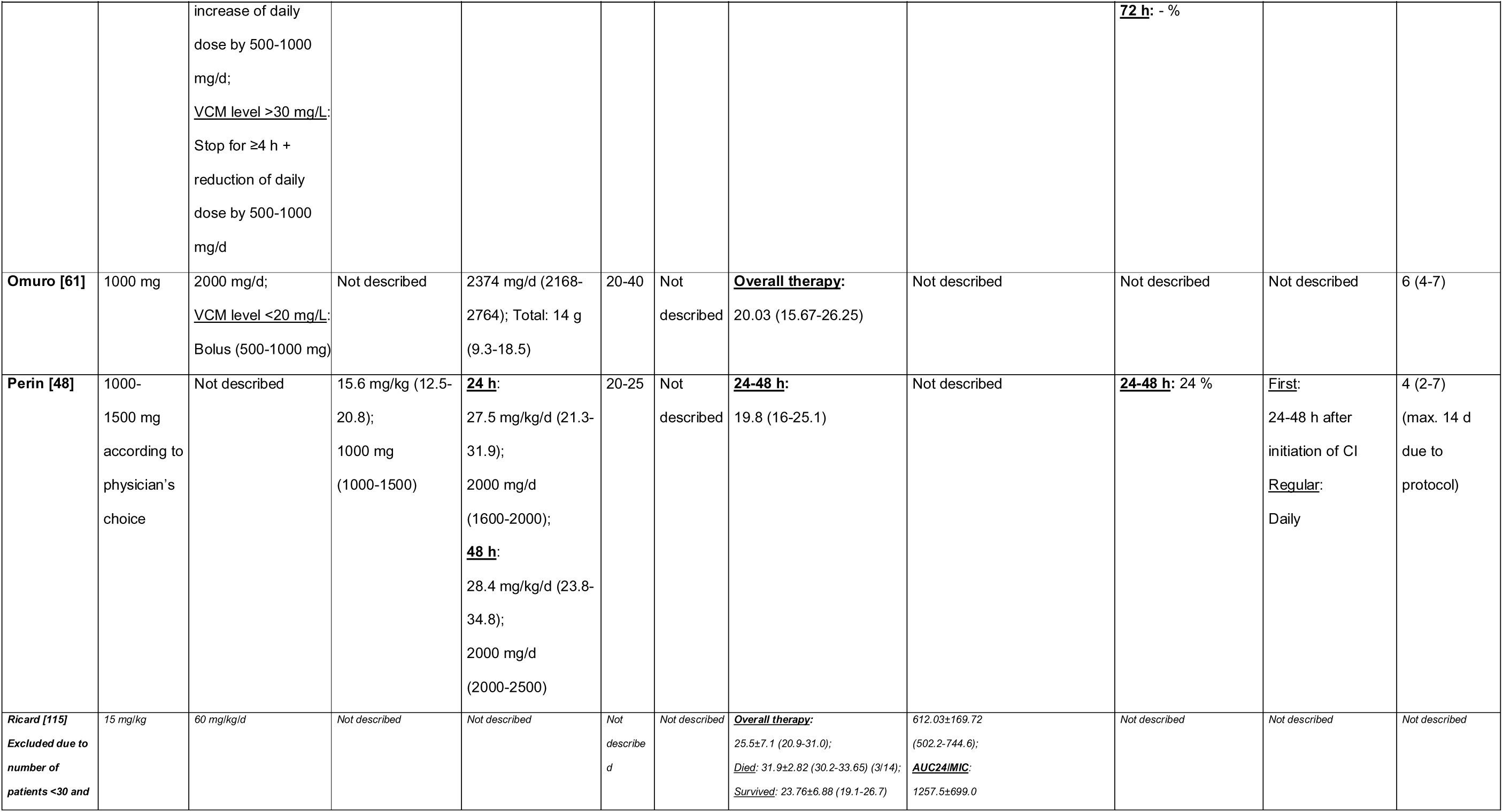
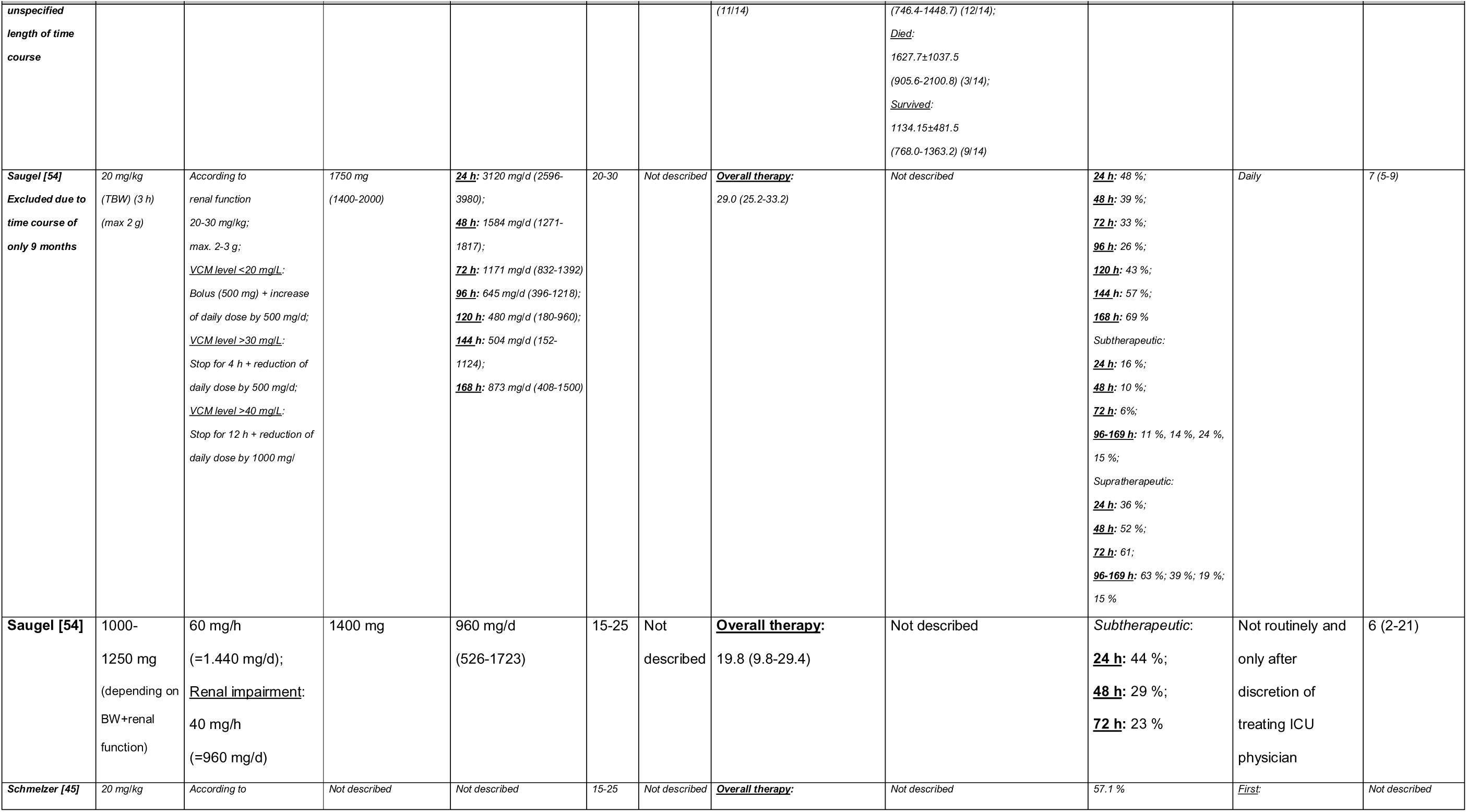
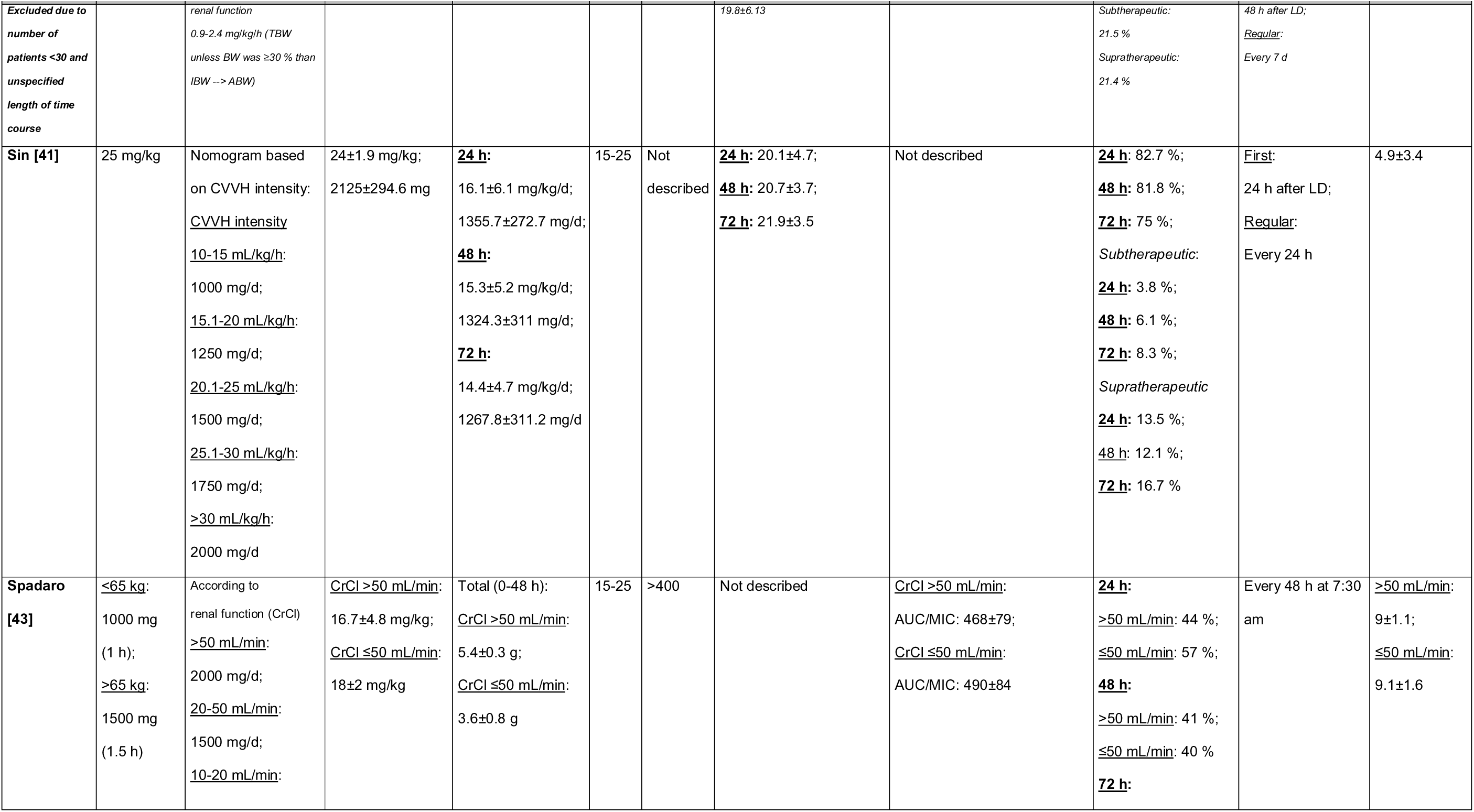
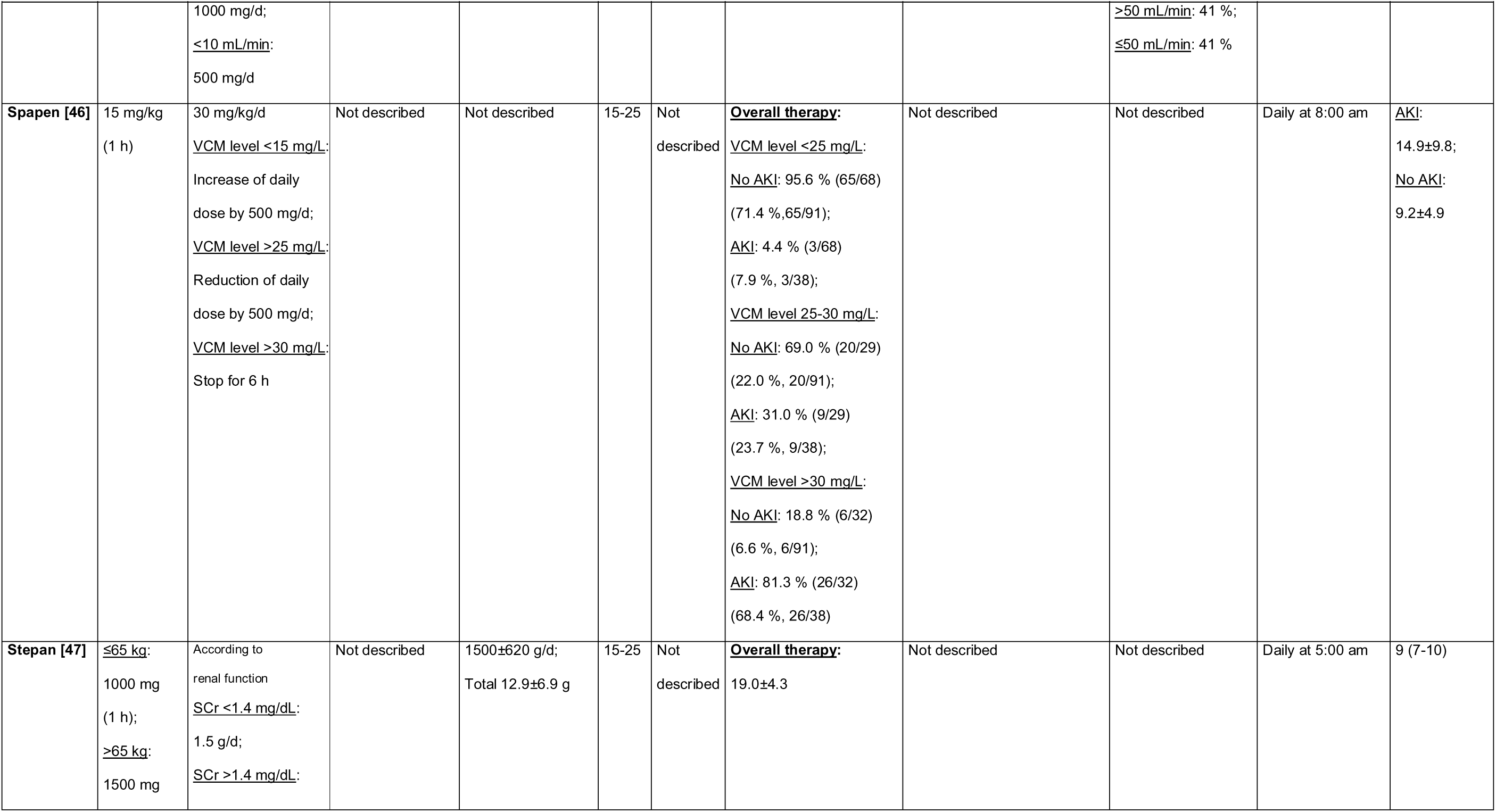
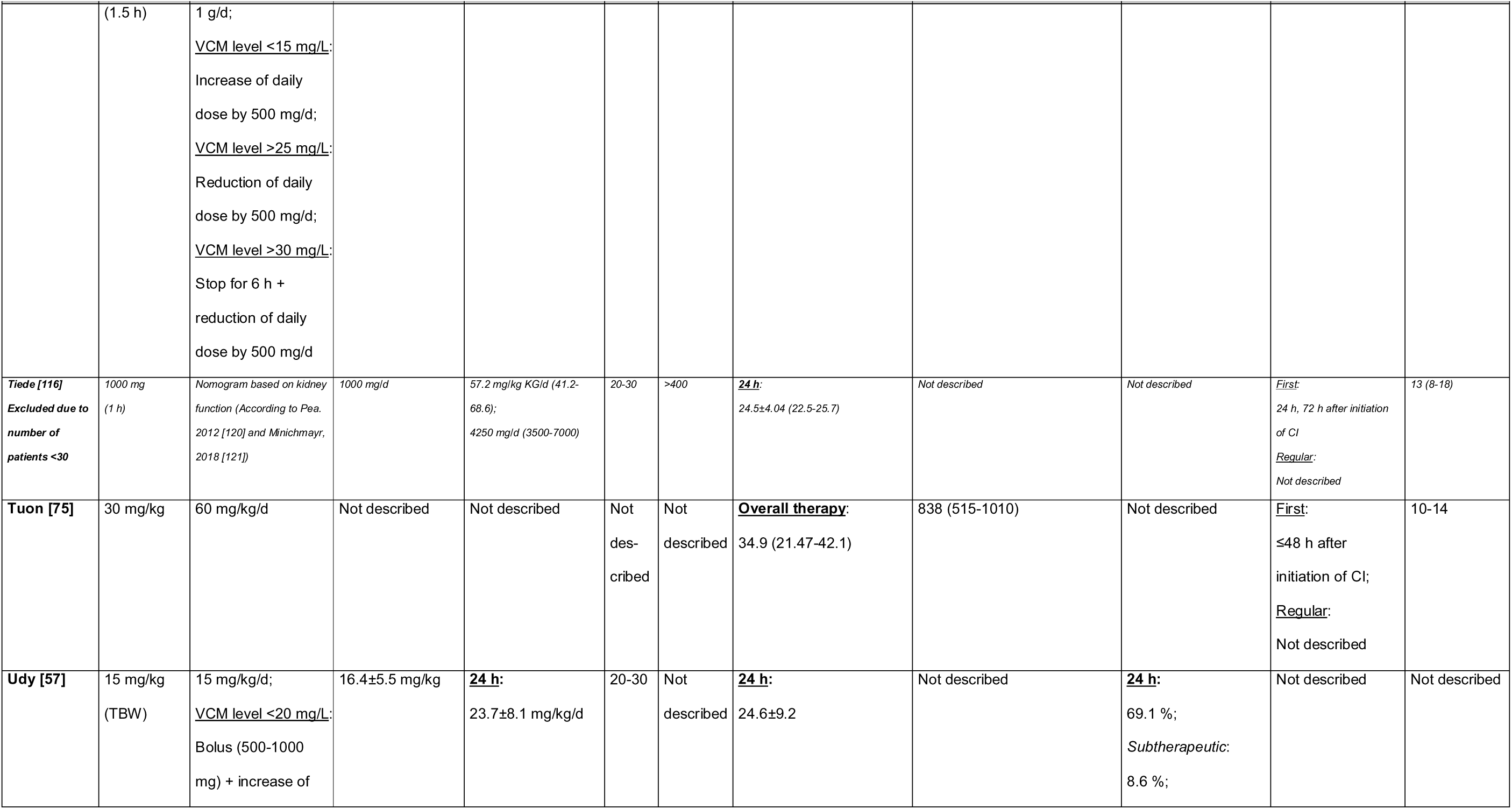
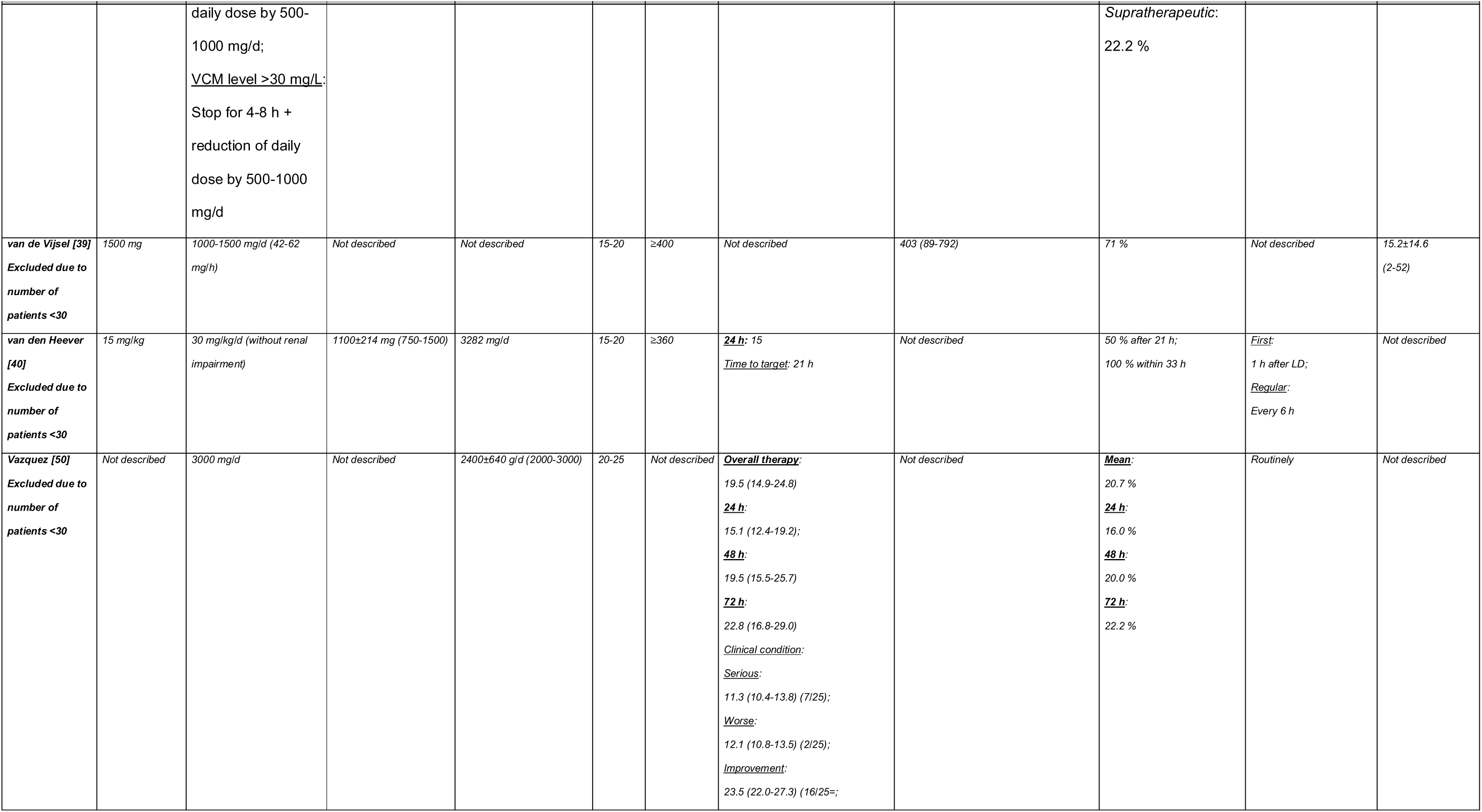
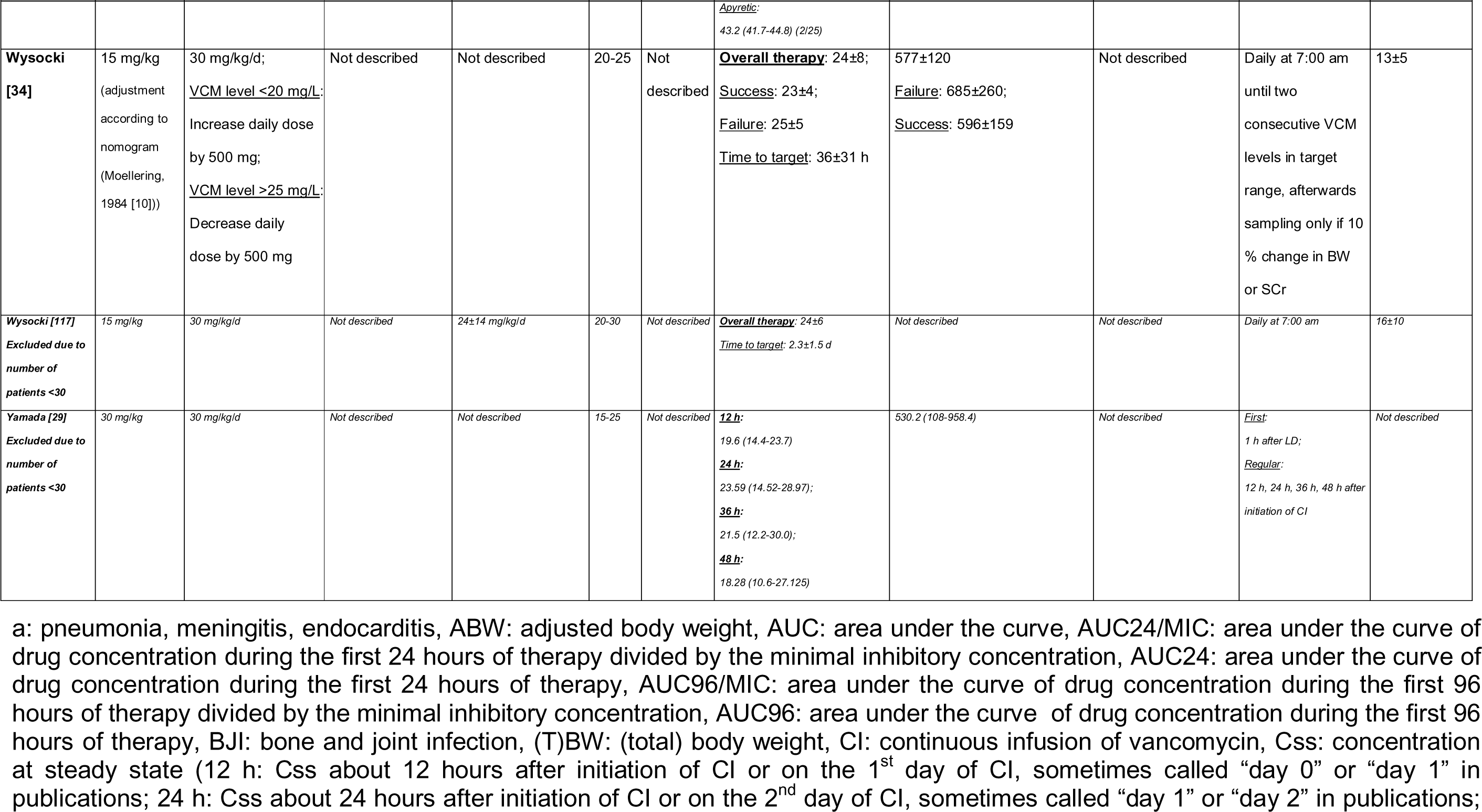
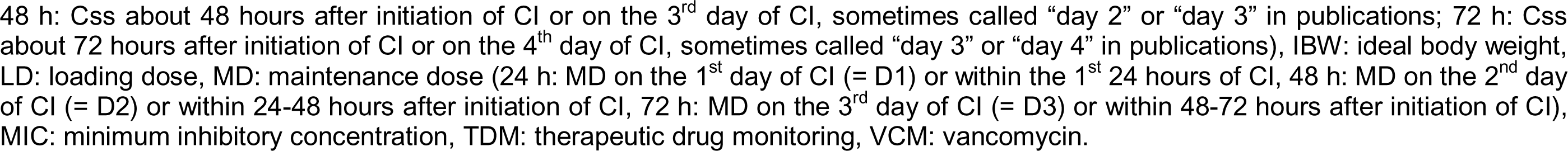
Overview of the characteristics of the continuous infusion of vancomycin. Thirty-eight studies were identified in the initial full-text selection process that presented data of steady-state serum concentrations of continuously administered vancomycin (CI) and efficacy (clinical or microbiological success (e.g. cure) or failure (e.g. mortality)) or safety (e.g. acute renal failure). Only twenty-one studies made it into the final analysis. Studies excluded from the final selection because of an increased risk of bias (e.g. unspecified or time course of less than 12 months or a CI study population of less than 30 patients) are written in italics.

| Reference | Planned |  | Measured |  | Target definition |  | Measured |  |  | TMD | Duration of Therapy [days] |
| --- | --- | --- | --- | --- | --- | --- | --- | --- | --- | --- | --- |
|  | Loading Dose (LD) | Maintenance Dose (MD) | Loading Dose (LD) | Maintenance Dose (MD) | Css [mg/L] | AUC24/ MIC | Css [mg/L] (mean/median) | AUC24 [mg*h/L] (mean/median) | Target attainment [%] |  |  |
| Akers [49] | Not described | 3000 mg/d | Not described | 29.5±11.8 mg/kg/d;<br>2500±720 mg/d;<br>*CVVH*:<br>Initially 31.7±10.4 mg/kg/d | 20-25 | ≥400 | **Overall therapy:**<br>20.0±3.8;<br>*Gram-positive bacteremia:*<br>19.3±3.2;<br>*Sepsis without proven gram-positive bacteremia:*<br>21.2±4.2;<br>*Pneumonia:*<br>22.0±3.9 | Lack of postinfusion peak<br>data prevented actual calculation of AUC | *Non-CVVH* (n=68):<br>30.7±19.9 %;<br>*Subtherapeutic:*<br>50.0±26.7 %;<br>*Supratherapeutic:*<br>19.3±21.7 %;<br>*CVVH* (n=22):<br>30.0±20.3 %;<br>*Subtherapeutic:*<br>45.1±29.6 %;<br>*Supratherapeutic:*<br>24.9±24.1 % | *First:*<br>24 h after initiation of CI<br>*Regular:*<br>Not described | *Gram-positive bacteremia:*<br>12.4±11.8;<br>*Sepsis without proven gram-positive bacteremia:*<br>8.7±5.9;<br>*Pneumonia:*<br>13.5±11.1 |
| Albanèse [60]<br>*Excluded due to number of patients <30 and unspecified length of time course* | 15 mg/kg (2 h) | 50-60 mg/kg/d | Not described | 62±17 mg/kg/d | 20-30 | Not described | **Overall therapy:**<br>32±8<br>(range:<br>*Meningitis:* 11.6-46.8;<br>*Without meningitis:* 14.6-36.1) | Not described | Not described | *First:*<br>1st day: 2:00, 2:30, 3:00, 4:00, 5:00, 6:00, 8:00 pm, midnight;<br>2nd day: 6:00 am; 12:00 noon, 2:00 pm, midnight;<br>3rd day: 6:00 am, 2:00 | 13±7 |
|  | Loading Dose (LD) | Maintenance Dose (MD) | Loading Dose (LD) | Maintenance Dose (MD) | Css [mg/L] | AUC24/MIC | Css [mg/L] (mean/median) | AUC24 [mg*h/L] (mean/median) | Target attainment [%] |  |  |
|  |  |  |  |  |  |  |  |  |  | pm;<br>Regular:<br>Daily at 6:00 am |  |
| **Baptista [53]** | <u>≤70 kg</u>:<br>1000 mg;<br><u>>70 kg</u>:<br>1500 mg | <u>A</u>: 30 mg/kg/d;<br><u>B</u>: nomogram | <u>A</u>: 14.3 mg/kg<br>(12.8-17.6);<br>1000 mg/d<br>(1000-1500);<br><u>B</u>: 18.8 mg/kg<br>(16.7-21.4);<br>1500 mg/d<br>(1000-1500) | <u>A</u>: 1920 mg/d<br>(1512-2400);<br><u>B</u>: 2072 mg/d<br>(1750-2622) | 20-30 | Not described | <u>24 h</u>:<br><u>A</u>: 20.6 (16.7-26);<br><u>B</u>: 24.5 (22.2-27.4) | Not described | <u>A</u>: 51 %;<br><u>B</u>: 84 % | Daily at 7:00 am | Not described |
| **Beumier [55]** | 35 mg/kg<br>(4 h)<br>(Adapted from Roberts, 2011. [118]) | 14 mg/kg/d | 2750 mg<br>(2250-3150) | 1100 mg/d<br>(975-1270) | 20-30 | >400 | <u>24 h</u>:<br>23 (19-31) | 652 (596-789) | <u>12 h</u>: 69 %;<br><u>24 h</u>: 63 %;<br>*Subtherapeutic*:<br><u>12 h</u>: 3 %;<br><u>24 h</u>: 19 %;<br>*Supratherapeutic*:<br><u>12 h</u>: 28 %;<br><u>24 h</u>: 19 % | <u>First</u>:<br>End of LD (T1);<br>12 h after initiation of CI (T2);<br>24 h after initiation of CI (T3);<br><u>Regular</u>: daily at 8:00 am | 3 (2-7) |
| **Carricajo [62]**<br>*Excluded due to number of patients <30 and unspecified length of time course* | 30 mg/kg<br>(1 h) | 30 mg/kg/d | Not described | Not described | 25-30 | ≥350 | <u>24 h</u>:<br>*All*:<br>21.33±7.64 (14.1-25.4);<br>*<120 mL/min*:<br>25.8±6.37 (21.9-27.3);<br>*≥120 mL/min*:<br>13.45±1.98 (12.6-14.7) | <u>AUC24/MIC</u>:<br>*All*:<br>548.3±300.3 (325.3-568.9);<br>*<120 mL/min*:<br>679.25±357.9 (371.8-667.0);<br>*≥120 mL/min*:<br>286.5±93.75 (264.8-345.3) | <u>Css</u>:<br>*All*: 27.3 %;<br>*<120 mL/min*: 42.9 %;<br>*≥120 mL/min*: 0 %;<br>*Subtherapeutic*:<br>*All*: 59.1 %;<br>*<120 mL/min*: 35.7 %;<br>*≥120 mL/min*: 100 %; | <u>First</u>:<br>24 h after initiation of CI;<br><u>Regular</u>:<br>Not described | Not described |
|  | Loading Dose (LD) | Maintenance Dose (MD) | Loading Dose (LD) | Maintenance Dose (MD) | Css [mg/L] | AUC24/ MIC | Css [mg/L] (mean/median) | AUC24 [mg*h/L] (mean/median) | Target attainment [%] |  |  |
|  |  |  |  |  |  |  |  |  | *Supratherapeutic:*<br><u>All:</u> 13.6 %;<br>*<120 mL/min:* 21.4 %;<br>*≥120 mL/min:* 0 %;<br><u>AUC/MIC:</u><br><u>All:</u> 66.7 %;<br>*<120 mL/min:* 87.5 %;<br>*≥120 mL/min:* 25 % |  |  |
| Cianferoni [56] | 15 mg/kg (1-1.5 h) | According to renal function<br>20-30 mg/kg/d (TBW);<br><u>VCM level <20 mg/L:</u><br>Bolus (500 mg) + increase of daily dose by 500-1000 mg/d;<br><u>VCM level >30 mg/L:</u><br>Stop for ≥4 h + reduction of daily dose by 500-1000 mg/d | 1000 mg (1000-1500) | <u>All:</u> 2000 mg/d (1697-2500);<br><u>No AKI:</u> 2000 mg/d (1800-2642);<br><u>AKI:</u> 1940 mg/d (1431-2212);<br><u>(Early AKI:</u><br>1747 mg/d<br>1206-2000);<br><u>Late AKI:</u><br>2050 mg/d (1714-2691))<br><u>Total dose</u><br><u>All:</u><br>8.1 g (6-13.5);<br><u>No AKI:</u><br>9.45 g (7-14);<br><u>AKI:</u><br>11.5 g (7.5-16.9) | 20-30 | Not described | <u>Overall therapy:</u><br><u>All:</u> 22.3±7.0;<br><u>No AKI:</u> 21.2±6.8;<br><u>AKI:</u> 25.8±6.6;<br>(Early AKI: 27.2±5.1;<br>Late AKI: 24.1±7.8)<br><u>24 h:</u><br><u>All:</u> 19.7±8.5;<br><u>No AKI:</u> 18.7±7;<br><u>AKI:</u> 22.5±9.6;<br>(Early AKI: 24.5±7.8;<br>Late AKI: 20.2±11.1) | Not described | Not described | Daily at 8:00 am | <u>All:</u> 4 (3-6);<br><u>No AKI:</u><br>4 (3-6);<br><u>AKI:</u><br>5 (3-9);<br>(Early AKI:<br>5 (3-7);<br><u>Late AKI:</u><br>5 (4-9)) |
|  | Loading Dose (LD) | Maintenance Dose (MD) | Loading Dose (LD) | Maintenance Dose (MD) | Css [mg/L] | AUC24/ MIC | Css [mg/L] (mean/median) | AUC24 [mg*h/L] (mean/median) | Target attainment [%] |  |  |
| **Covajes [58]** | 15 mg/kg (TBW) | 20-30 mg/kg/d;<br><u>VCM level <20 mg/L:</u><br>Bolus (500-1000 mg)<br>+ increase of daily dose by 500-1000 mg/d;<br><u>VCM level >30 mg/L:</u><br>Stop for 4-8 h + reduction of daily dose by 500-1000 mg/d | 16.4±6.4 mg/kg | **24 h:**<br>23.5 ± 8.1 mg/kg/d;<br>**48 h:**<br>23.2 ± 7.4 mg/kg/d;<br>**72 h:**<br>23.3 ± 11.0 mg/kg/d | 20-30 | Not described | **24 h:** 24.7±9.0;<br>**48 h:** 26.0±8.1;<br>**72 h:** 27.7±9.3 | Not described | **24 h:** 51 %;<br>**48 h:** 56 % ;<br>**72 h:** 67 %;<br>*Subtherapeutic:*<br>**24 h:** 29 %;<br>**48 h:** 22 % ;<br>**72 h:** 8 %;<br>*Supratherapeutic:*<br>**24 h:** 20 %;<br>**48 h:** 21 % ;<br>**72 h:** 26 | Daily at 8:00 am | 4 (3-7) |
| **Cristallini [52]** | 35 mg/kg (4 h) | According to renal function<br><u>>150 mL/min:</u><br>45 mg/kg/d;<br><u>120-150 mL/min:</u><br>40 mg/kg/d;<br><u>80-119 mL/min:</u><br>30 mg/kg/d;<br><u>50-69 mL/min:</u><br>25 mg/kg/d;<br><u>25-50 mL/min:</u><br>14 mg/kg/d;<br><u><25 (anuria):</u> | 2650 mg | 2112 mg/d | 20-30 | Not described | **12 h:** 25 (21-32);<br>**24 h:** 22 (19-28);<br>**48 h:** 26 (22-30) | 771 (664-905) | **12 h:** 56 %;<br>**24 h:** 54 %;<br>**48 h:** 73 % | <u>First:</u><br>End of LD;<br>12 h, 24 h, 48 h after initiation of CI<br><u>Regular:</u><br>Daily at 8:00 am | Not described |
|  | Loading Dose (LD) | Maintenance Dose (MD) | Loading Dose (LD) | Maintenance Dose (MD) | Css [mg/L] | AUC24/MIC | Css [mg/L] (mean/median) | AUC24 [mg*h/L] (mean/median) | Target attainment [%] |  |  |
|  |  | 7 mg/kg/d;<br><u>VCM level <20 mg/L:</u><br>Bolus (500-1000 mg) + increase of daily dose by 25 %/d;<br><u>VCM level >30 mg/L:</u><br>Stop for 4 h + reduction of daily dose 25 %/d |  |  |  |  |  |  |  |  |  |
| *Di Filippo [63]*<br>*Excluded due to number of patients <30 and time course of only 7 months* | 500 mg<br>(10 min) | 2000 mg/d | Not described | Not described | 30-40 | Not described | <u>24 h:</u> 13.88±5.7;<br><u>48 h:</u> 17.76±7.6;<br><u>72 h:</u> 24.3±3.9 | Not described | Not described | <u>First:</u><br>2 h, 48 h, 96 h after initiation of CI;<br><u>Regular:</u><br>Not described | 6.14±1.37 |
| *Duszynska [31]*<br>*Excluded due to number of patients <30* | 500 mg | 30 mg/kg/d (TBW);<br><u>VCM level <15 mg/L:</u><br>Bolus (500 mg) + increase of daily dose by 25 %/d;<br><u>VCM level >20 mg/L:</u><br>Reduction of daily dose by 25 %/d | 500 mg | 29.6±5.5 mg/kg/d;<br>2219±476 mg/d;<br><u>Total:</u> 17.7±5.2 g | 15-20 | ≥400 | <u>Time to target:</u><br>72 h (24-120) | 316 (123-577);<br>MIC: 2 (1-2);<br><u>AUC24/MIC:</u><br><u>All:</u> 195 (133-343);<br><u>Clinical cure:</u> 194 (134-389);<br><u>Microbiological eradication:</u><br>162 (116-389)<br><u>AUC96/MIC:</u><br><u>All:</u> 1112 (650-1414) | <u>Css:</u> 71.4 %<br>Subtherapeutic: 19 %;<br>Supratherapeutic: 10 %<br><u>AUC24/MIC:</u> 23.8 %;<br><u>AUC96/MIC:</u> 24 % | <u>First:</u><br>Immediately + 30 min after LD;<br>24 h, 48 h, 96 h after initiation of CI;<br><u>Regular:</u><br>Not described | 7 (7-8) |
| *Eldemiry [38]*<br>*Excluded due to number of patients <30* | none | Rate of infusion (g/d) =<br>[0.0205 CrCl + 3.47] * (Css)<br>* (24/1000) | Not described | <u>Total:</u> 9.93±2.21 g | 15-20 | Not described | <u>Overall therapy:</u><br>18.13±14.15 | Not described | 40 %;<br>Subtherapeutic: 30 %;<br>Supratherapeutic: 30 % | Not described | 5.05±0.99 |
|  | Loading Dose (LD) | Maintenance Dose (MD) | Loading Dose (LD) | Maintenance Dose (MD) | Css [mg/L] | AUC24/ MIC | Css [mg/L] (mean/median) | AUC24 [mg*h/L] (mean/median) | Target attainment [%] |  |  |
| Hanrahan [76] | Not described | Not described | Not described | 1700 mg/d (1200-2100);<br><u>Total:</u><br>9.0 g (6.0-14.4) | Not described | Not described | <u>Overall therapy:</u><br>18.4 (15.6-21.2) | Not described | Not described | <u>First:</u><br>≥18 h after dose change;<br><u>Regular:</u><br>Not described | 5.3 (3.4-10.3) |
| Hong [26]<br>Excluded due to time course of only 9 months | 20 mg/kg (1 g/h) | According to renal function<br>≥90 mL/min:<br>40 mg/kg/d;<br>50-90 mL/min:<br>30 mg/kg/d;<br>20-50 mL/min:<br>15 mg/kg/d;<br>(Dose adjustment via ratio method (current total daily dose x desired Css = new total daily dose x current Css) or pharmacokinetic equation: $Css = (dose/24 h)/clearance$) | 19.9±3 mg/kg | Achieving goal:<br>33±10.6 mg/kg/d;<br>2572±784 mg/d | 20-30 | >400 | <u>24 h:</u> 28±9.2<br><u>Time to target:</u> 1.96 d;<br><u>Time within target:</u> 55 % | 671.5±219.9 | <u>24 h:</u> 40 %;<br>Subtherapeutic:<br>21.5 %<br>Supratherapeutic:<br>38.5 %<br><u>AUC24/MIC (S. aureus, 11/15):</u><br>MIC=0.5: 100 % (2/2);<br>MIC=1: 88.9 % (8/9);<br>MIC=2: 0 % (0/0)<br><u>Population-based:</u><br>MIC=0.5: 100 % (65/65);<br>MIC=1: 92.3 % (60/65);<br>MIC=2: 27.7 % (18/65) | <u>First:</u><br>24 h after initiation of CI;<br><u>CrCl <50 mL/min:</u> 36-48 h after initiation of CI;<br><u>Regular:</u><br>not described | 10.4±7.8 |
| Hutschala [33] | 20 mg/kg (1 h) | 36 mg/kg/d (0.025 mg/kg/min) | Not described | 1935±688 mg/d (352-3411) | 20-25 | Not described | <u>Overall therapy:</u><br>25.0±4.0 (7-30);<br><u>Time to target:</u> 16±8 h | 529±98 | Not described | Daily at 7:00 am | 9±6 (3-19);<br><u>Time between surgery and start of therapy:</u><br>4.9±1.8 |
| Lin [32]<br>Excluded due to number of | 25 mg/kg | 1500 mg/d (60 mg/h) | 26.65±3.06 mg/kg;<br>2232.14±385.61 mg | <u>Overall therapy:</u><br>15.66±6.26 mg/kg/d;<br><u>24 h:</u> 1329.85±335.61 | 15-25 | Not described | <u>24 h:</u> 20.35±2.78;<br><u>48 h:</u> 20.8±2.66;<br><u>72 h:</u> 19.25±2.01; | Not described | <u>24 h:</u> 100 % | <u>First:</u><br>24 h after initiation of CI; | 6.57±3.25 |
|  | Loading Dose (LD) | Maintenance Dose (MD) | Loading Dose (LD) | Maintenance Dose (MD) | Css [mg/L] | AUC24/ MIC | Css [mg/L] (mean/median) | AUC24 [mg*h/L] (mean/median) | Target attainment [%] |  |  |
| patients <30 |  |  |  | mg/d;<br><u>48 h</u>: 1339.28±348.17 mg/d;<br><u>72 h</u>: 1428.57±301.09 mg/d;<br><u>Total dose prior to first 24 h level</u>: 3625±497.5 mg |  |  | <u>Time to target</u>: 24±0.56 h |  |  | <u>Regular</u>:<br>Daily, but no further monitoring, when two consecutive serum concentrations within target range and no change in CVVH flow rate |  |
| Lin [42] | 25-30 mg/kg | According to renal function<br>>60 mL/min:<br>2000 mg/d;<br>max. 4.25 g/d;<br><u>Concomitant CVVH</u>:<br>Initial dose 1500 mg/d;<br><u>VCM level <15 mg/L</u>:<br>Bolus (500-1000 mg) + proportional increase of daily dose;<br><u>VCM level >25 mg/L</u>:<br>Stop for 4-6 h + proportional reduction of daily dose | <u>Obese</u>:<br>24.8±2.05 mg/kg;<br>2894±525 mg;<br><u>Non-obese</u>:<br>25.6±1.66 mg/kg;<br>1923±345 mg;<br><u>CVVH</u>:<br><u>Obese</u>:<br>25.3±0.744 mg/kg;<br>2722±441 mg;<br><u>Non-obese</u>:<br>25.4±1.49 mg/kg;<br>1833±280 mg<br><u>Preserved renal function</u>:<br><u>Obese</u>: 24.6±2.47 | <u>Obese</u>:<br>24.8±12.7 mg/kg/d;<br>2885±1450 mg/d;<br><u>Non-obese</u>:<br>41.9±20 mg/kg/d;<br>3039±1313 mg/d;<br><u>CVVH</u>:<br><u>Obese</u>: 13.3±4.73 mg/kg/d;<br>1389±39 7 mg/d;<br><u>Non-obese</u>:<br>21.7±8.33 mg/kg/d;<br>1500±375 mg/d;<br><u>Preserved renal function</u>:<br><u>Obese</u>: 30.8±11.3 mg/kg/d; | 15-25 | Not described | <u>24 h</u>:<br><u>All</u>: 20.32±3.72;<br><u>Obese</u>: 20.3±3.81;<br><u>Non-obese</u>: 20.03±3.79;<br><u>CVVH</u>:<br><u>Obese</u>: 19.8±3.05;<br><u>Non-obese</u>: 21.6±3.02;<br><u>Preserved renal function</u>:<br><u>Obese</u>: 20.6±4.21;<br><u>Non-obese</u>: 19.6±4.41 | <u>Obese</u>: 488±91.5;<br><u>Non-obese</u>: 480.9±91.04;<br><u>CVVH</u>:<br><u>Obese</u>: 474±73.2;<br><u>Non-obese</u>: 517±48.6;<br><u>Preserved renal function</u>:<br><u>Obese</u>: 496±101;<br><u>Non-obese</u>: 471±106 | <u>24 h</u>:<br><u>Obese</u>: 92.3 %;<br><u>Non-obese</u>: 88.5 %<br><u>CVVH</u>:<br><u>Obese</u>: 100 %;<br><u>Non-obese</u>: 100 % | <u>First</u>:<br>24 h after initiation of CI;<br><u>Regular</u>:<br>Every 24 h until 2 serum concentrations within target range | <u>Obese</u>:<br>6.88±3.02;<br><u>Non-obese</u>:<br>6.5±3.06 |
|  | Loading Dose (LD) | Maintenance Dose (MD) | Loading Dose (LD) | Maintenance Dose (MD) | Css [mg/L] | AUC24/ MIC | Css [mg/L] (mean/median) | AUC24 [mg*h/L] (mean/median) | Target attainment [%] |  |  |
|  |  |  | mg/kg;<br>2985±555 mg;<br><u>Non-obese:</u><br>25.9±1.88 mg/kg;<br>1941±370 mg | 3676±1131 mg/d;<br><u>Non-obese:</u><br>53.4±13.7 mg/kg/d;<br>3882±674 mg/d |  |  |  |  |  |  |  |
| **Ma [37]**<br>*Excluded due to inclusion of ICU and non-ICU patients in analysis* | *Not described* | *Not described* | *Not described* | *Initial: 2047±466 mg/d (1000-3750);*<br>*Final: 2098±766 mg/d (600-4000)* | *15-20;*<br>*BJI: 12-20* | *Not described* | *Initial: 13±5 (2-27);*<br>*Final: 16±3 (7-46)* | *402±118;*<br>*Nephrotoxic risk: 535±233;*<br>*No nephrotoxic risk: 360±116* | *78 %* | *At steady state* | *46±31* |
| **Mohammedi [74]** | <u>A</u>: 500 mg;<br><u>B</u>: 15 mg/kg (1 h) | 0.227 * CrCl + 5.67 (according to Rodvold 1988 [119]) | <u>A</u>: 500 mg;<br>7.54±1.52 mg/kg;<br><u>B</u>: 1147±317 mg;<br>15 mg/kg | Not described | 20 | Not described | <u>24 h</u>:<br><u>A</u>: 14.9±5.4;<br><u>B</u>: 18.5±6.4 | Not described | Not described | <u>First</u>:<br>1 h after end of LD;<br><u>Regular</u>:<br>Not described | Not described |
| **Ocampos-Martinez [59]** | 15 mg/kg (1-1.5 h) | According to renal function<br><u>≥80 mL/min</u>:<br>30 mg/kg/d;<br><u>50-80 mL/min</u>:<br>20-30 mg/kg/d;<br><u>≤50 mL/min</u>:<br>10-20 mg/kg/d;<br><u>VCM level <20 mg/L</u>:<br>Bolus (500 mg) + | 16.2±5.4 mg/kg;<br>1199±366 mg | <u>24 h</u>:<br>26.8±8.1 mg/kg/d;<br>1975±507 mg/d;<br><u>48 h</u>:<br>28.7±10.1 mg/kg/d;<br>2103±636 mg/d;<br><u>72 h</u>:<br>29.7±13.9 mg/kg/d;<br>2232±688 mg/d | 20-30 | Not described | <u>24 h</u>: 20.4±8.9;<br><u>48 h</u>: 24.1±9.1;<br><u>72 h</u>: 25.5±10.1;<br><u>Time to target</u>: 2 d (1-3) | Not described | <u>24 h</u>: 47 %;<br><u>48 h</u>: 67 %;<br><u>72 h</u>: 86 %;<br><u>Subtherapeutic</u>:<br><u>24 h</u>: 42 %;<br><u>48 h</u>: 14 %;<br><u>72 h</u>: - %;<br><u>Supratherapeutic</u>:<br><u>24 h</u>: 11 %;<br><u>48 h</u>: 19 %; | <u>First</u>:<br>≥16 h after initiation of CI;<br><u>Regular</u>: daily at 8:00 am | 4 (3-7) |
|  | Loading Dose (LD) | Maintenance Dose (MD) | Loading Dose (LD) | Maintenance Dose (MD) | Css [mg/L] | AUC24/MIC | Css [mg/L] (mean/median) | AUC24 [mg*h/L] (mean/median) | Target attainment [%] |  |  |
|  |  | increase of daily dose by 500-1000 mg/d;<br><u>VCM level >30 mg/L:</u><br>Stop for ≥4 h + reduction of daily dose by 500-1000 mg/d |  |  |  |  |  |  | <u>72 h:</u> - % |  |  |
| Omuro [61] | 1000 mg | 2000 mg/d;<br><u>VCM level <20 mg/L:</u><br>Bolus (500-1000 mg) | Not described | 2374 mg/d (2168-2764); Total: 14 g (9.3-18.5) | 20-40 | Not described | <u>Overall therapy:</u><br>20.03 (15.67-26.25) | Not described | Not described | Not described | 6 (4-7) |
| Perin [48] | 1000-1500 mg according to physician's choice | Not described | 15.6 mg/kg (12.5-20.8);<br>1000 mg (1000-1500) | <u>24 h:</u><br>27.5 mg/kg/d (21.3-31.9);<br>2000 mg/d (1600-2000);<br><u>48 h:</u><br>28.4 mg/kg/d (23.8-34.8);<br>2000 mg/d (2000-2500) | 20-25 | Not described | <u>24-48 h:</u><br>19.8 (16-25.1) | Not described | <u>24-48 h:</u> 24 % | <u>First:</u><br>24-48 h after initiation of CI<br><u>Regular:</u><br>Daily | 4 (2-7)<br>(max. 14 d due to protocol) |
| Ricard [115]<br>Excluded due to number of patients <30 and | 15 mg/kg | 60 mg/kg/d | Not described | Not described | Not described | Not described | <u>Overall therapy:</u><br>25.5±7.1 (20.9-31.0);<br><u>Died:</u> 31.9±2.82 (30.2-33.65) (3/14);<br><u>Survived:</u> 23.76±6.88 (19.1-26.7) | 612.03±169.72 (502.2-744.6);<br><u>AUC24/MIC:</u><br>1257.5±699.0 | Not described | Not described | Not described |
|  | Loading Dose (LD) | Maintenance Dose (MD) | Loading Dose (LD) | Maintenance Dose (MD) | Css [mg/L] | AUC24/ MIC | Css [mg/L] (mean/median) | AUC24 [mg*h/L] (mean/median) | Target attainment [%] |  |  |
| *unspecified length of time course* |  |  |  |  |  |  | (11/14) | (746.4-1448.7) (12/14);<br><u>Died:</u><br>1627.7±1037.5<br>(905.6-2100.8) (3/14);<br><u>Survived:</u><br>1134.15±481.5<br>(768.0-1363.2) (9/14) |  |  |  |
| **Saugel [54]**<br>*Excluded due to time course of only 9 months* | 20 mg/kg (TBW) (3 h) (max 2 g) | According to renal function<br>20-30 mg/kg;<br>max. 2-3 g;<br><u>VCM level <20 mg/L:</u><br>Bolus (500 mg) + increase of daily dose by 500 mg/d;<br><u>VCM level >30 mg/L:</u><br>Stop for 4 h + reduction of daily dose by 500 mg/d;<br><u>VCM level >40 mg/L:</u><br>Stop for 12 h + reduction of daily dose by 1000 mg/ | 1750 mg (1400-2000) | **24 h:** 3120 mg/d (2596-3980);<br>**48 h:** 1584 mg/d (1271-1817);<br>**72 h:** 1171 mg/d (832-1392)<br>**96 h:** 645 mg/d (396-1218);<br>**120 h:** 480 mg/d (180-960);<br>**144 h:** 504 mg/d (152-1124);<br>**168 h:** 873 mg/d (408-1500) | 20-30 | Not described | **Overall therapy:**<br>29.0 (25.2-33.2) | Not described | **24 h:** 48 %;<br>**48 h:** 39 %;<br>**72 h:** 33 %;<br>**96 h:** 26 %;<br>**120 h:** 43 %;<br>**144 h:** 57 %;<br>**168 h:** 69 %<br>Subtherapeutic:<br>**24 h:** 16 %;<br>**48 h:** 10 %;<br>**72 h:** 6 %;<br>**96-168 h:** 11 %, 14 %, 24 %, 15 %;<br>Supratherapeutic:<br>**24 h:** 36 %;<br>**48 h:** 52 %;<br>**72 h:** 61;<br>**96-168 h:** 63 %, 39 %, 19 %, 15 % | Daily | 7 (5-9) |
| **Saugel [54]** | 1000-1250 mg (depending on BW+renal function) | 60 mg/h (=1.440 mg/d);<br><u>Renal impairment:</u><br>40 mg/h (=960 mg/d) | 1400 mg | 960 mg/d (526-1723) | 15-25 | Not described | **Overall therapy:**<br>19.8 (9.8-29.4) | Not described | **Subtherapeutic:**<br>**24 h:** 44 %;<br>**48 h:** 29 %;<br>**72 h:** 23 % | Not routinely and only after discretion of treating ICU physician | 6 (2-21) |
| **Schmelzer [45]** | 20 mg/kg | According to | Not described | Not described | 15-25 | Not described | **Overall therapy:** | Not described | 57.1 % | <u>First:</u> | Not described |
|  | Loading Dose (LD) | Maintenance Dose (MD) | Loading Dose (LD) | Maintenance Dose (MD) | Css [mg/L] | AUC24/ MIC | Css [mg/L] (mean/median) | AUC24 [mg*h/L] (mean/median) | Target attainment [%] |  |  |
| Excluded due to number of patients <30 and unspecified length of time course |  | renal function<br>0.9-2.4 mg/kg/h (TBW unless BW was ≥30 % than IBW --> ABW) |  |  |  |  | 19.8±6.13 |  | Subtherapeutic:<br>21.5 %<br>Supratherapeutic:<br>21.4 % | 48 h after LD;<br>Regular:<br>Every 7 d |  |
| Sin [41] | 25 mg/kg | Nomogram based on CVVH intensity:<br><u>CVVH intensity</u><br>10-15 mL/kg/h:<br>1000 mg/d;<br><u>15.1-20 mL/kg/h:</u><br>1250 mg/d;<br><u>20.1-25 mL/kg/h:</u><br>1500 mg/d;<br><u>25.1-30 mL/kg/h:</u><br>1750 mg/d;<br><u>>30 mL/kg/h:</u><br>2000 mg/d | 24±1.9 mg/kg;<br>2125±294.6 mg | **24 h:**<br>16.1±6.1 mg/kg/d;<br>1355.7±272.7 mg/d;<br>**48 h:**<br>15.3±5.2 mg/kg/d;<br>1324.3±311 mg/d;<br>**72 h:**<br>14.4±4.7 mg/kg/d;<br>1267.8±311.2 mg/d | 15-25 | Not described | **24 h:** 20.1±4.7;<br>**48 h:** 20.7±3.7;<br>**72 h:** 21.9±3.5 | Not described | **24 h:** 82.7 %;<br>**48 h:** 81.8 %;<br>**72 h:** 75 %;<br>Subtherapeutic:<br>**24 h:** 3.8 %;<br>**48 h:** 6.1 %;<br>**72 h:** 8.3 %;<br>Supratherapeutic<br>**24 h:** 13.5 %;<br>**48 h:** 12.1 %;<br>**72 h:** 16.7 % | First:<br>24 h after LD;<br>Regular:<br>Every 24 h | 4.9±3.4 |
| Spadaro [43] | <65 kg:<br>1000 mg (1 h);<br>>65 kg:<br>1500 mg (1.5 h) | According to renal function (CrCl)<br><u>>50 mL/min:</u><br>2000 mg/d;<br><u>20-50 mL/min:</u><br>1500 mg/d;<br><u>10-20 mL/min:</u> | <u>CrCl >50 mL/min:</u><br>16.7±4.8 mg/kg;<br><u>CrCl ≤50 mL/min:</u><br>18±2 mg/kg | Total (0-48 h):<br><u>CrCl >50 mL/min:</u><br>5.4±0.3 g;<br><u>CrCl ≤50 mL/min:</u><br>3.6±0.8 g | 15-25 | >400 | Not described | <u>CrCl >50 mL/min:</u><br>AUC/MIC: 468±79;<br><u>CrCl ≤50 mL/min:</u><br>AUC/MIC: 490±84 | **24 h:**<br><u>>50 mL/min:</u> 44 %;<br><u>≤50 mL/min:</u> 57 %;<br>**48 h:**<br><u>>50 mL/min:</u> 41 %;<br><u>≤50 mL/min:</u> 40 %<br>**72 h:** | Every 48 h at 7:30 am | <u>>50 mL/min:</u><br>9±1.1;<br><u>≤50 mL/min:</u><br>9.1±1.6 |
|  | Loading Dose (LD) | Maintenance Dose (MD) | Loading Dose (LD) | Maintenance Dose (MD) | Css [mg/L] | AUC24/MIC | Css [mg/L] (mean/median) | AUC24 [mg*h/L] (mean/median) | Target attainment [%] |  |  |
|  |  | 1000 mg/d;<br><u><10 mL/min:</u><br>500 mg/d |  |  |  |  |  |  | >50 mL/min: 41 %;<br><u>≤50 mL/min:</u> 41 % |  |  |
| Spapen [46] | 15 mg/kg<br>(1 h) | 30 mg/kg/d<br><u>VCM level <15 mg/L:</u><br>Increase of daily dose by 500 mg/d;<br><u>VCM level >25 mg/L:</u><br>Reduction of daily dose by 500 mg/d;<br><u>VCM level >30 mg/L:</u><br>Stop for 6 h | Not described | Not described | 15-25 | Not described | **Overall therapy:**<br><u>VCM level <25 mg/L:</u><br><u>No AKI:</u> 95.6 % (65/68)<br>(71.4 %, 65/91);<br><u>AKI:</u> 4.4 % (3/68)<br>(7.9 %, 3/38);<br><u>VCM level 25-30 mg/L:</u><br><u>No AKI:</u> 69.0 % (20/29)<br>(22.0 %, 20/91);<br><u>AKI:</u> 31.0 % (9/29)<br>(23.7 %, 9/38);<br><u>VCM level >30 mg/L:</u><br><u>No AKI:</u> 18.8 % (6/32)<br>(6.6 %, 6/91);<br><u>AKI:</u> 81.3 % (26/32)<br>(68.4 %, 26/38) | Not described | Not described | Daily at 8:00 am | <u>AKI:</u><br>14.9±9.8;<br><u>No AKI:</u><br>9.2±4.9 |
| Stepan [47] | <u>≤65 kg:</u><br>1000 mg<br>(1 h);<br><u>>65 kg:</u><br>1500 mg | According to renal function<br><u>SCr <1.4 mg/dL:</u><br>1.5 g/d;<br><u>SCr >1.4 mg/dL:</u> | Not described | 1500±620 g/d;<br>Total 12.9±6.9 g | 15-25 | Not described | **Overall therapy:**<br>19.0±4.3 | Not described | Not described | Daily at 5:00 am | 9 (7-10) |
|  | Loading Dose (LD) | Maintenance Dose (MD) | Loading Dose (LD) | Maintenance Dose (MD) | Css [mg/L] | AUC24/ MIC | Css [mg/L] (mean/median) | AUC24 [mg*h/L] (mean/median) | Target attainment [%] |  |  |
|  | (1.5 h) | 1 g/d;<br><u>VCM level <15 mg/L:</u><br>Increase of daily dose by 500 mg/d;<br><u>VCM level >25 mg/L:</u><br>Reduction of daily dose by 500 mg/d;<br><u>VCM level >30 mg/L:</u><br>Stop for 6 h + reduction of daily dose by 500 mg/d |  |  |  |  |  |  |  |  |  |
| **Tiede [116]**<br>*Excluded due to number of patients <30* | 1000 mg (1 h) | Nomogram based on kidney function (According to Pea, 2012 [120] and Minichmayr, 2018 [121]) | 1000 mg/d | 57.2 mg/kg KG/d (41.2-68.6);<br>4250 mg/d (3500-7000) | 20-30 | >400 | <u>24 h:</u><br>24.5±4.04 (22.5-25.7) | Not described | Not described | <u>First:</u><br>24 h, 72 h after initiation of CI<br><u>Regular:</u><br>Not described | 13 (8-18) |
| **Tuon [75]** | 30 mg/kg | 60 mg/kg/d | Not described | Not described | Not described | Not described | <u>Overall therapy:</u><br>34.9 (21.47-42.1) | 838 (515-1010) | Not described | <u>First:</u><br>≤48 h after initiation of CI;<br><u>Regular:</u><br>Not described | 10-14 |
| **Udy [57]** | 15 mg/kg (TBW) | 15 mg/kg/d;<br><u>VCM level <20 mg/L:</u><br>Bolus (500-1000 mg) + increase of | 16.4±5.5 mg/kg | <u>24 h:</u><br>23.7±8.1 mg/kg/d | 20-30 | Not described | <u>24 h:</u><br>24.6±9.2 | Not described | <u>24 h:</u><br>69.1 %;<br>*Subtherapeutic:*<br>8.6 %; | Not described | Not described |
|  | Loading Dose (LD) | Maintenance Dose (MD) | Loading Dose (LD) | Maintenance Dose (MD) | Css [mg/L] | AUC24/ MIC | Css [mg/L] (mean/median) | AUC24 [mg*h/L] (mean/median) | Target attainment [%] |  |  |
|  |  | daily dose by 500-1000 mg/d;<br><u>VCM level >30 mg/L</u>;<br>Stop for 4-8 h + reduction of daily dose by 500-1000 mg/d |  |  |  |  |  |  | *Supratherapeutic:*<br>22.2 % |  |  |
| van de Vijzel [39]<br>*Excluded due to number of patients <30* | 1500 mg | 1000-1500 mg/d (42-62 mg/h) | Not described | Not described | 15-20 | ≥400 | Not described | 403 (89-792) | 71 % | Not described | 15.2±14.6 (2-52) |
| van den Heever [40]<br>*Excluded due to number of patients <30* | 15 mg/kg | 30 mg/kg/d (without renal impairment) | 1100±214 mg (750-1500) | 3282 mg/d | 15-20 | ≥360 | <u>24 h</u>: 15<br><u>Time to target</u>: 21 h | Not described | 50 % after 21 h;<br>100 % within 33 h | <u>First</u>:<br>1 h after LD;<br><u>Regular</u>:<br>Every 6 h | Not described |
| Vazquez [50]<br>*Excluded due to number of patients <30* | Not described | 3000 mg/d | Not described | 2400±640 g/d (2000-3000) | 20-25 | Not described | <u>Overall therapy</u>:<br>19.5 (14.9-24.8)<br><u>24 h</u>:<br>15.1 (12.4-19.2);<br><u>48 h</u>:<br>19.5 (15.5-25.7)<br><u>72 h</u>:<br>22.8 (16.8-29.0)<br><u>Clinical condition</u>:<br><u>Serious</u>:<br>11.3 (10.4-13.8) (7/25);<br><u>Worse</u>:<br>12.1 (10.8-13.5) (2/25);<br><u>Improvement</u>:<br>23.5 (22.0-27.3) (16/25=; | Not described | <u>Mean</u>:<br>20.7 %<br><u>24 h</u>:<br>16.0 %<br><u>48 h</u>:<br>20.0 %<br><u>72 h</u>:<br>22.2 % | Routinely | Not described |
|  | Loading Dose (LD) | Maintenance Dose (MD) | Loading Dose (LD) | Maintenance Dose (MD) | Css [mg/L] | AUC24/MIC | Css [mg/L] (mean/median) | AUC24 [mg*h/L] (mean/median) | Target attainment [%] |  |  |
|  |  |  |  |  |  |  | *Apyretic:*<br>43.2 (41.7-44.8) (2/25) |  |  |  |  |
| **Wysocki [34]** | 15 mg/kg<br>(adjustment according to nomogram (Moellering, 1984 [10])) | 30 mg/kg/d;<br><u>VCM level <20 mg/L:</u><br>Increase daily dose by 500 mg;<br><u>VCM level >25 mg/L:</u><br>Decrease daily dose by 500 mg | Not described | Not described | 20-25 | Not described | <u>Overall therapy:</u> 24±8;<br><u>Success:</u> 23±4;<br><u>Failure:</u> 25±5<br><u>Time to target:</u> 36±31 h | 577±120<br><u>Failure:</u> 685±260;<br><u>Success:</u> 596±159 | Not described | Daily at 7:00 am until two consecutive VCM levels in target range, afterwards sampling only if 10 % change in BW or SCr | 13±5 |
| **Wysocki [117]**<br>*Excluded due to number of patients <30* | 15 mg/kg | 30 mg/kg/d | Not described | 24±14 mg/kg/d | 20-30 | Not described | <u>Overall therapy:</u> 24±6<br><u>Time to target:</u> 2.3±1.5 d | Not described | Not described | Daily at 7:00 am | 16±10 |
| **Yamada [29]**<br>*Excluded due to number of patients <30* | 30 mg/kg | 30 mg/kg/d | Not described | Not described | 15-25 | Not described | <u>12 h:</u><br>19.6 (14.4-23.7)<br><u>24 h:</u><br>23.59 (14.52-28.97);<br><u>36 h:</u><br>21.5 (12.2-30.0);<br><u>48 h:</u><br>18.28 (10.6-27.125) | 530.2 (108-958.4) | Not described | <u>First:</u><br>1 h after LD;<br><u>Regular:</u><br>12 h, 24 h, 36 h, 48 h after initiation of CI | Not described |

a: pneumonia, meningitis, endocarditis, ABW: adjusted body weight, AUC: area under the curve, AUC24/MIC: area under the curve of drug concentration during the first 24 hours of therapy divided by the minimal inhibitory concentration, AUC24: area under the curve of drug concentration during the first 24 hours of therapy, AUC96/MIC: area under the curve of drug concentration during the first 96 hours of therapy divided by the minimal inhibitory concentration, AUC96: area under the curve of drug concentration during the first 96 hours of therapy, BJI: bone and joint infection, (T)BW: (total) body weight, CI: continuous infusion of vancomycin, Css: concentration at steady state (12 h: Css about 12 hours after initiation of CI or on the 1<sup>st</sup> day of CI, sometimes called “day 0” or “day 1” in publications; 24 h: Css about 24 hours after initiation of CI or on the 2<sup>nd</sup> day of CI, sometimes called “day 1” or “day 2” in publications;
48 h: C<sub>ss</sub> about 48 hours after initiation of CI or on the 3<sup>rd</sup> day of CI, sometimes called “day 2” or “day 3” in publications; 72 h: C<sub>ss</sub> about 72 hours after initiation of CI or on the 4<sup>th</sup> day of CI, sometimes called “day 3” or “day 4” in publications), IBW: ideal body weight, LD: loading dose, MD: maintenance dose (24 h: MD on the 1<sup>st</sup> day of CI (= D1) or within the 1<sup>st</sup> 24 hours of CI, 48 h: MD on the 2<sup>nd</sup> day of CI (= D2) or within 24-48 hours after initiation of CI, 72 h: MD on the 3<sup>rd</sup> day of CI (= D3) or within 48-72 hours after initiation of CI), MIC: minimum inhibitory concentration, TDM: therapeutic drug monitoring, VCM: vancomycin.

### 9.4 Main findings Clinical and Microbiological Efficacy

**Table S4:** Overview of the clinical and microbiological efficacy of continuously applied vancomycin. Thirty-five studies were identified in the initial full-text selection process that presented data of steady-state serum concentrations of continuously administered vancomycin (CI) and efficacy (clinical or microbiological success (e.g. cure) or failure (e.g. mortality)). Only eighteen studies made it into the final analysis. Studies excluded from the final selection because of an increased risk of bias (e.g. unspecified or time course of less than 12 months or a CI study population of less than 30 patients) are written in italics.

| Reference | Category | Therapy Failure |  | Therapy Success |  | Independent predictors |  |
| --- | --- | --- | --- | --- | --- | --- | --- |
|  |  | Definition | Results | Definition | Results | Univariate | Multivariate |
| Akers [49] | Clinical failure<br>+ success | Mortality (in-hospital, 14-day,<br>28-day) | <u>In-hospital mortality:</u><br>All: 32.2 % (29/90);<br><u>Non-CVVH:</u> 19.1 % (13/68);<br><u>Gram-positive bacteremia:</u> 16<br>%, 4/25 / 4.4 %, 4/90;<br><u>Sepsis without gram-positive</u><br><u>bacteremia:</u> 70 %, 14/20 / 15.6<br>%, 14/90;<br><u>Pneumonia:</u> 35 %, 7/20 / 7.8 %,<br>7/90;<br><u>14-d mortality:</u><br>All: 10 % (9/90);<br><u>28-d mortality:</u><br>All: 18.9 % (17/90) | Not defined | Not defined | Not defined | Not defined |
|  | Microbiological<br>failure | Recurrence of gram-positive<br>bacteremia after beginning of<br>CI | <u>MSSA:</u> (2 of an unknown<br>quantity);<br><u>MRSA:</u> (2 of an unknown<br>quantity ); | Not defined | Not defined | Not defined | Not defined |
|  |  | Definition | Results | Definition | Results | Univariate | Multivariate |
|  |  |  | Enterococci: (3 of an unknown quantity )<br>CI+II: 17.5 % (10/57) |  |  |  |  |
| Albanèse [60]<br>Excluded due to number of patients <30 and unspecified length of time course | Clinical success | Not defined | Not defined | Cure;<br>Recovery | 100 % | Not defined | Not defined |
|  | Microbiological success | Not defined | Not defined | Bacteriological cure | 100 % | Not defined | Not defined |
| Beumier [55] | Clinical failure<br>+ success | Mortality (ICU) | 57 % (18/32) | Target attainment | 12 h: 69 %;<br>24 h: 63 %;<br>Subtherapeutic:<br>12 h: 3 %;<br>24 h: 19 %;<br>Supratherapeutic:<br>12 h: 28 %;<br>24 h: 19 % | Target attainment:<br>Body weight;<br>CRRT intensity | Not defined |
| Carricajo [62]<br>Excluded due to number of patients <30 and unspecified length of time course | Clinical success | Not defined | Not defined | Target attainment | Css:<br>All: 27.3 %;<br>≥120 mL/min: 0 %;<br><120 mL/min: 42.9 %;<br>Subtherapeutic:<br>All: 59.1 %;<br>≥120 mL/min: 100 %;<br><120 mL/min: 35.7 %;<br>Supratherapeutic:<br>All: 13.6 %;<br>≥120 mL/min: 0 %;<br><120 mL/min: 21.4 %;<br>AUC/MIC: | Not defined | Not defined |
|  |  | Definition | Results | Definition | Results | Univariate | Multivariate |
| | | | | | *All*: 66.7 %;<br>$\geq 120 \text{ mL/min}$: 25 %;<br>$< 120 \text{ mL/min}$: 87.5 % | | |
| Cianferoni<br>[56] | Clinical failure | Mortality (ICU) | *All*: 23 % (48/207);<br>*No AKI*: 18 % (28/157);<br>*AKI*: 40 % (20/50);<br>*(Early AKI)*: 46 % (13/28);<br>*Late AKI*: 32 % (7/22) | Not defined | Not defined | Not defined | Not defined |
| Covajes [58] | Clinical failure<br>+ success | Mortality (ICU) | 59 % (50/85) | Target attainment | **24 h**: 51 %;<br>**48 h**: 56 % ;<br>**72 h**: 67 %;<br>*Subtherapeutic*:<br>**24 h**: 29 %;<br>**48 h**: 22 % ;<br>**72 h**: 8 %;<br>*Supratherapeutic*:<br>**24 h**: 20 %;<br>**48 h**: 21 % ;<br>**72 h**: 26 % | **24 h target attainment**:<br>Age;<br>Daily VCM dose (mg/kg);<br>CRRT intensity | **24 h target attainment**:<br>Age;<br>Daily VCM dose (mg/kg);<br>CRRT intensity |
| Cristallini<br>[52] | Clinical failure<br>+ success | Mortality (ICU) | 22 % (24/107) | Target attainment | **12 h**: 56 %;<br>**24 h**: 54 %;<br>**48 h**: 73 % | Not defined | **24 h**:<br>BMI;<br>Body weight;<br>uCrCl |
| Di Filippo [63]<br>Excluded due to<br>number of<br>patients <30 and<br>time course of | Clinical success | Not defined | Not defined | Improvement<br>(= Reduction in SOFA score; leukocytes;<br>body temperature (T); tachycardia;<br>tachypnea) | *SOFA*: -33.8 % (before initiation of CI:<br>6.62±2.2; end of therapy: 4.37±3.5);<br>*Leukocytes*: -37.6 % (before initiation of<br>CI: 17.24±12.84; end of therapy:<br>10.76±3.61); | Not defined | Not defined |
|  |  | Definition | Results | Definition | Results | Univariate | Multivariate |
| only 7 months |  |  |  |  | <u>I</u>: 100 % (11/11);<br>Tachycardia: 100 % (11/11);<br>Tachypnea: 100 % (11/11);<br>(<u>CI+II</u>): Resolution of fever, tachycardia, tachypnea) |  |  |
|  | Microbiological success | Not defined | Not defined | Cure<br>(= Negative culture of the same type of biological sample) | 100 % (25/25) | Not defined | Not defined |
| Duszynska [31]<br>Excluded due to number of patients <30 | Clinical failure + success<br>(evaluation 24 h after end of CI) | Persistence or progression of clinical symptoms of infection;<br>Initial recovery followed by deterioration;<br>Change to broader spectrum antibiotic therapy (escalation);<br>Increased radiological infiltrates/computer tomography (CT)-scan pathology;<br>Worsening of laboratory data;<br>Need for surgery source control;<br>Catecholamines | <u>Css</u>:<br><u>All</u>: 19 % (4/21);<br><u>Reaching target at least once</u>: 17.6 % (3/17);<br><u>No reaching of target</u>: 25 % (1/4);<br><u>AUC24/MIC</u>:<br><u>≥400</u>: 20 % (1/5);<br><u>≤400</u>: 18.7 % (3/16) | Cure<br>(= Hemodynamic stabilization: resolution of clinical infection signs (body temperature < 37.5 °C); laboratory abnormalities (reduction of leukocytes, CRP, PCT concentration by more than 80 % compared to baseline); no further need for VCM) | <u>Css</u>:<br><u>All</u>: 81 % (17/21);<br><u>Reaching target at least once</u>: 82.4 % (14/17);<br><u>No reaching of target</u>: 75 % (3/4)<br><u>AUC24/MIC</u>:<br><u>>400</u>: 80 % (4/5);<br><u>≤400</u>: 81.3 % (13/16);<br><u>Clinical cure</u>: 194 (134-389) | Not defined | Not defined |
|  | Microbiological failure + success<br>(evaluation 24 h after end of CI) | Persistence colonization of new gram-positives or the same pathogen(s) in laboratory samples | <u>Css</u>:<br><u>All</u>: 19 % (4/21);<br><u>Reaching target at least once</u>: 23.5 % (4/17);<br><u>No reaching of target</u>: 0 % (0/4)<br><u>AUC24/MIC</u>:<br><u>≥400</u>: 20 % (1/5);<br><u>≤400</u>: 18.7 % (3/16) | Microbiological eradication<br>(= Negative culture from blood, intra-abdominal fluids, CSF, etc.) | <u>Css</u>:<br><u>All</u>: 81 % (17/21);<br><u>Reaching target at least once</u>: 76.5 % (13/17);<br><u>No reaching of target</u>: 100 % (4/4)<br><u>AUC24/MIC</u>:<br><u>>400</u>: 80 % (4/5);<br><u>≤400</u>: 81.3 % (13/16);<br><u>Microbiological eradication</u>: 162 (116-389) | Not defined | Not defined |
| Eldemiry [38]<br>Excluded due to number of patients <30 | Clinical failure + success | Mortality (15-day) | Not described | Improvement<br>(= Reduction in BUN; leukocytes; pCO<sub>2</sub>; body temperature (T); APACHE II; increase in pO<sub>2</sub>; spO<sub>2</sub>) | <u>BUN</u>: (-23.1 %; 46.8±20.27/36±18.71);<br><u>Leukocytes</u>: -40.6 % (16.82±13.23/9.99±3.78);<br><u>pCO<sub>2</sub></u>: -15.2 % (42.55±11.7/36.07±8.34);<br><u>T</u>: -3.1 % (reduction in 90 % of patients; 38.335±1.16/37.15±0.42);<br><u>APACHE II</u>: -30.3 % (14.2±4.6/9.9±5.54); | Not defined | Not defined |
|  |  | Definition | Results | Definition | Results | Univariate | Multivariate |
| | | | | | $pO_2$: +36.3 % (69.3±46.79/94.45±30.1);<br>$spO_2$: +22.2 % (76.94±17.8/94.01±11.03) | | |
|  | Microbiological success | - | - | Cure | 83.3 % (2/12) | - | - |
| Hanrahan [76] | Clinical failure | Mortality (ICU all-cause; within 72 h of last recorded VCM dose) | ICU: 26.3 % (172/653);<br><u>Within 72 h of last VCM:</u><br>19.9 % (130/653);<br><u>Mortality or nephrotoxicity within 72 h of cessation:</u><br>38.7 % (253/653) | Not defined | Not defined | Not defined | Not defined |
| Hong [26]<br>Excluded due to time course of only 9 months | Clinical failure + success | Mortality (ICU) | 15.4 % (10/65) | Target attainment | <u>24 h</u>: 40 %;<br>Subtherapeutic: 21.5 %<br>Supratherapeutic: 38.5 %<br><u>AUC<sub>24</sub>/MIC (*S. aureus*, 11/15):</u><br>MIC=0.5: 100 % (2/2);<br>MIC=1: 88.9 % (8/9);<br>MIC=2: 0 % (0/0)<br><u>Population-based:</u><br>MIC=0.5: 100 % (65/65);<br>MIC=1: 92.3 % (60/65);<br>MIC=2: 27.7 % (18/65)<br><u>Time to target</u>: 1.96 d;<br><u>Time within target</u>: 55 % | Not defined | Not defined |
|  | Microbiological success | - | - | Eradication | 55.5 % (11/20) | Not defined | Not defined |
| Hutschala [33] | Clinical failure + success | Mortality (ICU, in-hospital) | ICU: 21 % (25/119);<br><u>In-hospital</u>: 30.3 % (36/119);<br><u>Timing of death in course of VCM administration:</u> | Improvement<br>(= Reduction of CRP; leukocytes) | <u>CRP</u>: -66.4±41.6 %<br>(before CS: 3.7±5.1 (0.5-25.1);<br>at start of VCM: 22.64±10.1 | Not defined | Not defined |
|  |  | Definition | Results | Definition | Results | Univariate | Multivariate |
|  |  |  | 9±3.3 d (1-20) |  | (7.3-55);<br>on 10th day of VCM: 7.6±5.69<br>(0.7-32.1)) |  |  |
| **Lin [32]**<br>*Excluded due to number of patients <30* | Clinical failure + success | Mortality (in-hospital) | 50 % (7/14) | Target attainment;<br>Time to target | **24 h:** 100 %<br><u>Time to target:</u> 24±0.56 h | - | - |
| **Lin [42]** | Clinical failure + success | Mortality (in-hospital) | <u>Obese:</u> 19.2 % (5/26);<br><u>Non-obese:</u> 15.4 % (4/26) | Target attainment | **24 h:**<br><u>Obese:</u> 92.3 %;<br><u>Non-obese:</u> 88.5 %<br><u>CVVH</u><br><u>Obese:</u> 100 %;<br><u>Non-obese:</u> 100 % | Not defined | Not defined |
| **Ma [37]**<br>*Excluded due to inclusion of ICU and non-ICU patients in analysis* | Clinical failure + success | Mortality (infection-related, in-hospital) | <u>Infection-related:</u> 1 % (2/141);<br><u>All:</u> 2 % (3/141) | Target attainment<br>Cure<br>(= Resolution of infection signs and symptoms) | 78 %<br>93 % (62/67) | Not defined<br>Not defined | Not defined<br>Not defined |
|  | Microbiological success | Readmission within 14 d of discharge for infection recurrence;<br>Initiation of a new antibiotic with a similar spectrum due to lack of response | 7 % (3/40) | Cure<br>(= Negative culture during/up to 14 d after discontinuing CI after initial positive culture) | 93 % (37/40) | Not defined | Not defined |
| **Mohammedi [74]** | Clinical failure + success | Mortality (ICU) | <u>A (LD 500 mg):</u> 50 % (10/20);<br><u>B (LD 15 mg/kg):</u> 35 % (7/20) | Cure (in patients with proven infection) | <u>A (LD 500 mg):</u> 56 % (10/18);<br><u>B (LD 15 mg/kg):</u> 93 % (14/15) | Not defined | Not defined |
| **Ocampos-Martinez [59]** | Clinical failure + success | Mortality (ICU) | 29 % (77/261) | Target attainment | **24 h:** 47 %;<br>**48 h:** 67 %;<br>**72 h:** 86 %;<br>*Subtherapeutic:*<br>**24 h:** 42 %; | <u>Target attainment:</u><br>**24 h:**<br>CrCl;<br>Loading dose;<br>SCr; | <u>Target attainment:</u><br>**24-48 h:**<br>CrCl;<br>Gender (male) |
|  |  | Definition | Results | Definition | Results | Univariate | Multivariate |
|  |  |  |  |  | **48 h:** 14 %;<br>**72 h:** ND<br>*Supratherapeutic:*<br>**24 h:** 11 %;<br>**48 h:** 19 %;<br>**72 h:** ND | **48 h:**<br>CrCl;<br>Gender (male);<br>SCr;<br>SOFA |  |
| Omuro [61] | Clinical failure | Mortality (ICU) | <u>CI:</u> 25.2 % (30/119);<br><u>(VCM-AKI (CI+II):</u> 48 %;<br><u>Non VCM-AKI (CI+II):</u> 16.8 %) | Not defined | Not defined | <u>ICU Mortality:</u><br>AKI | Not defined |
| Perin [48] | Clinical failure<br>+ success | Mortality (ICU, in-hospital, 28-day) | **28-day:**<br><u>All:</u> 26 % (46/178);<br><u>D2/3 VCM level 20-25 mg/L:</u><br>19 % (8/42);<br><u>D2/3 VCM level <>20-25 mg/L:</u><br>28 % (37/132) | Target attainment | **24-48 h:** 24 % | <u>Target attainment:</u><br>Prolonged ICU stay before initiation of CI;<br><u>Subtherapeutic:</u><br>AKI on d1/2;<br>Prolonged ICU stay before initiation of CI | <u>Target attainment:</u><br>Prolonged ICU stay before initiation of CI;<br><u>Subtherapeutic:</u><br>AKI on d1/2 |
|  |  | Persistence or progression of baseline signs and symptoms of infection after ≥ 2 d of treatment, consistent with active infection<br>Requirement for additional antibacterial treatment | <u>Failure without identification:</u><br>13 % (23/179) | Cure<br>(= Resolution of clinical signs and symptoms compared with baseline)<br>(= No requirement for additional antibacterial treatment) | 65 % (116/179)<br>(test of cure could not be assessed in 13 % (23/179)) |  |  |
|  | Microbiological failure + success | Relapse<br>(= Finding of same pathogen);<br>Reinfection | <u>Relapse:</u> 3 % (5/179);<br><u>Reinfection</u> 7 % (12/179); | Not defined | Not defined | Not defined | Not defined |
|  |  | Definition | Results | Definition | Results | Univariate | Multivariate |
|  |  | (= Presence of another pathogen) |  |  |  |  |  |
| **Ricard [115]**<br>Excluded due to number of patients <30 and unspecified length of time course | clinical failure + success | Mortality (ICU) | 21.4 % (3/14) | Cure;<br>Improvement<br>(= Reduction of CSF leukocytes; CSF protein; CSF glucose); | Cure: 64.3 % (9/14);<br>Improvement: 14.3 % (2/14);<br>(Leukocyte: 1st lumbar puncture: 8±24;<br>2nd lumbar puncture: 0.7±0.11;<br>Protein: 1st: 8.6±7.22; 2nd: 5.4±4.35;<br>Glucose: 1st: 0.5±0.77; 2nd: 2.6±1.43) | Not defined | Not defined |
|  | Microbiological success | Not defined | Not defined | Cure<br>(= Eradication of pathogens in CSF) | Gram-positive cocci:<br>46 % (1st: 13; 2nd: 7/13);<br>S pneumoniae:<br>100 % (1st: 12; 2nd: 0/12) | Not defined | Not defined |
| **Saugel [54]**<br>Excluded due to time course of only 9 months | Clinical failure | Mortality (in-hospital; ICU) | ICU: 44 % (15/34);<br>Hospital: 59 % (20/34) | Not defined | Not defined | Not defined | Not defined |
| **Schmelzer [45]**<br>Excluded due to number of patients <30 and unspecified length of time course | Clinical success | Not defined | Not defined | Target attainment | 57.1 %<br>Subtherapeutic:<br>21.5 %<br>Supratherapeutic:<br>21.4 % | Not defined | Not defined |
| **Sin [41]** | Clinical failure<br>+ success | Mortality (in-hospital) | 63.5 % (33/52) | Target attainment | **24 h:** 82.7 %;<br>**48 h:** 81.8 %;<br>**72 h:** 75 %;<br>Subtherapeutic:<br>**24 h:** 3.8 %;<br>**48 h:** 6.1 %;<br>**72 h:** 8.3 %;<br>Supratherapeutic | Not defined | Not defined |
|  |  | Definition | Results | Definition | Results | Univariate | Multivariate |
|  |  |  |  |  | <u>24 h</u>: 13.5 %;<br><u>48 h</u>: 12.1 %;<br><u>72 h</u>: 16.7 % |  |  |
| Spadaro [43] | Clinical failure<br>+ success | Mortality (ICU) | <u>CrCl >50 mL/min</u>: 21.4 %<br>(45/210);<br><u>CrCl ≤50 mL/min</u>: 23.9 %<br>(33/138) | Target attainment | <u>24 h</u>:<br><u>CrCl >50 mL/min</u>: 44 %;<br><u>CrCl ≤50 mL/min</u>: 57 %;<br><u>48 h</u>:<br><u>CrCl >50 mL/min</u>: 41 %;<br><u>CrCl ≤50 mL/min</u>: 40 %<br><u>72 h</u>:<br><u>CrCl >50 mL/min</u>: 41 %;<br><u>CrCl ≤50 mL/min</u>: 41 % | Not defined | <u>Mortality</u>:<br>Subtherapeutic level at first VCM measurement (→ AUC/MIC was in therapeutic range);<br>SAPS II score |
| Spapen [46] | Clinical failure | Mortality<br>(no differentiation between "infection-related" or "AKI-induced") | <u>AKI</u>: 53 % (20/38);<br><u>No AKI</u>: 20 % (18/91) | Not defined | Not defined | Not defined | Not defined |
| Stepan [47] | Clinical failure<br>+ success | Mortality (infection-related) | 18 % (6/33) | Cure<br>(= Vanishing of all symptoms of an infection);<br>Improvement<br>(= Reduction of symptoms of infection; LOS; LOV; LOT;<br>Reduction of leukocytes;<br>Reduction of CRP) | <u>Cure/improvement</u>: 64 %<br>(21/33);<br><u>LOS</u>: 26 d (12.8-40.3);<br><u>LOV</u>: 25 d (10.5-39.3)<br><u>LOT</u>: 9 (7-10)<br><u>Leukocytes</u>: d0: 13.4 (9.7-18.9); end: 10.9 (8.6-17.9) (-18.7 %);<br><u>CRP</u>: d0: 192.0±98.1; end: 123±83.1 (-35.9 %) | Not defined | Not defined |
|  |  | Persistence of symptoms;<br>Increase of symptoms | Not described |  |  |  |  |
|  |  | Definition | Results | Definition | Results | Univariate | Multivariate |
|  | Microbiological failure + success | Persistency;<br>Superinfection;<br>Relapse | Not described | Cure<br>(=Eradication;<br>Suspected eradication) | 76 % (25/33) | Not defined | Not defined |
| **Tiede [116]**<br>*Excluded due to number of patients <30* | Clinical success<br>(study not designed for efficacy evaluation) | Not defined | Not defined | Cure;<br>Improvement<br>(= Reduction of infection parameter within 7 d after initiation of Ct: CRP (serum), leukocyte (serum, CSF)) | Cure: 100 % (9/9);<br>CRP: 100 % (9/9);<br>Leukocyte serum: overall decrease, but not in every patient;<br>Leukocyte CSF: 100 % (9/9) | Not defined | Not defined |
| **Udy [57]** | Clinical failure | Mortality (not specified) | 58 % (47/81) | Not defined | Not defined | Not defined | Not defined |
| **van de Vijzel [39]**<br>*Excluded due to number of patients <30* | Clinical failure + success | Mortality (ICU, infection-related) | ICU: 92 % (20/22);<br>Infection-related: 59 % (13/22) | Target attainment | 71 % | Not defined | Not defined |
| **van den Heever [40]**<br>*Excluded due to number of patients <30* | Clinical failure + success | Mortality (ICU) | 10 % (1/10) | Target attainment | 50 % after 21 h;<br>100 % within 33 h | Not defined | Not defined |
| **Vazquez [50]**<br>*Excluded due to number of patients <30* | Clinical failure + success | Worsening of clinical condition;<br>Serious condition | Worsening: 8 % (2/25)<br>Serious: 28 % (7/25) | Improvement of clinical condition;<br>Afebrile | Improvement: 64 % (16/25);<br>Apyretic: 8 % (2/25) | Not defined | Not defined |
| **Wysocki [34]** | Clinical failure + success | Mortality (infection-related;<br>10-day; end of treatment;<br>overall: 10-day; end of treatment; ICU) | **Infection-related:**<br>D10: 8 % (5/61);<br>End of treatment: 10 % (6/61);<br>**Overall:**<br>D10: 8 % (5/61);<br>End of treatment: 18 % (11/61);<br>ICU: 37 % (21/61) | Cure;<br>Improvement | 78.7 % (48/61) | Not defined | Not defined |
|  |  | Persistency or impairment of | D10: 21 % (13/61); |  |  |  |  |
|  |  | Definition | Results | Definition | Results | Univariate | Multivariate |
|  |  | clinical, laboratory, radiological statuses | <u>End of treatment</u>: 21 % (13/61) |  |  |  |  |
|  | Microbiological failure + success | Presence of *Staphylococcus* spp. or other species in specimen on day 5 | <u>Staphylococcus</u>: 39 % (24/61);<br><u>Other species</u>: 15 % (9/61);<br><u>Bacteremia</u>: 28 % (17/61) | Cure<br>(= Eradication of *Staphylococcus* spp. in specimen on day 5) | 46 % (28/61) | Not defined | Not defined |
| Wysocki [117]<br>Excluded due to number of patients <30 | Clinical failure + success | Mortality (infection-related, overall) | <u>Infection-related</u>:<br>15 % (0-33) (2/13)<br><u>ICU</u>: 38 % (11-65) (5/13) | Duration of fever (<38.5 °C) | 3 d (1-8) | Not defined | Not defined |
|  | Microbiological success | Not defined | Not defined | Duration of bacteremia | 2 d (1-7) | Not defined | Not defined |
| Yamada [29]<br>Excluded due to number of patients <30 | Clinical failure | Mortality (not specified) | 36 % (4/11) | Not defined | Not defined | Not defined | Not defined |

AKI: acute kidney injury, APACHE: Acute Physiology and Chronic Health Evaluation, BUN: blood urea nitrogen, BW: body weight, CI: continuous infusion of vancomycin, (u)CrCl: creatinine clearance (measured on daily urine excretion), CRP: C-reactive protein, CRRT: continuous renal replacement therapy, CSF: cerebrospinal spinal fluid, LOS: length of stay, LOT: length of therapy with vancomycin, LOV: length of ventilation, PCO2: partial pressure of carbon dioxide, PCT: procalcitonin, PO2: partial pressure of oxygen, SOFA: sepsis-related organ failure, VCM: vancomycin.

### 9.5 Main findings Safety

**Table S5:**
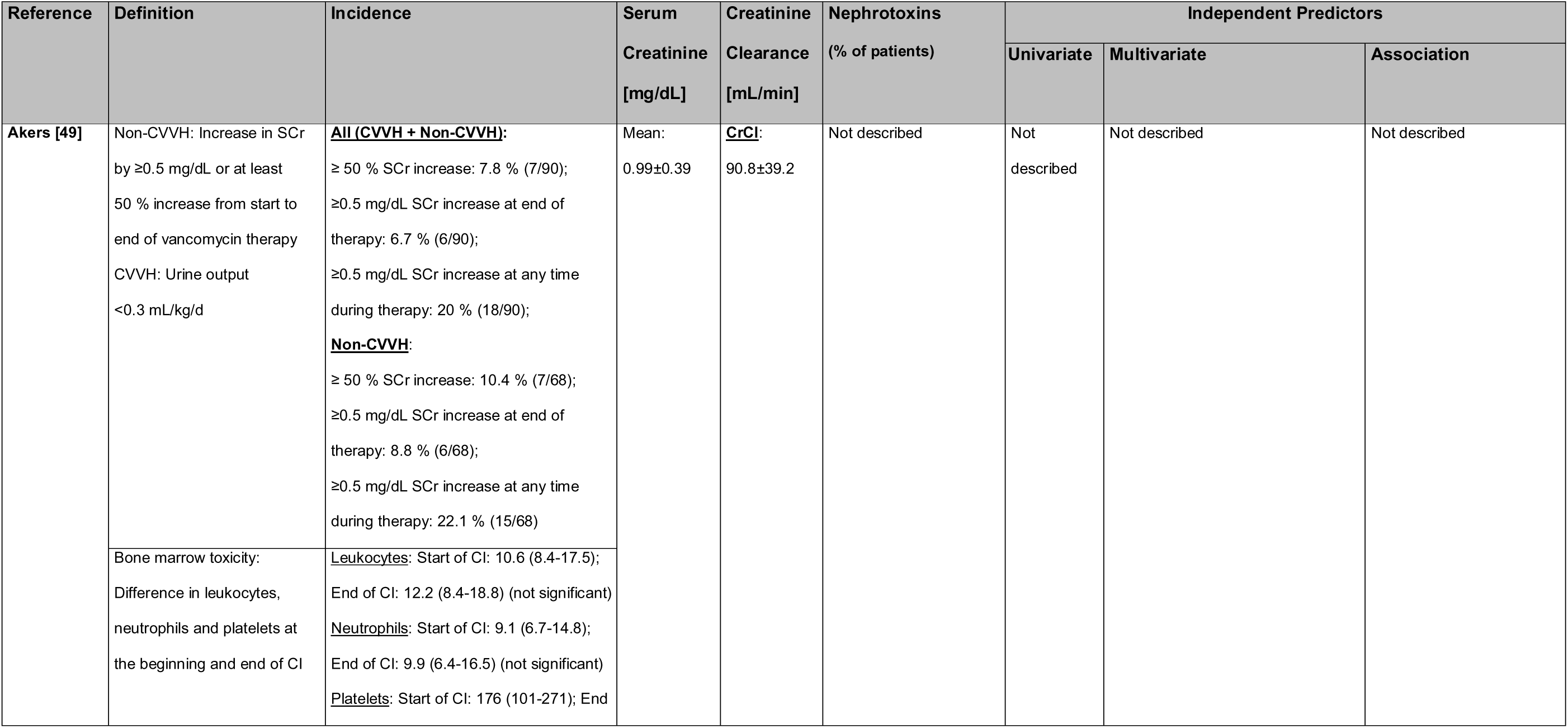
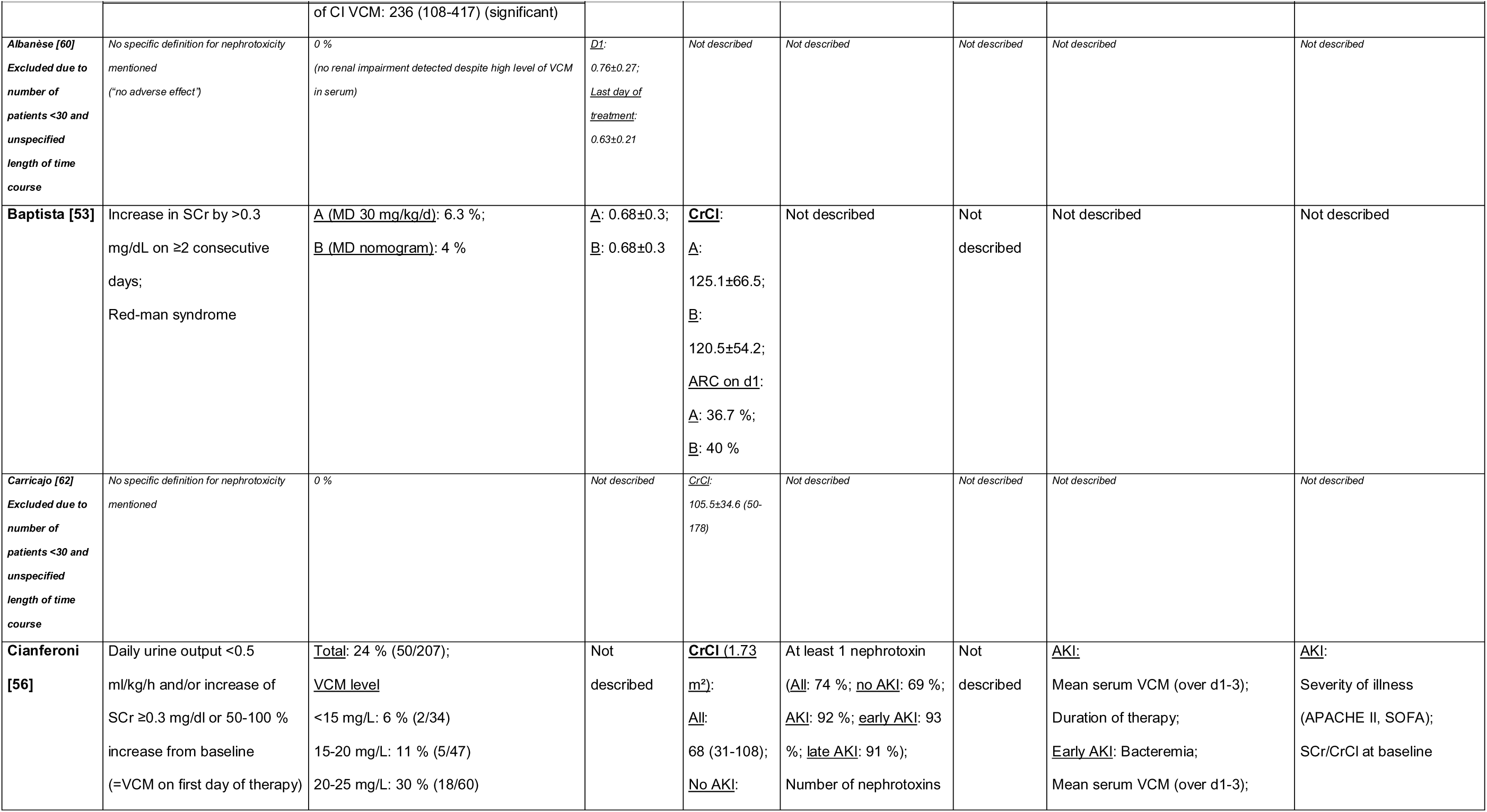
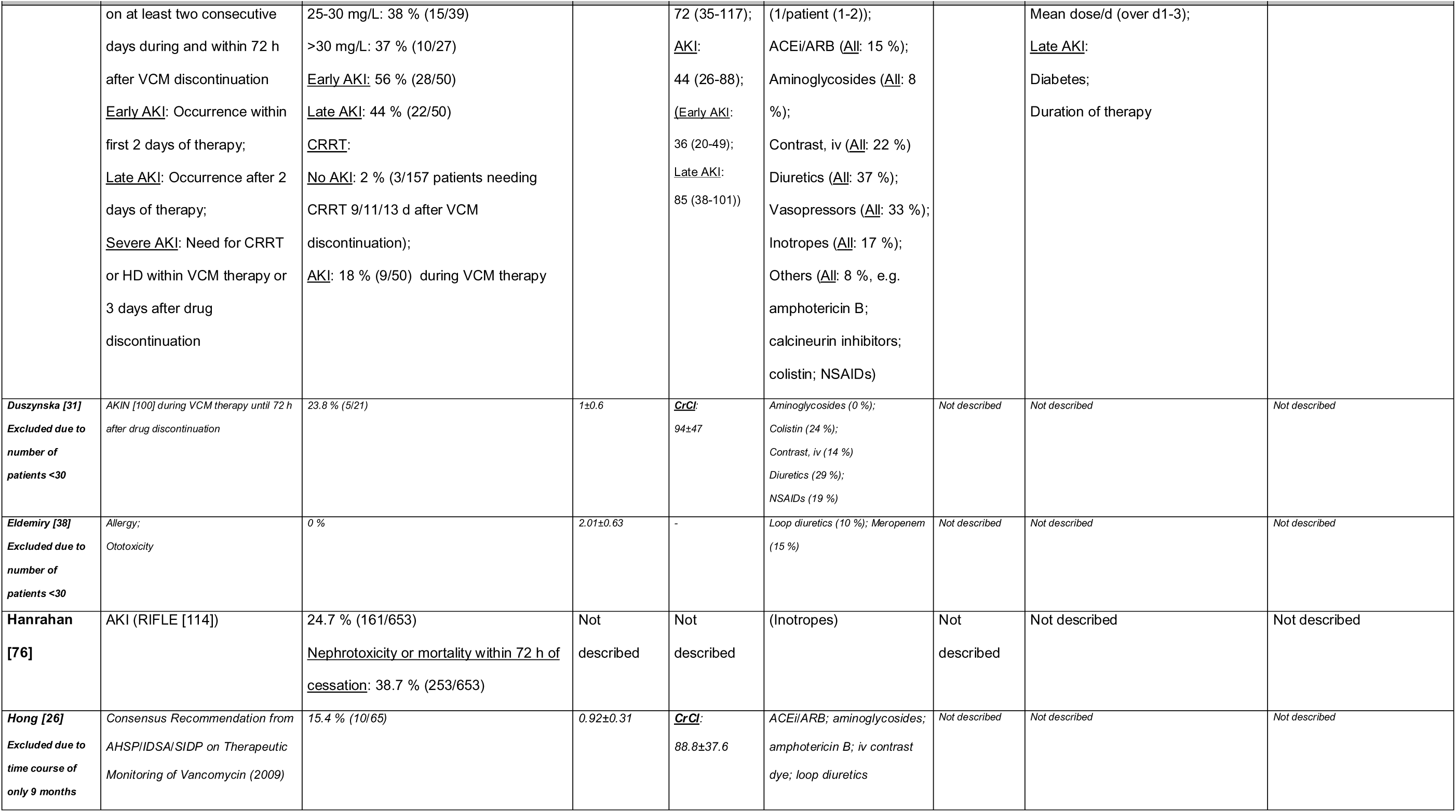
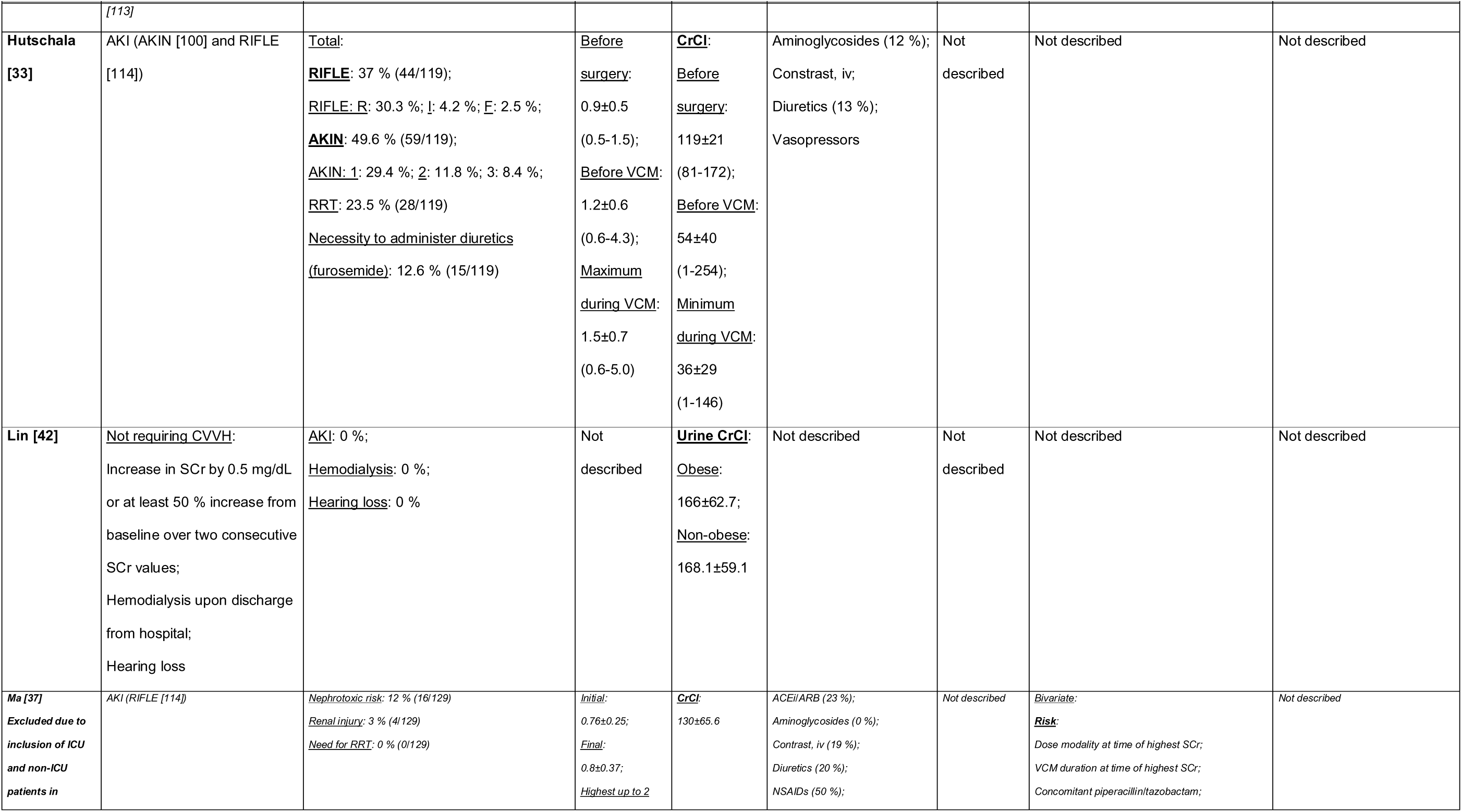
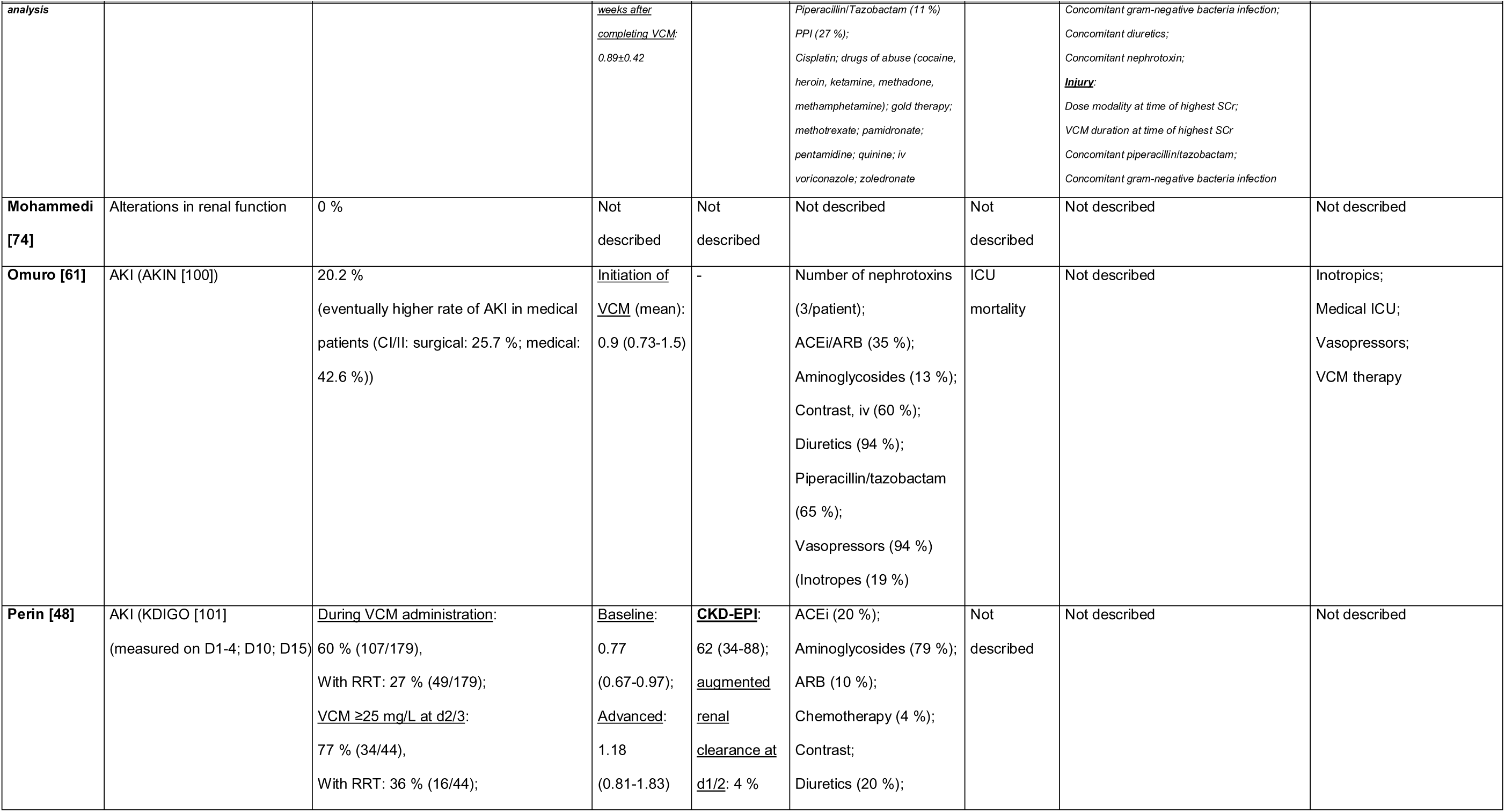
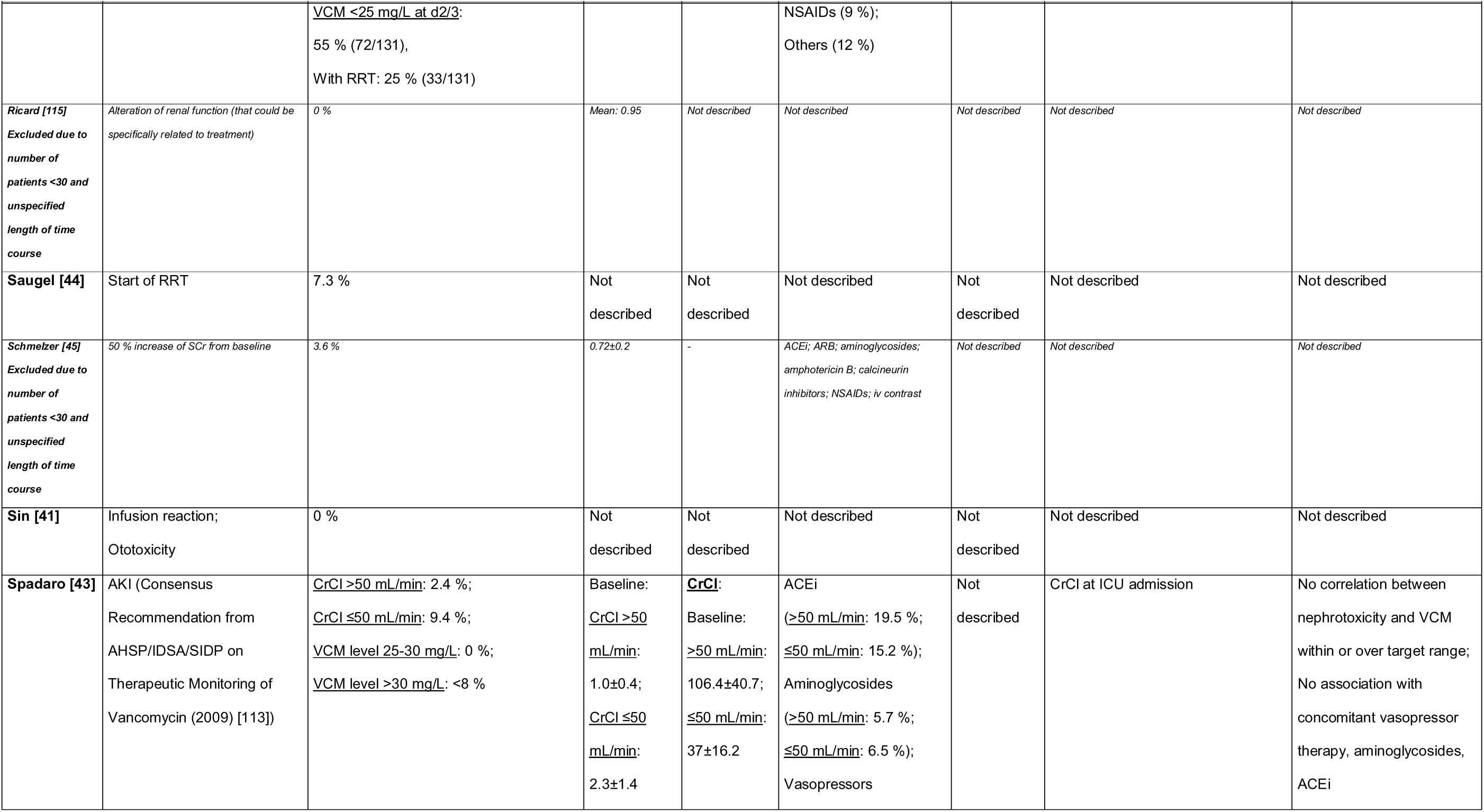
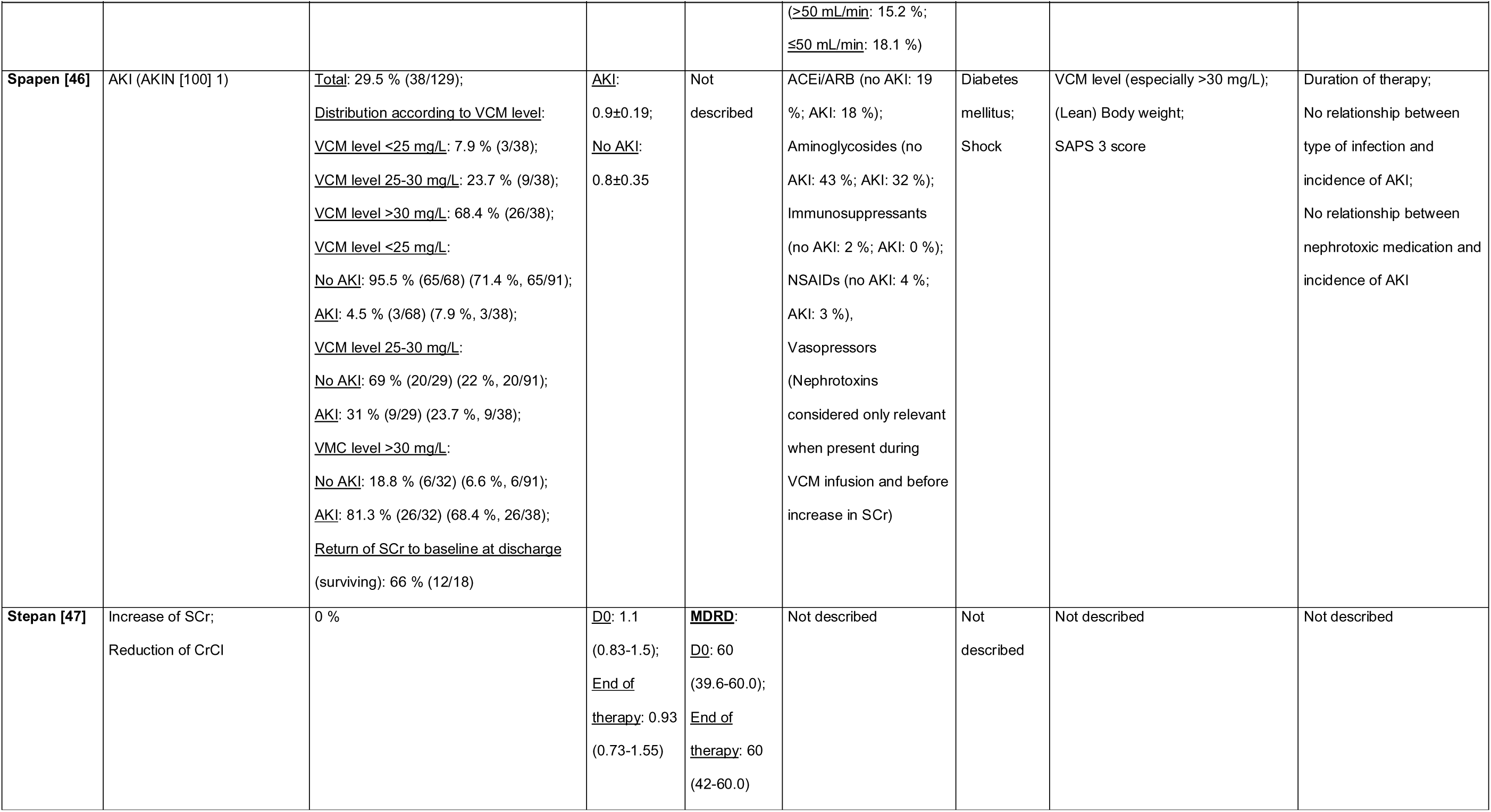
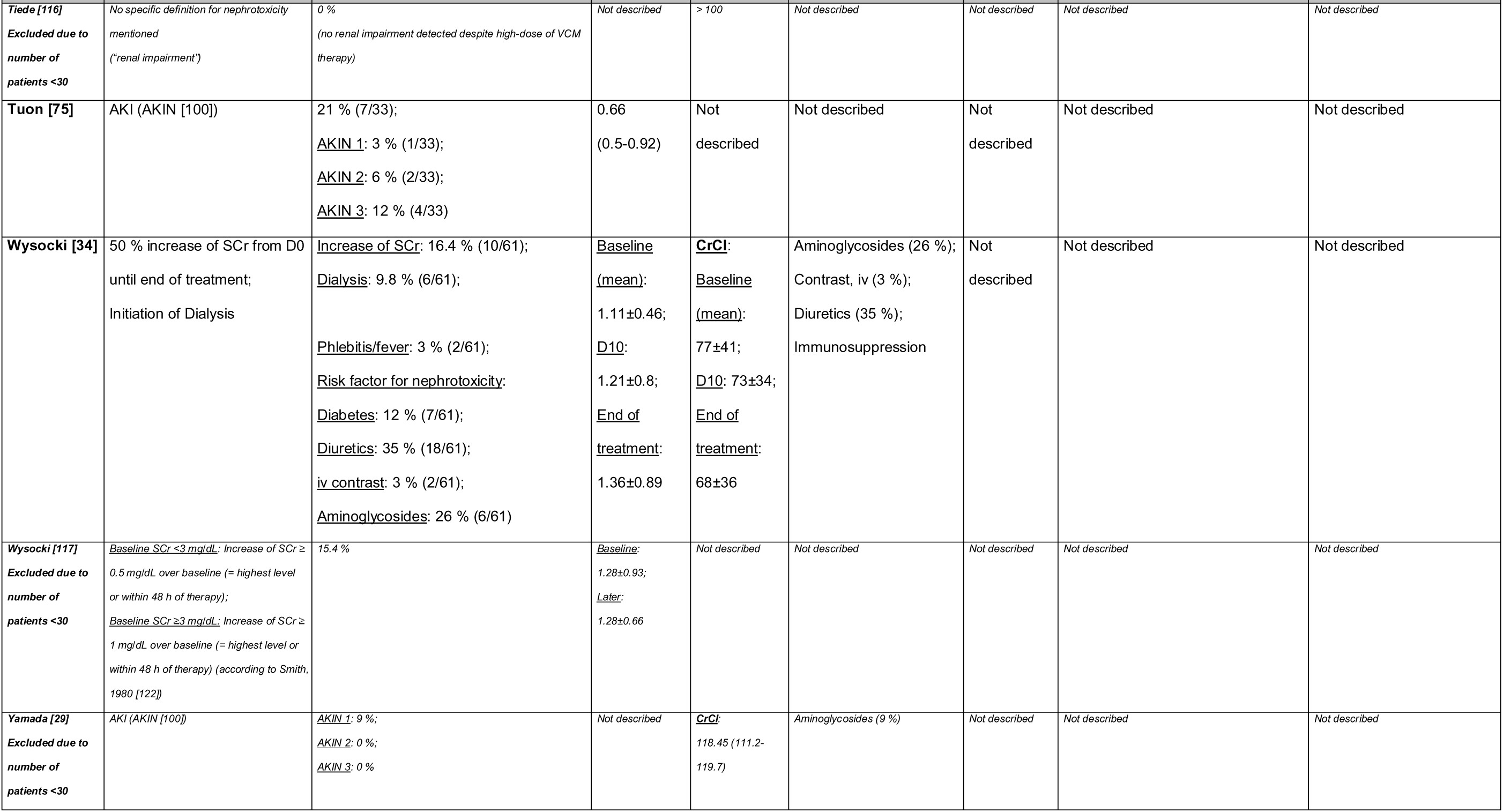
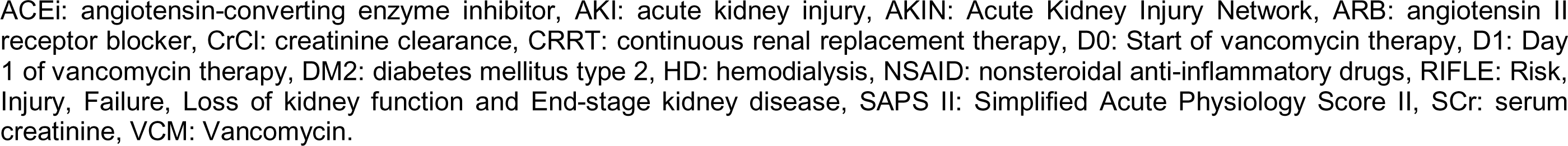
Overview of the safety of continuously applied vancomycin. Twenty-seven studies were identified in the initial full-text selection process that presented data of steady-state serum concentrations of continuously administered vancomycin (CI) and safety (e.g. nephrotoxicity in the form of acute renal failure). Only sixteen studies made it into the final analysis. Studies excluded from the final selection because of an increased risk of bias (e.g. unspecified or time course of less than 12 months or a CI study population of less than 30 patients) are written in italics.

**Table S6:**
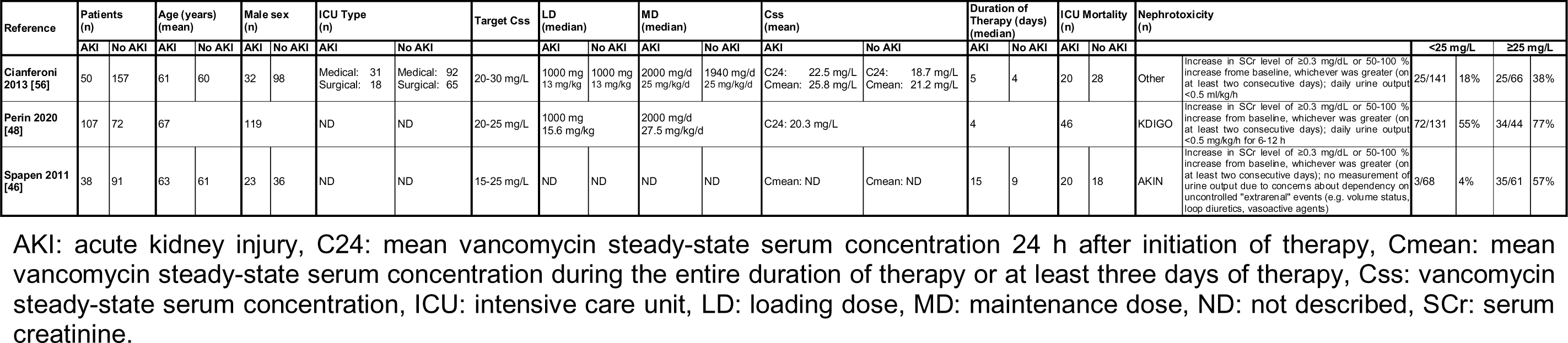
Characteristics of studies included in the forest plot analysis comparing the influence of vancomycin steady-state serum concentration below or above 25 mg/L on nephrotoxicity (= acute kidney injury) (n=3).

**Table S7:** Comparison of the influence of vancomy concentration below or above 25 mg/L on the rate of acut.

| Reference | RR | Lower limit | Upper limit | Z-value | P-value |
| --- | --- | --- | --- | --- | --- |
| Cianferoni 2013 [56] | 0.468 | 0.292 | 0.750 | -3.159 | 0.0016 |
| Perin 2020 [48] | 0.711 | 0.569 | 0.889 | -2.995 | 0.0027 |
| Spapen 2011 [46] | 0.077 | 0.025 | 0.237 | -4.460 | <0.0001 |
| Summary | 0.535 | 0.432 | 0.662 | -5.745 | <0.0001 |

RR: relative risk, RR <1 indicates a lower rate of acute kidney injury at a vancomycin serum concentration of <25 mg/L in contrast to >25 mg/L. P-value <0.05 was considered significant. Calculations according to Altman 2011 [73]

### 9.6 Quality Assessment

**Table S8:**
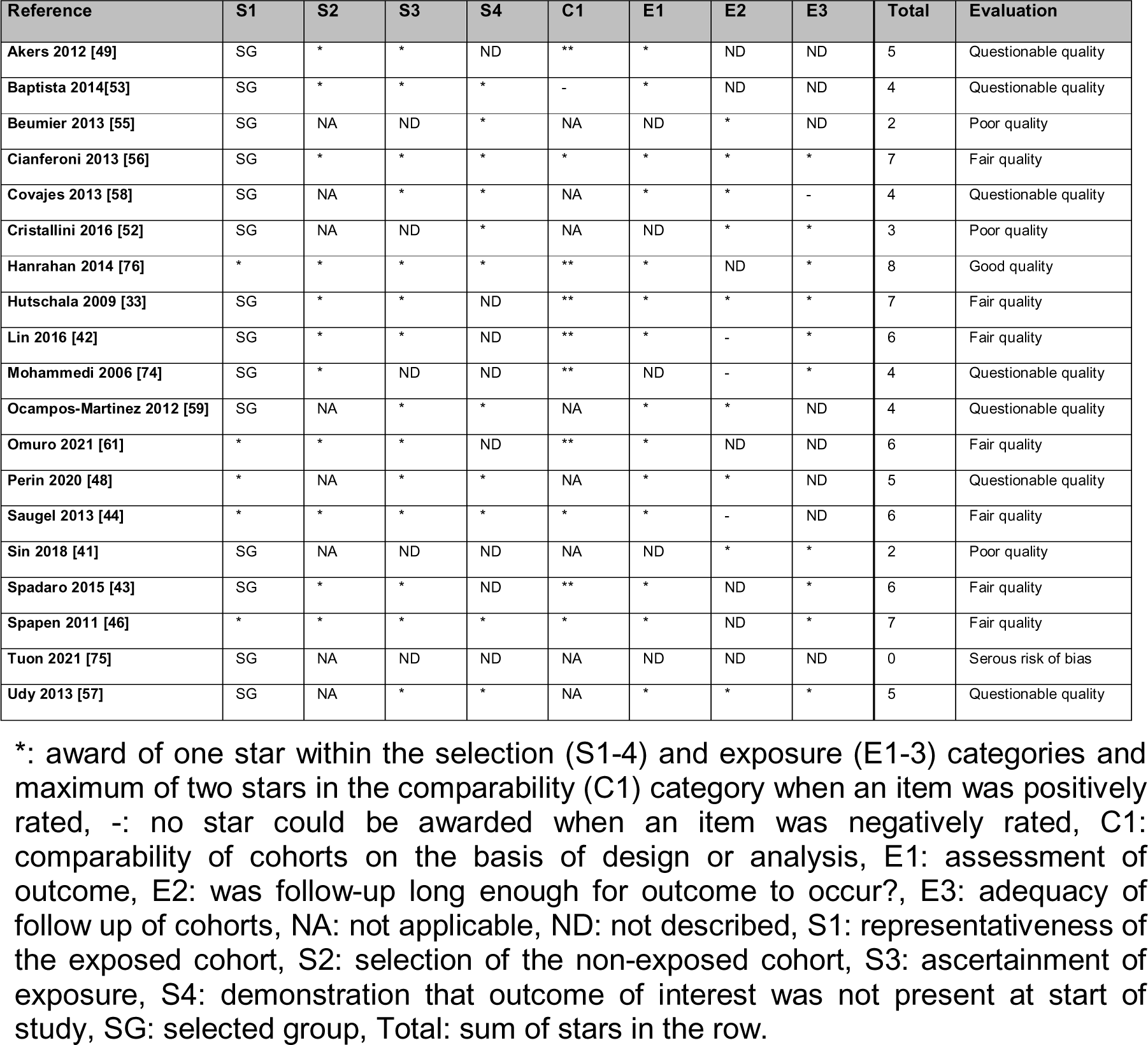
Quality assessment of the non-randomized studies using the Newcastle-Ottawa Scale (NOS). A star system described the suitability of the selection of the study groups (S1-S4) (max. four stars), the comparability of the groups (C1) (max. two stars) and the ascertainment of the exposure or outcome of interest (E1-E3) (max. three stars). The total number of stars was interpreted as “good quality” at 8-9 stars, “fair quality” at 6-7 stars, “questionable quality” at 4-5 stars, “poor quality” at 2-3 stars and “serious risk of bias” at 0-1 stars.

| Reference | S1 | S2 | S3 | S4 | C1 | E1 | E2 | E3 | Total | Evaluation |
| --- | --- | --- | --- | --- | --- | --- | --- | --- | --- | --- |
| Akers 2012 [49] | SG | * | * | ND | ** | * | ND | ND | 5 | Questionable quality |
| Baptista 2014[53] | SG | * | * | * | - | * | ND | ND | 4 | Questionable quality |
| Beumier 2013 [55] | SG | NA | ND | * | NA | ND | * | ND | 2 | Poor quality |
| Cianferoni 2013 [56] | SG | * | * | * | * | * | * | * | 7 | Fair quality |
| Covajes 2013 [58] | SG | NA | * | * | NA | * | * | - | 4 | Questionable quality |
| Cristallini 2016 [52] | SG | NA | ND | * | NA | ND | * | * | 3 | Poor quality |
| Hanrahan 2014 [76] | * | * | * | * | ** | * | ND | * | 8 | Good quality |
| Hutschala 2009 [33] | SG | * | * | ND | ** | * | * | * | 7 | Fair quality |
| Lin 2016 [42] | SG | * | * | ND | ** | * | - | * | 6 | Fair quality |
| Mohammedi 2006 [74] | SG | * | ND | ND | ** | ND | - | * | 4 | Questionable quality |
| Ocampos-Martinez 2012 [59] | SG | NA | * | * | NA | * | * | ND | 4 | Questionable quality |
| Omuro 2021 [61] | * | * | * | ND | ** | * | ND | ND | 6 | Fair quality |
| Perin 2020 [48] | * | NA | * | * | NA | * | * | ND | 5 | Questionable quality |
| Saugel 2013 [44] | * | * | * | * | * | * | - | ND | 6 | Fair quality |
| Sin 2018 [41] | SG | NA | ND | ND | NA | ND | * | * | 2 | Poor quality |
| Spadaro 2015 [43] | SG | * | * | ND | ** | * | ND | * | 6 | Fair quality |
| Spapen 2011 [46] | * | * | * | * | * | * | ND | * | 7 | Fair quality |
| Tuon 2021 [75] | SG | NA | ND | ND | NA | ND | ND | ND | 0 | Serous risk of bias |
| Udy 2013 [57] | SG | NA | * | * | NA | * | * | * | 5 | Questionable quality |

\*: award of one star within the selection (S1-4) and exposure (E1-3) categories and maximum of two stars in the comparability (C1) category when an item was positively rated, -: no star could be awarded when an item was negatively rated, C1: comparability of cohorts on the basis of design or analysis, E1: assessment of outcome, E2: was follow-up long enough for outcome to occur?, E3: adequacy of follow up of cohorts, NA: not applicable, ND: not described, S1: representativeness of the exposed cohort, S2: selection of the non-exposed cohort, S3: ascertainment of exposure, S4: demonstration that outcome of interest was not present at start of study, SG: selected group, Total: sum of stars in the row.

**Table S9:**
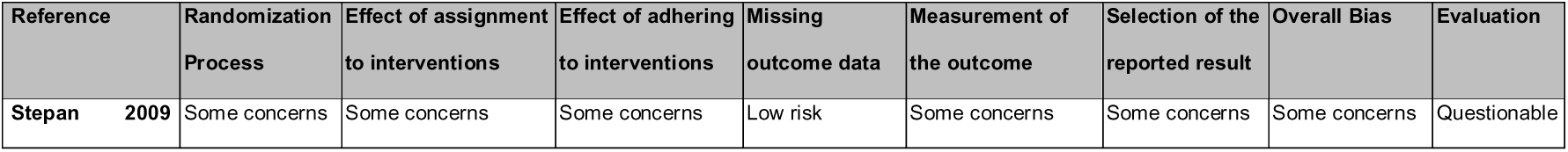
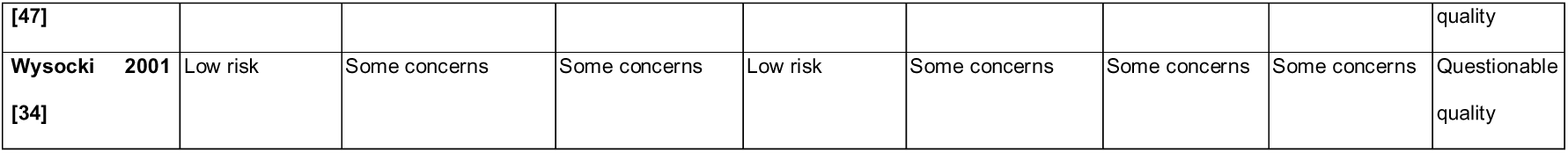
Quality assessment of the randomized trials using the revised Cochrane risk of bias tool (RoB 2). The overall bias “some concerns” was interpreted as “questionable quality” in the summary of the quality assessment results.

### 9.7 Efficacy

**Table S10:**
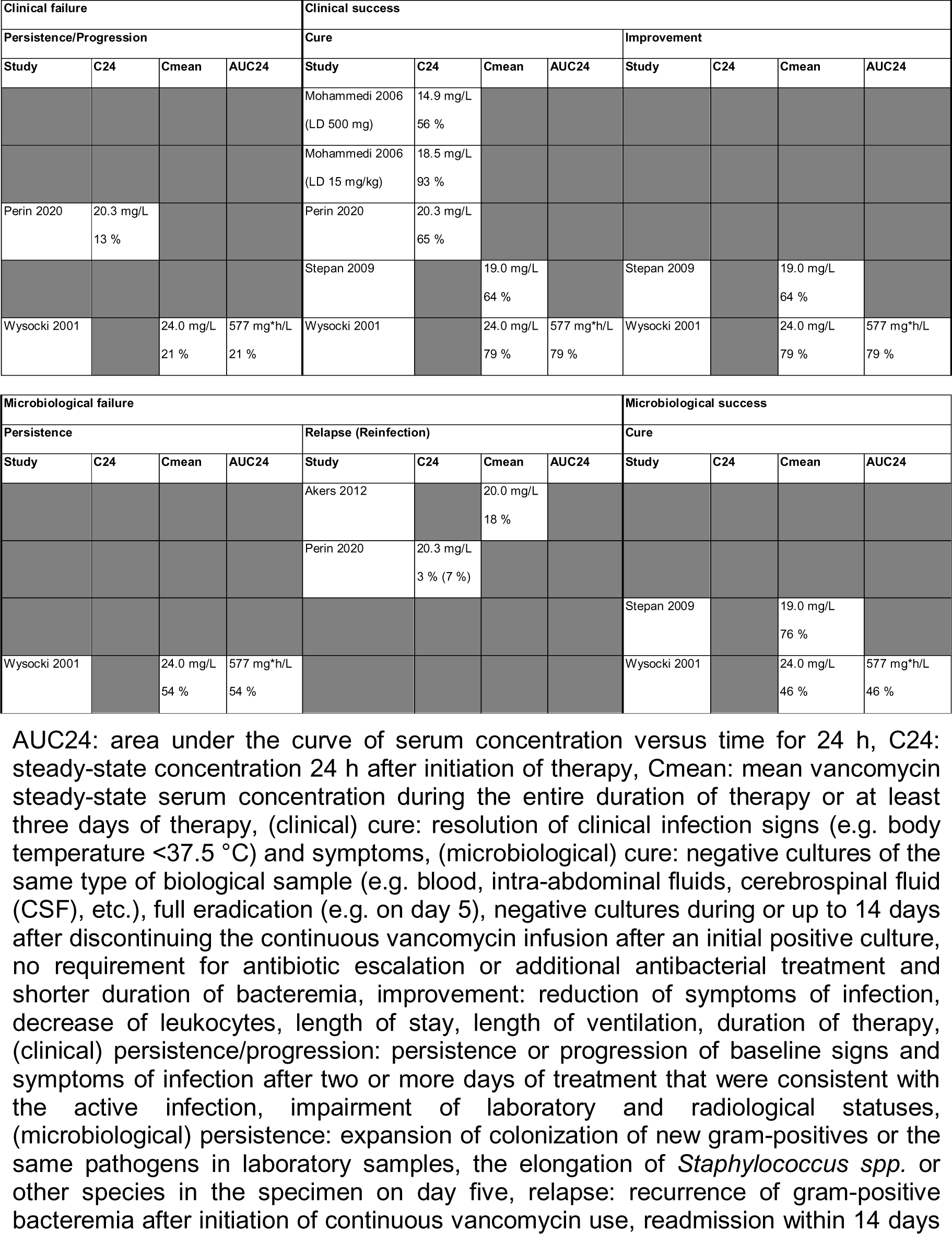
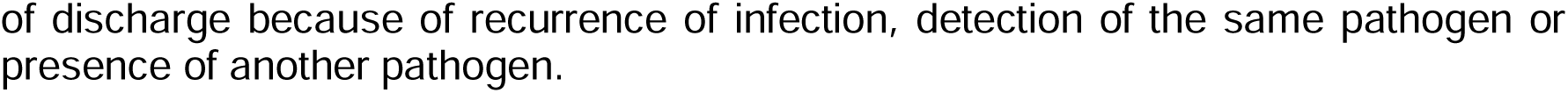
Overview of the clinical failure rates other than mortality, the clinical success rates and the microbiological failure and success rates in comparison to the vancomycin steady-state serum concentrations presented in the eighteen studies included in the efficacy analysis of continuously administered vancomycin.

**Table S11:** Overview of the clinical and microbiological failure and success parameters and frequency ranges presented in the eighteen studies included in the efficacy analysis of continuously administered vancomycin.

| Parameter |  | Definition | Range | Reference |
| --- | --- | --- | --- | --- |
| Clinical failure | Mortality | Intensive care unit (ICU) | 21-59 % | [33, 46, 47, 58, 63, 70, 71, 73, 75, 98, 100] |
|  |  | In-hospital | 17-63 % | [46, 47, 56, 57, 63, 64] |
|  |  | X-Day (10-day, 14-day, 28-day, within 72 h of last recorded vancomycin dose) | 8-26 % | [47, 63, 64, 100] |
|  |  | Infection-related | 8-18 % | [47, 62] |
|  | Persistence or progress | Persistence or progression of baseline signs and symptoms of infection after two or more days of treatment that were consistent with the active infection, impairment of laboratory and radiological statuses | 13-21 % | [47, 62, 63] |
| Clinical success | Cure | Resolution of clinical infection signs (e.g. body temperature <37.5 °C) and symptoms | 64-93 % | [46, 47, 62, 63, 98] |
|  | Improvement | Reduction of symptoms of infection, decrease of leukocytes, length of stay (LOS), length of ventilation (LOV), duration of therapy (LOT) |  |  |
| Microbiological failure | Persistence | Expansion of colonization of new gram-positives or the same pathogens in laboratory samples and the elongation of *Staphylococcus spp.* or other species in the specimen on day five | 3-54 % | [34, 48, 49] |
|  | Escalation | Initiation of a new antibiotic with a similar spectrum to vancomycin due to lack of response and superinfection |  |  |
|  | Relapse | Recurrence of gram-positive bacteremia after initiation of continuous vancomycin use, readmission within 14 days of discharge because of recurrence of infection, detection of the same pathogen or presence of another pathogen |  |  |
| Microbiological success | Cure | Negative cultures of the same type of biological sample (e.g. blood, intra-abdominal fluids, cerebrospinal fluid (CSF), etc.) or during or up to 14 days after discontinuing CI after an initial positive culture, full eradication (e.g. on day 5), no requirement for antibiotic escalation or additional antibacterial treatment and shorter duration of<br>bacteremia | 46-91 % | [47, 62, 63] |

### 9.8 Additional Plots

**Figure S1:**
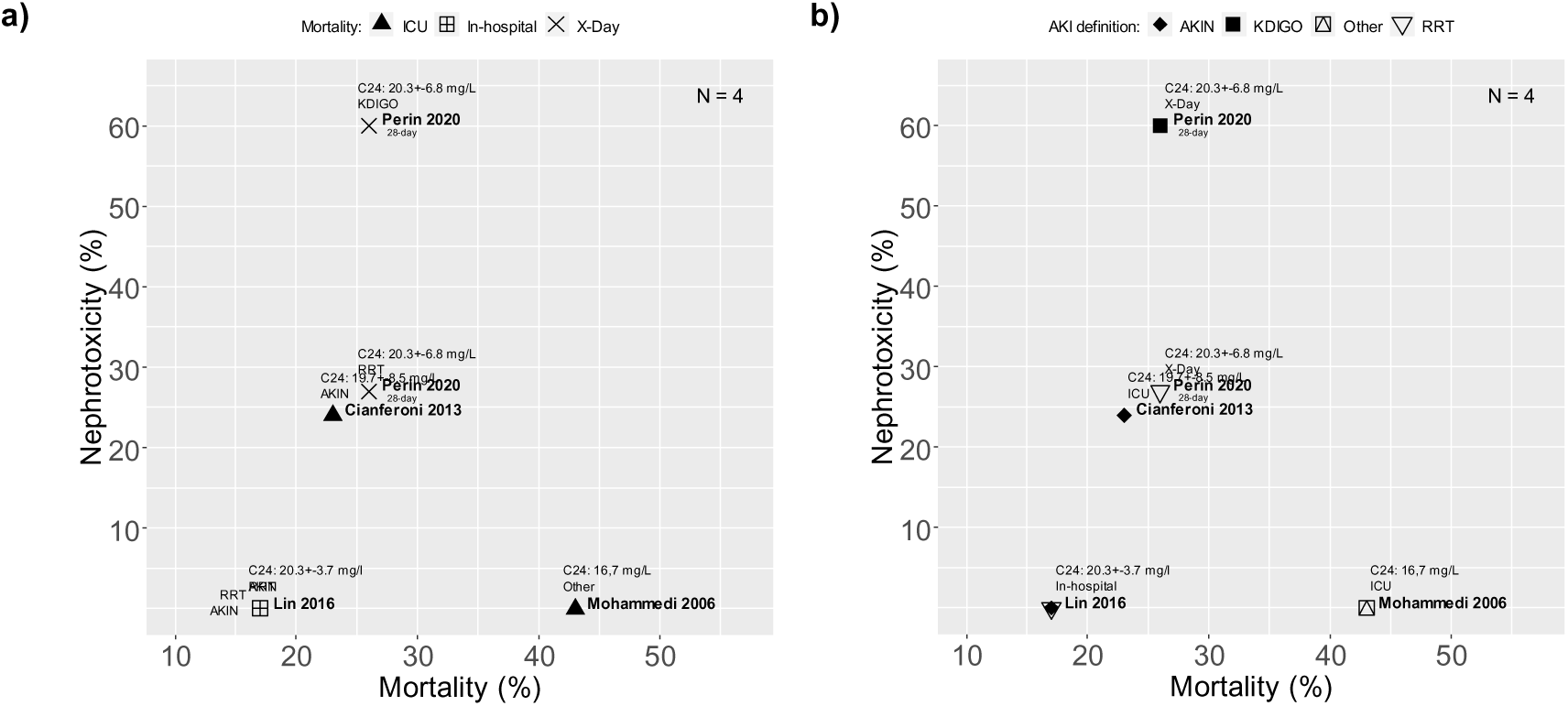
Scatterplots of the mortality rate plotted against the rate of acute kidney injury (AKI) (= nephrotoxicity) for studies in which mean vancomycin serum levels measured approximately 24 h after initiation of therapy with continuous infusion of vancomycin (C24) were reported (n=4 studies representing 478 patients). **a)** Differentiation according to the type of mortality. **b)** Differentiation according to the AKI definition. Filled black triangle: ICU mortality, bordered cross: in-hospital mortality, X: 28-day mortality, filled black diamond: AKIN, filled black square: KDIGO, squared triangle: other definition of AKI, non-filled triangle: RRT AKI: acute kidney injury, AKIN: Acute Kidney Injury Network, ICU: intensive care unit, KDIGO: Kidney Disease Improving Global Outcomes, RRT: renal replacement therapy, X-Day: 28-day.

**Figure S2:**
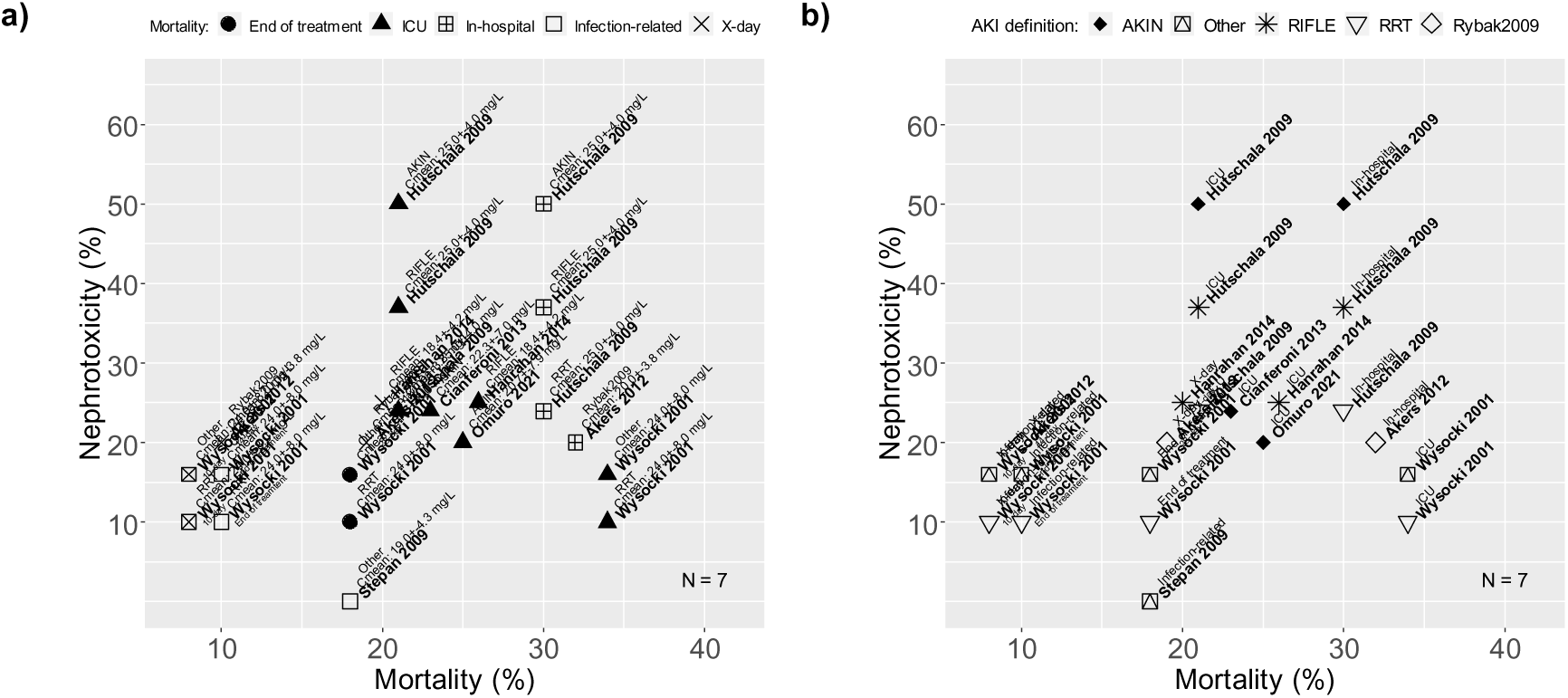
Scatterplots of the mortality rate plotted against the rate of acute kidney injury (AKI) (= nephrotoxicity) for studies in which mean vancomycin serum levels calculated from the average of the levels obtained during the entire duration of therapy with continuous infusion of vancomycin or at least three days of therapy (Cmean) were reported (n=7 studies representing 1,282 patients). **a)** Differentiation according to the type of mortality. **b)** Differentiation according to the AKI definition. Filled black circle: mortality at the end of treatment with vancomycin, filled black triangle: ICU mortality, bordered cross: in-hospital mortality, non-filled square: infection-related mortality, X: 28-day mortality, filled black diamond: AKIN, squared triangle: other definition of AKI, star: RIFLE, non-filled triangle: RRT, non-filled diamond: Rybak2009 AKI: acute kidney injury, AKIN: Acute Kidney Injury Network, ICU: intensive care unit, KDIGO: Kidney Disease Improving Global Outcomes, RIFLE: Risk, Injury, Failure, Loss, End Stage Renal Disease, RRT: renal replacement therapy, Rybak2009: Consensus Recommendation from AHSP/IDSA/SIDP on Therapeutic Monitoring of Vancomycin (2009) (AHSP: American Society of Health-System Pharmacists, IDSA: Infectious Diseases Society of America, SIDP: Society of Infectious Diseases Pharmacists).

**Figure S3:**
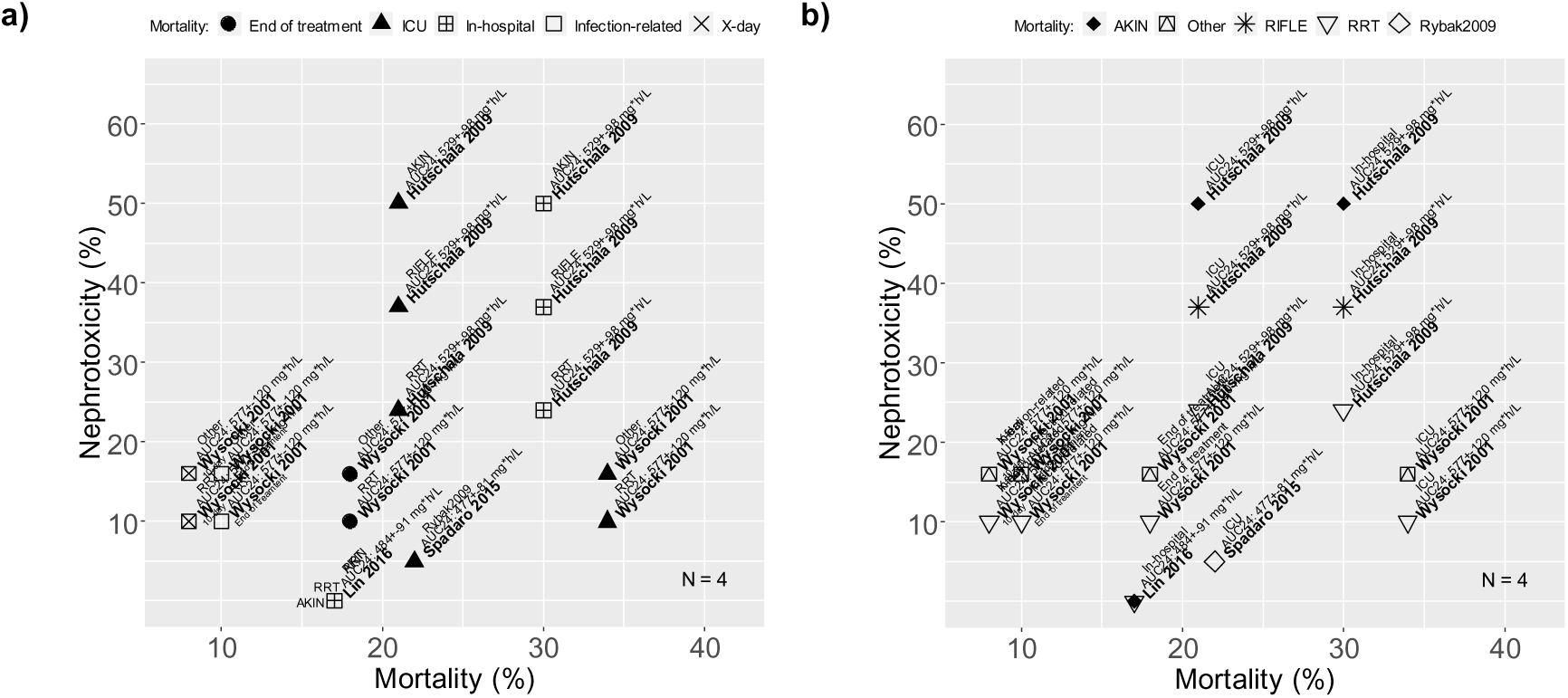
Scatterplots of the mortality rate plotted against the rate of acute kidney injury (AKI) (= nephrotoxicity) for studies in which the mean vancomycin areas under the serum concentration vs. time curve for 24 h during therapy with continuous infusion of vancomycin (AUC24) were reported (n=4 studies representing 580 patients). **a)** Differentiation according to the type of mortality. **b)** Differentiation according to the AKI definition. Filled black circle: mortality at the end of treatment with vancomycin, filled black triangle: ICU mortality, bordered cross: in-hospital mortality, non-filled square: infection-related mortality, X: x-day mortality (e.g. 10-day, 28-day, 30-day), filled black diamond: AKIN, squared triangle: other definition of AKI, star: RIFLE, non-filled triangle: RRT, non-filled diamond: Rybak2009 AKI: acute kidney injury, AKIN: Acute Kidney Injury Network, ICU: intensive care unit, KDIGO: Kidney Disease Improving Global Outcomes, RRT: renal replacement therapy, RIFLE: Risk, Injury, Failure, Loss, End Stage Renal Disease, RRT: renal replacement therapy, Rybak2009: Consensus Recommendation from AHSP/IDSA/SIDP on Therapeutic Monitoring of Vancomycin (2009) (AHSP: American Society of Health-System Pharmacists, IDSA: Infectious Diseases Society of America, SIDP: Society of Infectious Diseases Pharmacists).

**Figure S4:**
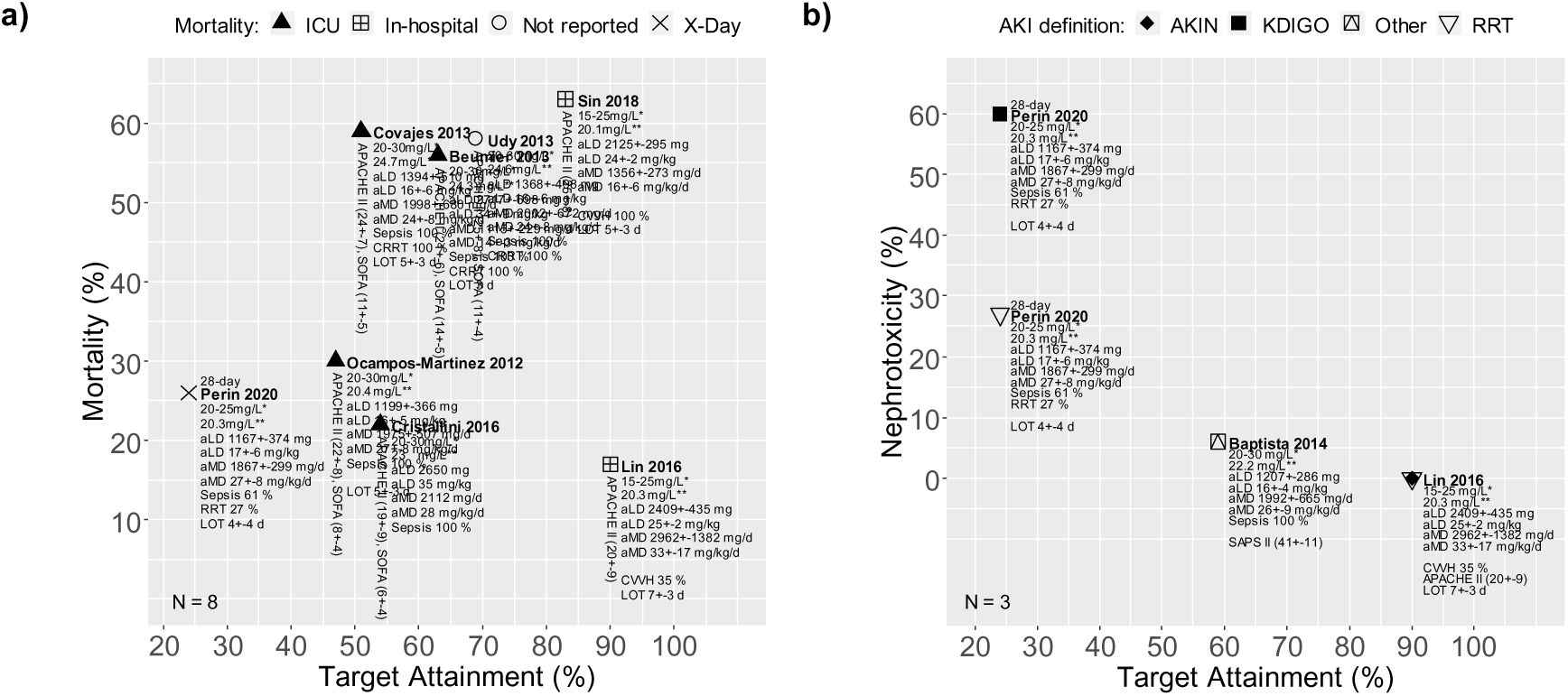
**a)** Scatterplot of studies in which a mean vancomycin level measured approximately 24 h after initiation of therapy with continuous infusion of vancomycin (C24), a target attainment rate and a mortality rate were reported (n=8 studies representing 768 patients). **b)** Scatterplot of studies in which a C24, a target attainment rate and a rate of acute kidney injury (= nephrotoxicity) were reported (n=3 studies representing 335 patients). Filled black triangle: ICU mortality, bordered cross: in-hospital mortality, non-filled circle: type of mortality was not reported, X: x-day mortality (e.g. 10-day, 28-day, 30-day), filled black diamond: AKIN, filled black square: KDIGO, squared triangle: other definition of AKI, non-filled triangle: RRT, * target range, ** C24 AKI: acute kidney injury, AKIN: Acute Kidney Injury Network, aLD: applied average loading dose, aMD: applied average maintenance dose, APACHE II: Acute Physiology And Chronic Health Evaluation II score, C24: mean vancomycin level measured approximately 24 h after initiation of therapy with continuous infusion of vancomycin, CVVH: continuous veno-venous hemofiltration, ICU: intensive care unit, KDIGO: Kidney Disease Improving Global Outcomes, LOT: length of therapy, RIFLE: Risk, Injury, Failure, Loss, End Stage Renal Disease, (C)RRT: (continuous) renal replacement therapy, Rybak2009: Consensus Recommendation from AHSP/IDSA/SIDP on Therapeutic Monitoring of Vancomycin (2009) (AHSP: American Society of Health-System Pharmacists, IDSA: Infectious Diseases Society of America, SIDP: Society of Infectious Diseases Pharmacists), SAPS II: Simplified Acute Physiology Score II, SOFA: Sepsis-related Organ Failure Assessment score.

**Figure S5:**
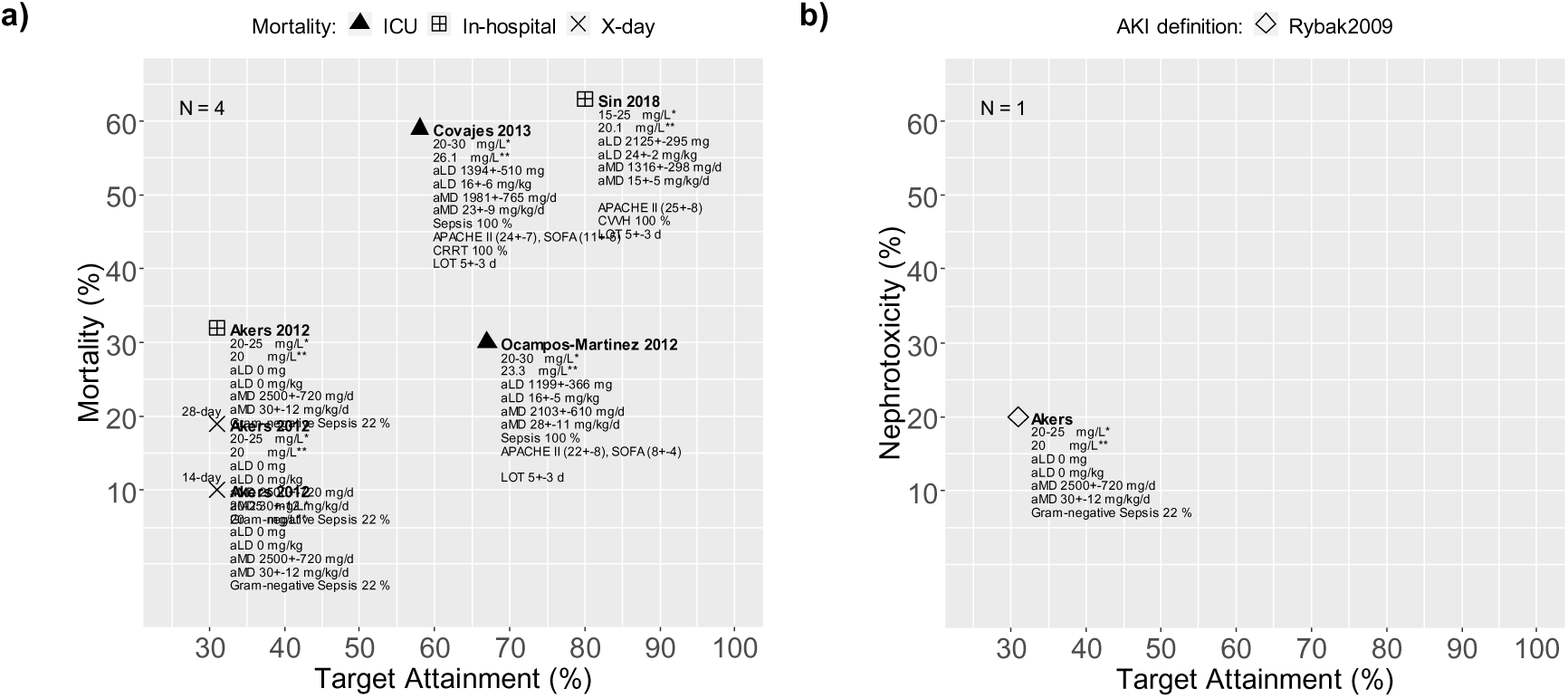
**a)** Scatterplot of studies in which a mean vancomycin steady-state serum level during the entire duration of continuously administered vancomycin or at least three days of therapy (Cmean), a target attainment rate and a mortality rate were reported (n=4 studies representing 488 patients). **b)** Scatterplot of studies in which a Cmean, a target attainment rate and a rate of acute kidney (= nephrotoxicity) injury were reported (n=1 study representing 90 patients). Filled black triangle: ICU mortality, bordered cross: in-hospital mortality, X: x-day mortality (e.g. 14-day, 28-day), non-filled diamond: Rybak2009, * target range, ** Cmean AKI: acute kidney injury, AKIN: Acute Kidney Injury Network, aLD: applied average loading dose, aMD: applied average maintenance dose, APACHE II: Acute Physiology And Chronic Health Evaluation II score, Cmean: mean vancomycin steady-state serum level during the entire duration of continuously administered vancomycin or at least three days of therapy, ICU: intensive care unit, KDIGO: Kidney Disease Improving Global Outcomes, LOT: length of therapy, RIFLE: Risk, Injury, Failure, Loss, End Stage Renal Disease, (C)RRT: (continuous) renal replacement therapy, Rybak2009: Consensus Recommendation from AHSP/IDSA/SIDP on Therapeutic Monitoring of Vancomycin (2009) (AHSP: American Society of Health-System Pharmacists, IDSA: Infectious Diseases Society of America, SIDP: Society of Infectious Diseases Pharmacists).

**Figure S6:**
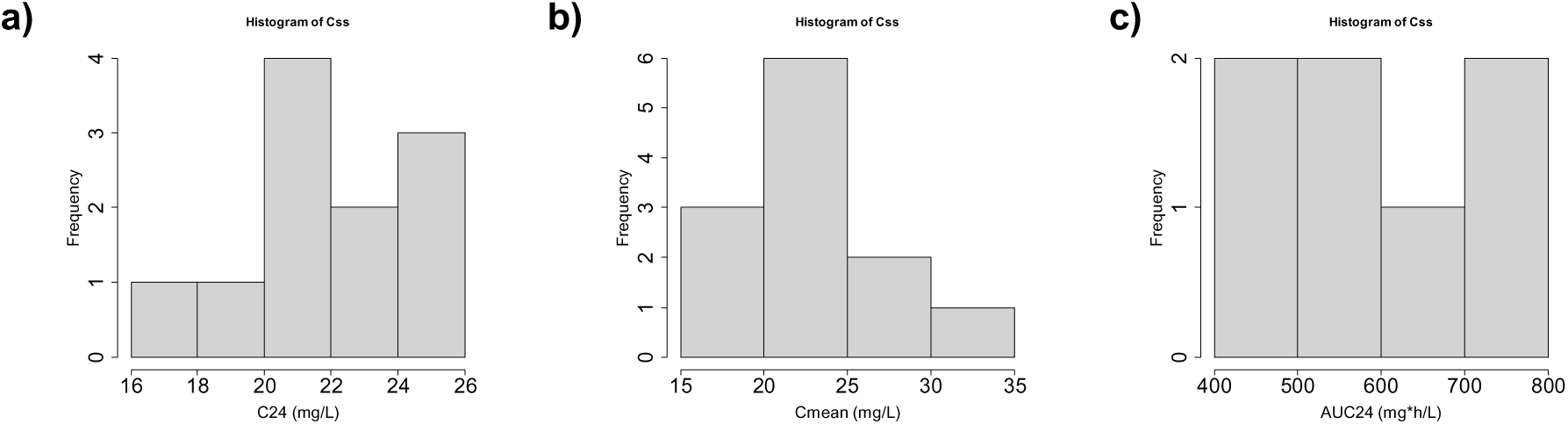
**a)** Distribution of studies for the mean vancomycin serum levels measured approximately 24 hours after initiation of therapy with continuously administered vancomycin (C24) (n=11 studies representing 1,200 patients). **b)** Distribution of studies for the mean vancomycin steady-state serum levels during the entire duration or at least three days of therapy with continuously administered vancomycin (Cmean) (n=12 studies representing 1,877 patients). **c)** Distribution of studies for the mean vancomycin area under the serum concentration-time curve for a period of 24 hours (AUC24) (n=7 studies representing 752 patients).

